# Examining the indirect effects of life satisfaction and perceived social support on selection optimization compensation and PTSD among the senior citizens of Ekiti State: A moderated mediation approach

**DOI:** 10.1101/2024.01.31.24301758

**Authors:** Dogbahgen Alphonso Yarseah, Ololade Omolayo Ogunsanmi, Joyce Olufunke Ogunsanmi, Alade Foladade Adesola, Falana Bernard Akinilabi, Olu Francis Ibimiluyi, Viola H. Cheeseman

**Affiliations:** Ekiti State University, Faculty of Education, Department of Guidance and Counseling Ekiti State Nigeria; Department of Public Health Babcock University, Ogun State, Nigeria; Ekiti State University, Faculty of Education Department of Guidance and Counseling Ekiti State, Nigeria; Ekiti State Universisty, Faculty of Education, Department of Guidance and Counseling, Ekiti State, Nigeria; Ekiti State University, Faculty of Education, Department of Guidance and Counseling Ado Ekiti, Nigeria; Wuhan University

**Keywords:** Aging, SOC, PTSD, life satisfaction, perceived social support

## Abstract

This study addresses the escalating concern of Posttraumatic Stress Disorder (PTSD) among the elderly population in Ekiti State, Nigeria. As the country experiences a growing number of older individuals, understanding and mitigating PTSD in this demographic is of paramount importance. Despite this urgency, there is a significant gap in the literature pertaining to PTSD among older Nigerians. To fill this void, our research investigates the impact of Selection, Optimization, and Compensation (SOC) strategies on perceived social support (PSS), life satisfaction (LS), and PTSD in the elderly.

Our study explores the relationships between PSS and LS, considering their potential roles in alleviating the psychological impact of trauma-related disorders. We also examine how age groups moderate the relationship between SOC and PTSD. Additionally, we explore the indirect effects of PSS and LS on the SOC-PTSD relationship.

Conducting a cross-sectional study, we collected data from 321 individuals aged 65 and above (260 males, 61 females) across four Local Government Areas. Our data collection instruments included socio-demographic variables, the SOC instrument, the PTSD scale, the Multidimensional scale of PSS, and the LS index-z.

Utilizing the Hayes Model 5 macro process for Structural Equation Modeling (SEM) analysis, our results highlight the significant moderating effect of age groups on the SOC-PTSD relationship. Furthermore, we find that PSS and LS partially mediate the relationships between SOC and PTSD, with SOC directly influencing both PTSD and PSS. These findings carry substantial practical implications and provide avenues for future research. In light of our study, we offer recommendations for addressing the mental health needs of Ekiti State’s elderly population, emphasizing the importance of further studies in the critical field of geropsychology.

## Introduction

Posttraumatic stress disorder (PTSD) remains a relatively underexplored subject within the African context, particularly among senior citizens in Nigeria. PTSD, a mental health challenge distinguished by intrusive memories of traumatic events, emotional withdrawal, and hyperarousal, can persist for at least a month following such an experience (Owens et al, 2005). Among older individuals, typical PTSD symptoms include sleep disturbances, physical discomfort, hyperarousal, and cognitive impairment, and use of terms such as “stress” to indicate trauma-related struggles (Sledjeski, Speisman, & Dierker,2008; Qureshi; 2009). Research reveals that while senior citizens may encounter PTSD symptoms similar to those experienced by adults, distinct symptom clusters emerge (Averill & Beck, 2000). In aging adults, heightened arousal and reduced re-experiencing symptoms are observed (Cook & Niederehe, 2007). Over time, the prevalence of re-experiencing symptoms diminishes, contrasting with the increasing prevalence of avoidance symptoms (McFarlane, 2000). In older adults, PTSD often coexists with other psychiatric conditions, including depression (Alamdari & Lagana, 2015). Older individuals with PTSD and depression tend to exhibit a history of recurrent depressive episodes, a greater incidence of chronic physical health conditions, heightened health-related impairment and diminished life satisfaction (Hegel et al., 2005).

Clinical studies focusing on senior citizens have highlighted unique patterns in their expression of distress following traumatic experiences. Senior citizens tend to report physical discomfort such as pain, sleep disturbances, gastrointestinal issues, and cognitive difficulties (Owens et al., 2005). More general terms such as “stress” may be used to describe trauma-related challenges while words such as depression or anxiety may be used less frequently to articulate emotional struggles (Avrill & Beck, 2000). Notably, there may be a tendency among older adults to disconnect current concerns from past traumatic experiences, which may have occurred long ago (Owens et al., 2005). Senior participants in research studies have reported experiencing distress when exposed to signals associated with trauma, such as loud noises or news about military conflicts. A diminished interest in customary activities has also been noted (Avrill & Beck, 2000). A similar trend has been identified in studies on depression among Nigerians, where symptoms are often described as sensations of heaviness or heat in the head, crawling sensations in the head or legs, bodily burning sensations, and perception of abdominal bloating (Ebiognos, 1982).

Previous research conducted in Nigeria has illuminated the prevalence of PTSD among various groups, including displaced individuals, urban residents, and victims of road traffic accidents. Prevalence rates range from 2.7% to 66.7% (Sekonil, Mall, & Chirstofides 2021; Sheikh, et al., 2019; Iteke, et al 2011; Asuquo, et al., 2017), and certain sociodemographic factors such as age, marital status, and wealth have beenshown to be associated with PTSD occurrence [Sekonil, Mall & Chirstofides, 2021).

The reasons for the high level of PTSD in Africa have been tied to violent conflicts and lack of development and government legislation (Nag et al., 2020; Hilft, 2017; Marc, N.D). People living in Africa are asymmetrically affected by manmade and natural disasters (Nag et al,.2020). Research from the World Health Organization (WHO) has revealed that the lifetime prevalence of road traffic deaths (WHO, 2015; 2008), intimate partnership violence, and nonpartner sexual violence is high on the continent of Africa (Abrahams, 2014). Additionally, African countries have experienced more natural disasters (Hift, 2017) and fight wars and asymmetrical wars as well as other armed conflicts for decades than other continents (Marc, N.D).

In 2019, more than 50% of the fragile and conflict-affected countries in sub-Saharan African nations were documented by the World Bank as hosting fragile and conflict-affected situations (World Bank, 2019). In the year 2023, Sub-Saharan Africa recorded the most significant increase in terrorism deaths, which was 60% of all terrorism deaths globally (Global Terrorism Index 2023), with four of the ten countries with the most considerable deterioration in Global Terrorism Index scores in region of Sub-Saharan Africa including Togo, Benin, Djibouti, and Center African Republic. More importantly, the Sahel region in Sub-Saharan Africa is now the nexus of terrorism, accounting for more terrorism deaths in 2022 than South Asia, the Middle East, or North Africa combined. Approximately 43% of the global deaths recorded in 2022 were in the Sahel region of Sub-Saharan Africa (Global Terrorism Index, 2023).

Coupled with these factors, Ekiti state contextual factors constrain the state’s resources. Southwest of Nigeria, the State is at 28% (Sasu, 2022), while Lagos State at 4%. In addition, the state has the highest fertility rate of 4.7 the highest dependency ratio of 0.8, and the highest life expectancy of 61.9% in southwestern Nigeria (National et al. Nigeria (2020). Therefore, these unique contextual factors seem to put challenging resource dynamics into the State leading to increase in prevalence of PTSD. This has led to decreased life satisfaction among senior citizens, and research has consistently associated life satisfaction with PTSD (Basharpoor & Eyni, 2021; Mccanlies et al., 2014). To date, we know little about the relationship between LS and PTSD among senior citizens in Ekiti State.

Importantly, inadequate perceived social support has emerged as a pivotal risk factor for the onset and persistence of PTSD symptoms within the general population [Brewin, Andrews, & Valentine, 2000; Ehlers & Clark, 2000; Holeva, Tarrier, & Wells, 2001; Ozer, Lipsey, & Weiss, 2003; Robinaugh et al., 2011]. Poverty has been identified as one of the factors leading to inadequate perceived social support (Weyers et al., 2008). Recent reports from the World Bank (April 2023) indicate that approximately 13 million Nigerians are living below the poverty line. According to the National Bureau of Statistics (2022), a substantial 63% of Nigerians (133 million people) experience multidimensional poverty, encompassing monetary poverty, limited access to education, and essential infrastructure services, with the projection that the rate of growth of senior citizens in Nigeria will increase from 6% in 2030 to 9.8 in 2050 (Togonu-Bicke resteth and Akinyemi2014).

Currently, more epidemiological researchers of PTSD have been accused of excluding older adults or not including sufficient numbers in their studies (Glick et al., 2018), and other scholars argue that the senior citizens often receive insufficient attention from policymakers, possibly due to their comparatively lesser contributions (Zelst et al., 2017). To the best of our knowledge, there is a dearth of literature on PTSD among senior citizens older adults in Nigeria.

Furthermore, recent estimates predict that by 2025, the senior citizens of Nigeria will increase to 11.5 million and 16.5 million in seven years (2030) and 25.5 million by 2050. While this demographic sector of the population is growing, research is seemingly silent on the level of PTSD in Ekiti state as many studies from Europe and the West have indicated that PTSD is common among senior citizens (Zelst et al., 2017; Lapp et al., 2007; Owens et al., 2005; Mainous et al., 2005).

Nigeria has no legislation for the aged population, however, Nigeria has the largest population of older people than any African nation and is the 19th largest population of senior citizens globally. It is projected to increase in the next three decades to approximately 33.3 million people and become the 11th largest in 2050 (Wan, 2022.)

Moreover, as the population of older adults grows, they are considered to face challenges related to their mental health conditions, especially life satisfaction (LS). It has been reported that senior citizens in Ekiti State are experiencing a decrease in life satisfaction (Arogundade and Adebayo (2011; Aina et al., 2023), with growth in older adults and decrease in mental health facilities (Chima et al., 2022). Indeed, this is the opposite of what is happening in Europe or Western society. Currently, the Western world and Europe are experiencing stable and peak mental health conditions among senior citizens, such as increased well-being at older ages (Carstensen et al., 2003; Carstensen et al., 1999; Fung et al., 2008), with fewer experiences of negative emotion (Gross et al., 1997) fewer stressors in daily life (Stawski et al., 2008), fewer negative affect (Charles et al., 2001), and fewer harmful effects than positive stimuli (Charles et al., 2003; Mather & Knight, 2005).

Moreover, Nigeria is lagging behind in term of the availability of mental health facilities and professionals. Currently, only 8 Federal and 15 State mental health services in Nigeria cater primarily to rural dwellers (Chimam et al., 2022; Ojagbemi, Bello, & Gureje,2018). The limited mental health workforce includes approximately five psychiatric nurses per 100,000 Nigerians, alongside a scarcity of clinical psychologists and social workers (Ojagbemi, Bello, & Gureje, 2018; Olugbile, et al, 2008; Ryan, 2020; UN, 2018; Nigeria (5 September 2018).

According to Consumer News and Business Channel Africa (CNBC, 2020), Nigeria lacks research on mental health demographics, and this lack of research culminate in further research that in which 20 to 30% of Nigerians suffer from mental health disorders while approximately 50 million Nigerians or one in four Nigerians are experiencing mental illness (The Guardian 4 Dec. 2018; API and EpiAFRIC Jan. 2020; Al Jazeera 2 Oct. 2019). Indeed, the Nigerian government has embarked on implementing the African Union (AU) recommendation for budget allocation of 2001 15% (THISDAYLIVE. 2019-09-05), which was implemented by the Nigerian government. However, Nigeria has maintained single digits with budget allocations moving from ascending order to descending 5.95% in 2012, 4.4% in 2014, 5.5% in 2015, 4.23% in 2016, 4.16% in 2017,and 3.9% in 2018. The population is increasing with the worst allocations of budgets to the State and Local Government (29 and 8% respectively) (Vanguard News. 2021-08-26). Despite the low budgetary allocation of health sectors, Nigeria still does not have a defined or definite budget allocation for mental health (THISDAYLIVE. 2019-09-05).

Coupled with these phenomena, the contextual factors of the Ekiti State are certainly depleting resources. The state is the poorest in the southwest region of Nigeria, with 28% of the country (Sasu, 2022) the highest fertility rate is 4.37, the highest dependency ratio is 0.8, and the highest life expectancy is 61.9% (National Population Commission of Nigeria (NPCP, 2020). These unique contextual factors put challenging resource dynamics on the State; therefore, the need to employ life management strategies to address and alleviate mental health challenges becomes imperative.

Selection, optimization, and compensation (SOC) is an age-sensitive life management intervention model of mental health that recognizes individual’s challenges as they age (Freund & Baltes, 1999) Freund and Baltes (200a) highlighted the critical role of SOC management strategies in mitigating mental health challenges caused by changes in various life domains during old age. Some researchers propose that SOC might be a trait variable influencing the risk of depression and relapse (Zuroff et al., 1999), however, to the best of our knowledge, the use of SOC as a therapeutic life management strategy is rare in Nigeria. As a result, conclusive evidence regarding the effectiveness of SOC as a therapy for mental health challenges remains elusive. Remarkably, this study is the first to delve into this uncharted territory, contributing to understanding how SOC strategies, which play a critical role in adaptive aging, intersect with the complex dynamics of PTSD. By addressing this novel research gap, this study seeks to illuminate the mechanisms underlying the interaction between SOC and PTSD, ultimately offering insights that hold potential implications for both theory and practice, guiding policymakers and social scientists in implementing intervention techniques that alleviate the effects of aging among older individuals.

Furthermore, perceived social support (PSS) has been recognized as a crucial resource and emotional regulatory mechanism across different age groups (Philipp, 2012). Research has highlighted the pivotal roles of life satisfaction (LS) and perceived social support (PSS) in influencing the physical and mental well-being of older adults (Borg et al., 2006; Harandi et al., 2017). The significance of SOC strategies is also stressed by their associations with various psychological factors, including personality traits and emotional and mental health indicators such as life satisfaction, perceived social support, and PTSD. Amazingly, there is a gap in the literature concerning the relationships between SOC strategies, PTSD, life satisfaction and perceived social support despite the SOC model providing valuable insights into PTSD.

Moreover, the inconclusiveness surrounding the impact of SOC strategies on mental health (Shirzadifard et al., 2020; Feund & Baltes, 1998) prompted us to propose a moderated mediation model. Specifically, we postulate that the associations between SOC and PTSD may be mediated by perceived social support and life satisfaction, with age groups serving as moderators. This approach aims to unveil the intricate interplay between these variables and discern the conditions under which they are linked, mainly focusing on the roles of indirect effects on PTSD.

This study aimed to address these knowledge gaps by investigating the connections between SOC strategies, PTSD symptoms, perceived social support, and life satisfaction among older persons in Ekiti State, Nigeria. By examining the moderating role of age groups, we seek to provide insights into the nuanced interactions among these variables and their implications for mental health outcomes. This investigation will not only advance the theoretical understanding of the relationships between SOC strategies, PTSD, and perceived social support, but also offer practical implications for interventions to promote mental well-being among older individuals. Additionally, the high prevalence of mental illness and the lack of dedicated budget allocation for mental health services underscore the urgent need for research and therapeutic interventions. The second objective this study was to examine the impact of age on life satisfaction, perceived social support, PTSD, and selection optimization compensation (SOC) among senior citizens. The study aimed to explore potential correlations, patterns, and trends in these variables across distinct age ranges (65-74, 75-84, and 85 years and older).

### GENERAL RESEARCH QUESTIONS

The objectives of this study, the following general research questions were formulated: What is the relationship between selection optimization compensation (SOC) strategies and posttraumatic stress disorder (PTSD) among the older adults in Ekiti State, Nigeria, considering the mediating role of perceived social support and life satisfaction and the moderating role of age groups? How do age-related differences influence the overall mental health and well-being of individuals?

### SPECIFIC RESEARCH QUESTIONS

Based on these general research questions, the following specific questions were raised:

1. To what extent do SOC strategies influence perceived social support among senior citizens in Ekiti state?
2. How do SOC strategies relate to life satisfaction among senior citizens in Ekiti State?
3. What is the association between SOC strategies and PTSD among older adult senior citizens in Ekiti State?
4. How do perceived social support impact PTSD among senior citizens in Ekiti State?
5. To what extent does life satisfaction influence PTSD among senior citizens in Ekiti State?
6. How do the age groups (young-old, old-old, and oldest-old) moderate the indirect relationship between SOC strategies and PTSD
7. How would the mediating roles of life satisfaction and perceived social support influence SOC and PTSD
8. How do age-related differences influence the overall mental health and well-being of individuals, considering variations in life satisfaction, perceived social support, PTSD, and SOC across different age groups?

On the basis of these research questions, the following hypotheses were proposed.

### HYPOTHESES

1. There was no significant relationship between SOC strategies and perceived social support among senior citizens in Ekiti State.
2. There was no significant relationship between SOC strategies and life satisfaction among senior citizens in Ekiti State.
3. There was no significant relationship between SOC strategies and PTSD among senior citizens in Ekiti State.
4. Perceived social support does not significantly influence PTSD among senior citizens in Ekiti State.
5. Life satisfaction does not significantly influence PTSD among senior citizens in Ekiti State.
6. Age groups (young-old, old-old oldest-old) will significantly moderate the relationship between SOC and PTSD among senior citizens in Ekiti State.
7. Life satisfaction and perceived social support will significantly mediate the relationship between SOC and PTSD.
8. There will be significant Age-related differences in life satisfaction, perceived social support, PTSD, and SOC across different age groups of the older people in Ekti state.

### The significance of the study

The significant of the current study lies in senior citizens’ direct impact on Ekiti State’s resource constraints. Within this context, this population is characterized by high life expectancy, a high fertility rate, and high dependency ratio which threaten individuals’ perceived support and life satisfaction. As a result, they are susceptible to mental health challenges, including PTSD. By addressing these critical issues, this research aims to improve the well-being of Ekiti’s senior citizens, offering practical solutions to alleviate their burdens and enhance their mental health and overall well-being. This pioneering study on SOC strategies in Nigeria, specifically in Ekiti State, is a groundbreaking exploration. This study offers a unique perspective on the vital role of SOC strategies among senior citizens within resource-depleted contexts, shedding light on their potential to mitigate the effects of mental health challenges in the State. By breaking new ground in this area of research, this study contributes to the well-being of Ekiti State’s senior citizens. This finding underscores the importance of SOC strategies in addressing mental health issues in resource-constrained settings.

This study holds significant importance for the people of Nigeria, particularly in Ekiti State, where individuals have become caregivers for their aging parents due to limited resource allocation and the absence of government policies for elderly individuals. By investigating the factors that lead to PTSD among senior citizens and employing SOC strategies such as psychotherapy, this research provides practical insights into how SOC as a protective factor can alleviate the accumulated stress of caregivers. Ultimately, the study directly benefits those who are responsible for caring for their aging parents by offering strategies to reduce their burden and enhance the well-being of senior citizens. This pioneering study, the first to investigate the association between SOC, PTSD and life satisfaction, holds paramount importance. This study bridges existing research gaps and yields practical implications for interventions and programs designed to enhance the psychological well-being of golden agers in Ekiti State. By potentially influencing healthcare and mental health support policies for older individuals, the study’s findings extend our understanding of how SOC moderates the effects of PTSD in this population. Operating in uncharted territory, this research informs policymakers and interventionists about the unique needs of senior citizens in Ekiti State, offering a context-specific perspective that enriches the literature and underscores the necessity of considering local factors when addressing challenges faced by older adults.

Additionally, the study will identify significant differences in mental health variables across age groups informing the development of tailored interventions, addressing the unique needs of different age cohorts and enhancing overall well-being. By revealing discernible patterns in these variables, the study will contribute to a nuanced understanding of age-related dynamics, enabling policymakers and healthcare providers to implement targeted support strategies for specific age ranges, thereby improving mental health outcomes. Moreso, this study holds significance in contributing to the understanding of the nuanced relationships between age and mental health among senior citizens. The findings may inform interventions tailored to specific age groups, facilitating the development of targeted strategies to enhance overall well-being in later stages of life. Additionally, the study provides valuable insights for policymakers, healthcare professionals, and researchers working in the field of geronpsychology, gerontology and mental health.

### Life satisfaction (LS) and Perceived Social Support (PSS) on PTSD

A germane explanatory mechanism of the relationship between SOC and PTSD can be derived from a study suggesting that senior citizens’ level of PTSD depends on life satisfaction and perceived social support (Basharpoor & Eyni, 2021). PSS is a cognitive construct that is embedded in personality characteristics. According to this present study, PSS are defined as the belief that family, friends, and significant others will be available to help, advise, or comfort when needed (Yarseah, et al.,2023). According to the stress buffering model, social support may reduce the psychological effect of a stressful event by attenuating the stress appraisal response (Cohen & Willis, 1985). After a stressful event, social support may ease cognitive and emotional processing, permitting victims to reevaluate the possibility of adapting to the phenomenon (Thoits, 1986; Williams & Joseph, 1999). Individuals may deem an event less stressful if they perceive that others will provide the resources needed to cope with it.

However, the concept of perceiver social support is inconsequential in Nigeria and has yet to receive the most needed scholarly approach. Many Nigerian scholars have not appreciated the conceptualization of received and perceived support, as most Nigerian scholars often mismatch social support and receive support and neglect must-needed PSS (Agbor, Samami, Uzochukwu, 2017; Onalu & Nwafor, 2021; Ebimgbo & Okoye, 2017; Osamor,2015; Etodike, Ezeh, Chukwure, 2017). Perceived social support is the information leading the individual to believe that he/she is a member of a social conveyor of mutual obligation (Cobb, 1976). Received social support provides emotional and instrumental support to individuals through close confidants or significant others (Schulz & Schwarzer, 2004). That is received social support is the quality and quantity of support given by the support providers. Many studies on perceived social support have shown that perceive social support is correlated with reduced stress and improved physical and mental health. On the other hand, research on received social support is inconclusive at best (Haber et al., 2007).

Previous studies have shown that, in general, PSS is correlated with improved physical and mental health (House et al., 1988; Lakey & Cronin, 2008; Uchino, 2009) and that PSS is more critical than received support in predicting adjustment to life stress (Wethington & Kessler, 1986). On the other hand, researchers have found that stressful situations, received social support is positively linked with negative affect (Peeters & Le Blanc, 2001; Yang & Carayon, 1995) and depression (Frese, 1999), and received social support and health are often negligible and inconsistent (Bolger & Amaral, (2007; Uchino, 2009), however, the association between PSS and health is consistently positive (Uchino, 2004, 2009; Uchino et al., 2011).

Indeed, PSS is an essential determinant for improving the LS of older people (Antonuocci et al., 1996; Lampinen et al., 2006; Seeman2000), and in recent years, PSS has been confirmed to be a significant factor influencing LS in a wide variety of population groups, such as older people (Wu et al., 2022). Perceived social support significantly predicts life satisfaction and adverse effects (Siedlecki et al., 2014; Shensa et al., 2020). However, the absence of perceived social support from family, friends, and significant others has been widely proven to decrease one’s life satisfaction (Yu, 2020; Chen, 20’7). Although studies have confirmed the impact of perceived social support on life satisfaction, the potential mechanisms underlying this correlation remain obscure due to its complexity (Song & Fan, 2013). In this atmosphere, little attention has been given to the relationship between perceived social support and life satisfaction among elderly people, especially in Ekiti State.

Furthermore, Neugarten, Havighurst & Tobin (1961) define LS as older person with a more positive self-concept that reflects the appropriateness of past goals and outcomes based on current living conditions. LS is closely related to a wide spectrum of human behavioral, psychological, social, interpersonal, and intrapersonal outcomes, such as buffering stress and diminishing externalizing behaviors and problem internalization (Proctor et al., 2009). Moreover, many studies have indicated that individuals with higher levels of PSS are innocuous of negative mental stability effects, such as social pressure, anxiety, psychological distress, and depression, and thus experience grater levels of satisfaction in life (McDougall et al., 2016).

Additionally, PTSD can develop after a person becomes exposed to life-threatening events outside a range of normal human experiences and can markedly distress everyone (Diagnostic and Statistical. Manual of (Mental Health Disorder, 1987). Several studies have indicated that the most prevalent symptom of PTSD among senior citizens is hypervigilance (sleep disturbance, anger, trouble concentrating, fatigue, muscle tension, or pain) (Lapp et al., 2007; Owens et al., 2005; Mainous et al., 2005). Some researchers have argued that PTSD symptoms in senior citizens are more likely to be accompanied by somatic symptoms and, particularly, hyperarousal symptoms (Owens et al., 2005; Mainous et al., 2005). However, more recent findings suggest that as PTSD victims age, their avoidance and arousal symptoms persist or increase, whereas re-experiencing symptoms decrease (Louwers 2015).

However, studies examining the relationships between PSS scores and PTSD incidence are noticeably inconclusive, as there are limited studies that have correlated PTSD and PSS scores. PSS moderates stress and depression and is correlated with positive health behavior and a sense of security among senior citizens (Mirowsky, 2002). It has been stated that poor perceived social support is one of the most critical risk factors for the maintenance of PTSD, the development of PTSD symptoms (Brewin et al., 2000; Ehlers & Clark, 2000; Holeva et al., 2001; Ozer et al., 2003; Robinaugh et al., 2011) and a lack of life satisfaction (Weyers, 2008). To date, a recent report from the Work Bank (April 2023) forecasts that approximately 13 million Nigerians fall below the poverty level, and the National Bureau of Statistics (2022) states that 63% (133 million people) of Nigerians are multidimensionally poor (monetary poverty, education, and basic infrastructure services). In particular, Ekiti State in southwestern Nigeria is the poorest state with a poverty headcount rate of 28% compared to Lagos which is the richest state in Nigeria (Sasu, 2022). In addition, Ekiti State has the highest dependency ratio and the highest total fertility rate in Southwest Nigeria, 0.8% and 4.37%, respectively (National et al. Nigeria, NPCN, 2020).

These demographic factors make Ekiti State a resource-depleting State, which could lead to poor perceived social support and life satisfaction and enhance the exacerbation of PTSD among elderly people; however, no scholar has taken a advantage of these factors. On the other hand, several researchers have portrayed PSS as a relatively stable trait (Thomas, et al 2022; Levy 2019) that shares commonalities with personality dispositions (Allemand, Schafhuser & Martin, 2015). However, PSSs can also be considered recidivists over time and often depend on one’s living circumstances, one’s current mental illness, and one’s exposure to positive or negative life events (Thoresen et al, 2019; Kaniasty K, Norris, 2008). Severe stressful event exposure could have a significant effect on perceived social support (Adler &, Castro 2013). Furthermore, individuals coping with severe stressful event exposure might have difficulties engaging in trusting relationships (Thoresen et al, 2019; Kaniasty K, Norris 2008). However, there are different results on whether PSSs mitigate, elevate, or remain constant during stressful events (Thoresen et al, 2019; Kaniasty K, Norris, 2008; Keane et al, 1985; Birkeland et al, 2016). In addition, there is evidence that grater psychological symptoms lead to lower PSS (Kaniasty K, Norris, 2008; Wang et al 2021; Lui, Glynn& Shetty 2009), which is often due to the avoidance of PTSD clusters by senior citizens (McFarlane, 2000). Psychological symptoms, such as PTSD and depressive symptoms, can affect people’s way of thinking and negatively influence their sense of social support [Couette, et al, 2020; Bishop et al, 2022). Another possible common cause of lower PSS scores could be a history of trauma which can be emotionally distressing. Exposure to traumatic events often contributes to psychopathology (Breslau, et al 1999) and is also associated with lower perceived social support (Thoresen, 2019).

Indeed, Kaniasty and Norris (2008) examined individuals exposed to natural disasters and showed that more PTSD symptoms were associated with less PSS 18 and 24 months post-trauma-trauma, supporting the notion of social selection, which posits that PTSD can influence social support. In a previous study, PSS was shown to play mediating roles between challenging life events and psychological distress, such as anxiety, depression, and behavioral distress (Russell & Cutrona, 1991). Research has also shown that higher levels of perceived social support are associated with more positive PTSD recovery outcomes (King et al., 1999).

However, social causation or social deterioration theory has also been used to explain why PSS has a negative impact on PTSD. The social erosion or social deterioration theory states that PTSD erodes because people with PTSD have an increased tendency to believe that others are dangerous and unsafe. (Clapp& Gayle-Beck 2009), embracing the idea that social support is diminished by poor mental health (Wagner et al., 2016; Woodward et al., 2018). These perceptions increase the likelihood of individuals with PTSD having difficulty establishing trust and, as a result, isolating themselves (Erbes et al., 2012; Wagner et al., 2016). From this perspective researchers have suggested that symptoms of PTSD, such as insecurity, skepticism, social avoidance, and social isolation, can lead to rejection and less PSS and LS from others (Mossakowski, 2014). Additionally, in another study of adults exposed to physical assault in Norway, higher PSS levels protected against the development of PTSD symptoms, and diminished PSS increased the risk of developing PTSD symptoms (Johansen et al., 2020). Many scholars have built from this perspective to suggest that due to the avoidance and re-experiencing of symptoms of PTSD, PSS could have a negligible effect on PTSD (Freedman et al., 2015; Wagner et al., 2016; Woodward et al., 2018).

In addition, PTSD symptoms have crippling effects on social relationships through expanded experiences of unpleasurable social interactions as well as through the erosion of social resources in close relationships across time (Freedman et al., 2015; Wagner et al., 2016; Woodward et al., 2018). A cross-sectional study of 24 individuals recruited to assess the relationship between PSS and the PTSD symptom clusters of the DSM-5 and ICD-11 suggested that individuals with a presentation of complex PTSD exhibited lower levels of PSS than individuals not presenting with CPTSD and that a lower PSS was significantly correlated with the likelihood of presenting with CPTSD (Simon et al., 2019). To resolve this obscurity, we reconsider the relationship between these two variables among senior citizens in the present study. Further knowledge of the relationships between PSS and PTSD may have significant implications for the development of social policy that enhances the psychosocial well-being of senior citizens in Ekiti State, Nigeria.

Given the nature of PTSD among senior citizens as avoidance, withdrawal, and decreased re-experiencing, and the nature of older adults reporting fewer experiences of negative emotion (Gross et al., 1997) and fewer stressors in daily life (Stawski et al., 2008) than younger adults older adults also experience decreases in negative affect (Charles et al., 2001) and recall fewer negative than positive stimuli compared to younger adults (Charles et al., 2003; Mather & Knight, 2005). We speculate that LS and PSS could be explanatory variables for SOC and PTSD. LS impats an individual’s sense of well-being and quality of life. It is the fit between the desired goals in life and actual life outcomes (Diener et al., 1985; Headley et al., 1993; George, 1991). Researchers have demonstrated that people who are dissatisfied with life are more likely to suffer psychiatric morbidity, including depression and anxiety which can lead to an increased risk of suicide and increased mortality (Deeg et al., 1989; Feller et al., 2013). A decrease in LS correlated with comorbidities and PTSD (Samaranayake & Fernando 2011; Qureshi et al., 2009).

Although there are many studies on LS among senior citizens in Nigeria and in Ekiti State in particular (Odetola et al., 2020; Bola, ND; Arogundade & Adebayo, 2011; Adebayo & Arogundarde, 2011), none of these studies have examined the influence of LS on PTSD incidence. In a study of LS among senior citizens living in geriatric homes in Lagos, Odetola et al. (2020) demonstrated that most senior citizens were generally satisfied with their lives. However, loneliness, childlessness, and illness were the main causes of life dissatisfaction. In Ekiti State, in particular, a study on the roles of life satisfaction judgment among educated adults, Arogundade and Adebayo (2011), demonstrated that L S decreases as one progresses in age and that family relations are the single predictor of life satisfaction (Adebayo and Arogundarde (2011). A more recent cross-sectional study of hospitalized patients was designed to determine the association between social support and life satisfaction among older adults in Ekiti State. The results showed that 73.8% of the senior citizens were dissatisfied with their lives, and a factor associated with their dissatisfaction was the lack of government support wherein 81% of the senior citizens depend on their relationships for support (Aina et al., 2023). The findings from these studies indicate that there are low levels of life satisfaction in Ekiti State, and the factors contributing to life dissatisfaction are loneliness, childlessness, and lack of government support. This shows that senior citizens in Nigeria might have some level of mental health challenges such as PTSD, but to date, no study has investigated this this area of life.

While these studies in Nigeria do not link LS with PTSD, some limited studies elsewhere have associated PTSD with LS. A study of community samples assessing the effect of LS on PTSD showed that people with average severe PTSD in the community were likely to struggle with lower levels of life satisfaction than people with other mental health conditions or those without any diagnoses (Karatzias et al., 2013). Furthermore, in a cross-sectional study among police officers following Hurricane Katrina in New Orleans on the association between resilience, gratitude, life satisfaction, and PTSD, the results indicated that positive factors such as resilience, satisfaction with life, and gratitude may help mitigate symptoms of PTSD (McCanlies et al., 2014). This is because older adults tend to report greater life satisfaction and positive affect, as well as less negative affect than younger adults (Carstensen et al., 2000; Mroczek & Kolarz, 1998; Mroczek & Spiro, 2005; Shook, 2010). Aditionally, in the general population, one study indicated that people with moderate or severe PTSD are likely to experience low levels of LS (Karatzias et al., 2013). Among military veterans in the United States, meaning in life was correlated with lower levels of PTSD symptoms (Owens et al., 2009). Another study showed that individuals with low life satisfaction may experience worse PTSD symptoms than may those who report high life satisfaction (McCanlies et al., 2014), and those with PTSD are likely to experience low levels of LS (Karatzias et al., 2013). While these studies are correlational in nature and cannot tell whether PTSD causes low LS or whether LS causes PTSD, some studies have reported that perceived social support and life satisfaction play important roles in the physical and mental health of senior citizens (Borg et al., 2006, Harandi, el al., 2017). To resolve this ambiguity, we therefore postulate that PSS and LS are explanatory variables for SOC and PTSD in the present study.

### SOC, PSS, on PTSD

Selection optimization and compensation (SOC) is a model developed by Baltes and Baltes (1990) as a process that older adults can use to cope with life stressors that are connected with aging (Freund & Baltes,200a). The meta-theory stipulates that three life management processes lead to successful aging. These include selection (a categorical choice for gain), optimization (method for maximizing gain), and compensation (minimizing losses) [Baltes and Baltes, 1990]. The theory is a life management model for adjusting to physical and behavioral changes (Fruend & Baltes,(200a). Individuals endeavor to exert energy to preserve physical and emotional stability while seeking measures of pay-off for losses and to become more proficient in activities they know how to do best (Baltes et al., 2006). Adult life experiences are characterized by contrition in the direction of less resources gained and more resources lost, meaning that aging measures both growth and decline (Baltes, 1987; Baltes et al., 1996). Baltes et al., (2006) further added that aging is mainly gain for young people and loss for senior citizens since adult development is a shift in the direction of less resource gain and more resource loss (Baltes, 1987). Similarly, Blanchard-Fields et al., (1997) stated that SOC could be a model of coping (Lazarus & Folkman, 1984), problem solving (Blanchard Fields et al., 1997), self-development, and goal adjustment (Brandtstädter & Rothermund, 2002).

However, we perceived SOC awareness and usage as emotional constraints that could exacerbate PTSD in elderly people. We therefore assumed that SOC has a significant relationship with PTSD through the PSS. This assumption is supported by a study that suggested that greater adherence to SOC strategies was correlated with higher levels of physical health and stress among older adults with multimorbidity, in which greater disability led to greater use of SOC strategies (Yuen & Vogtle, 2016). To our knowledge, there is a dearth of literature on the relationship between SOC and PTSD, and only limited studies have examined PSS and SOC. Moreover, proactive strategies to promote SOC have been animated as part of optimal psychotherapy in the case of poststroke depression (Aspinwall & Taylor, 1997; Broomfield et al., 2010; Ouwehand et al., 2007). Currently, the SOC model has been extended to social aging due to demands bounced by social relationships (Weiland, 2011), which might reasonably have some connotations for the correlations between perceived social support and the development of PTSD, especially in old age (Rohr & Lang, 2009). Moreover, differing viewpoints also exist on this concept (Shirzadifard et al., 2020). For instance, Shirzadifard et al. (2020) reported that implementing SOC strategies leads to significant variability in life satisfaction and correlated with increased depressive symptoms (Feund & Baltes, 1998).

Moreover, some studies suggest that SOC is mood-dependent and reduced by age and that advanced age and resource deficits may restrict SOC strategies (Baltes & Lang, 1997). Freund & Baltes (1998; Weiland, Dammermann, Stoppe.2011). In addition, SOC has been regarded as a trait variable that can increase or decrease the risk of depression and/or, at times, deterioration (Zuro et al., 1999), although this claim has been disputed by other scholars who perceive SOC as a therapeutic variable (Teshale & Lachman, 2016; Samantary & Kay, 2021).

On the other hand, the use of SOC strategies significantly exacerbates depressive symptoms (Gestsgottir et al., 2010), depression, drinking, and smoking (Gestsgottir et al., 2009). Of course, more recent studies have proven otherwise (Welland et al., 2011); therefore, appears that SOC could be characterized as a state and trait variable that has dysfunctional behavior in both short and long time frames, which could maximize the development of PTSD among senior citizens.

Indeed, previous studies have failed to focus on the association between SOC and PTSD through the PSS. To close this knowledge gap, we stipulate that the relationships between SOC and PTSD could be explained through the mechanism of PSS and LS. While previous studies have not shown that content is associated with PTSD through LS or PSS, we propose that the relationship between SOC content and PTSD could be aligned associated with depression. Studies have shown that there are some overlaps in depression and PTSD symptoms as there are some common symptoms of PTSD and depression such as insomnia, decreased concentration, avoidance and withdrawal, lack of interest distance from others, and anhedonia (Grubaugh et al., 2007; Brady et al., 2000), additionally, the comorbidity rate is 54.41–54.72% after PTSD and depression (Brady et al., 2000). Again, that depression elevates the risk of developing PTSD after trauma exposure (Koenen et al., 2002), and PTSD increases the risk of developing first-onset depression following a trauma (Breslau et al., 2000; Kessler et al., 1995). These correlations indicate that the SOC could influence PTSD among elderly people, but to this extent, no study has examined this relationship.

Given the nature of greater SOC, such as resource loss, social convoy selection, learning to adjust to a new environment, obtaining help from significant others, and modifying relationships, it seems plausible to contribute to the literature that PSS could explain the relationship between SOC and PTSD for the first time. PSS is a primary mutual or social resource essential for managing or coping with stress and has been associated with psychological wellbeing during times of stress (Norris & Kaniasty, 1996). Compared with received social support, self-compassion has been firmly connected to people’s ability to adapt, regulate, adjust, and cope with stress (Norris & Kaniasty, 1996). Specially, a study has shown that after leaving the war zone, veterans who had a more positive view of being supported after coming from the battlefield had less intense PTSD symptoms and better mental health and quality of life than did those who did not(Jukic, 2020).

A meta-analysis indicated that not having a network of meaningful relationships in life is more predictive of mortality that could result in an antisocial lifestyle, such as smoking or physical activity (Holt-Lunstad & Smith, 2012). More significantly, cross-sectional research has shown that social support and PTSD are negatively related; individuals who survive a disaster but with more social support have less severe PTSD symptoms, and those with more severe PTSD symptoms report less social support (Brewin, et al., 2000; Gray et al., 2006; Ozer et al., 2003; Wagner et al., 2006). Conversely, perceived social support plays a more critical role in the emotional and psychological wellbeing and positive outcomes of some traumatic events than does large social networks or the type of support offered (Kaniasty Norris,(2008). Research has suggested that perceived social support mediates the association between unbearable disasters and psychological problems, such as stress, psychological distress, and behavioral distress (Russell & Cutrona, 1991). However, it is generally known that poor perceived social support is one of the most critical vulnerabilities to the development and maintenance of PTSD symptoms (Brewin et al., 2000; Ehlers & Clark, 2000; Holeva et al., 2001; Ozer et al., 2003).

Although there is a research gap between SOC and PSS, the conservation of resources (COR) theory of Hobfol (1998) can play a critical role in bridging the gap between perceived social support and SOC by providing a comprehensive framework that explains the underlying mechanisms and interactions between these concepts. COR theory is particularly essential for understanding how people manage and accumulate primary, secondary, and tertiary resources to cope with stressors and challenges. (The COR) theory is fascinating in the present study because it goes beyond merely correlating resources to performance and other mental health issues among elderly people. COR theory (Hobfoll,1998; 1989) emerged from resource and psychosocial theories of stress and human motivation. In the context of the relationship between perceived social support and SOC strategies, COR theory offers insights into how individuals strategically invest their resources to maintain and enhance their wellbeing (Hobfoll, 1998). Perceived social support can be conceptualized as a valuable subjective psychological resource that individuals can draw upon when facing stressors or adversity. By perceiving support from their social network, elderly people can experience emotional comfort, validation, and assistance in coping with difficulties.

On the other hand, SOC strategies involve actively managing resources to adapt to changing circumstances and demands (Freund and Baltes, 2002). These strategies include selecting the most relevant goals and relationships, optimizing the use of available resources, and compensating for potential deficits. This theory relates to the COR theory that individuals strive to acquire, retain, and protect resources, as losses and threats to resources can lead to adverse outcomes such as stress and psychological distress (Hobfoll, 1988; 1998). Considering the interplay between perceived social support and SOC strategies, COR theory suggests that individuals who perceive grater social support may have a more incredible resource pool from which to draw (Hobfoll, 1988). This enhanced resource pool could facilitate more effective SOC strategies, as individuals with a strong support network might experience lower stress levels and have more energy and capacity to engage in adaptive coping behaviors.

The perceived availability of social support may act as a buffer against resource loss, reducing the impact of stressors on an individual’s wellbeing. Individuals facing challenges can rely on their perceived social support to alleviate stress and minimize the need for extensive resource compensation (Cohen & Wills 1985). This aligns with the core principle of COR theory, which emphasizes the importance of resource preservation. However, it has been demonstrated that SOC is a psychological and behavioral management model for adaptating to change (Grove et al., 2009) therefore, we speculated that SOC will significantly influence PSS to close this knowledge gap. SOC strategies involve various techniques and mechanisms for helping people deal with the myriad of changes that occur throughout their lifespan (Baltes & Baltes, 1990). The model can explain how loss-based selection in response to the loss of previously available resources and redirecting energy to account for those losses can be psychologically and physically distressing (Freund et al., 2009).

It is argued that optimization and compensation are effortful and can often become more stressful and demanding with age, possibly exceeding the individual, social, and technical availability (Freund et al., 2009). In this case, PSS could mediate SOC since SOC compensation involves helping others, developing new skills, and devoting more energy and time to compensate for the situation (Baltes & Christensen, 2003). Research has indicated that perceived social support is significantly associated with personality traits (Pierce et al., 1997; Swickert, 2009), and studies have indicated that cognitive and personality variables are the strongest and most stable variables that explain SOC management strategies (SOCs) (Baltes & Freund, 2002). Again, Baltes (2002) demonstrated that the SOC measure predicts uniqueness in intellectual functioning and thinking styles (Sternberg, 1994) and adaptive coping styles, such as flexible goal adjustment and tenacious goal pursuit (Brandsta & Renner, 1990). At a behavioral level, Lang, Rieckmann, and Baltes (2002) found that, compared with resource-poor older adults, people who were rich in social–personality (those who enjoy social relationships, seek help, and physical closeness) resources exhibited everyday functional patterns that pointed to greatedd use of SOC strategies. The SOC model complements models of coping (Lazarus & Folkman, 1984), problem solving (Blanchard-Fields et al., 1997), LS development, and goal adjustment (Brandtstädter, 1999; Brandtstädter & Rothermund, 2002). A recent study of this topic involved examining how SOC influences PTSD through PSS. Interestingly, no study has focused on SOC and PTSD through the PASS. Given the prevalence of PTSD among elderly people, this knowledge gap could be closed. SOC could be considered a trait variable that could increase or decrease the risk of PTSD (Zuroff et al., 1999).

### SOC, LS, ON PTSD

There are two theories of SOC. The first theory states that as one grows older, he/she becomes better at using SOC strategies due to the agglomeration of life experiences, while the second postulates that the biological and physical constraints associated with aging imply a loss of resources that may limit the use of SOC strategies (Baltes & Baltes, 1990). This second theory depicts how resource depletion could enhance depression, encompassing an older person, and lead to PTSD. Consequently, old age becomes SOC-related, and behaviors might decline (Freund& Baltes, 2002). The SOC model contends that one’s resources (mental, physical, and social) are limited, and as one ages, one tends to stretch across many domains; this decline in physical activity and behaviour could lead to psychological health problems such as PTSD that could be explained by life satisfaction. This thought is informed by the finding that SOC life-management strategies could become more complex with reduced resources (Jopp & Smith, 2006). However, studies on the effect of SOC through the LS on PTSD has been lacking in the literature. LS is a person’s global assessment of subjective quality of life or evaluative wellbeing component (Diener et al., 1985). Life satisfaction is considered one of the predictors of successful biopsychological adaptation in later life when an individual is often confronted with challenges related to loss of resources such as losses in health, physical and cognitive functioning, social convoy, and productive activities (Allerhand et al., 2014; Berg et al., 2007; Smith & Ryan, 2016; Schaie, 2016). Aging related losses are not accompanied by reduced life satisfaction in many people, a phenomenon known as the paradox of wellbeing. Despite natural age-related declines in health, income, and physical or cognitive abilities, senior citizens may remain satisfied because they make an accommodative shift that aligns their aims and goals with what is achievable (Brandstadter & Greve, 1994; Krauss-Whitbourne & Sneed, 2002). Older people’s ability to cultivate life satisfaction later life (Mroczek & Kolarz, 1998) is believed to be due to their improved ability to engage and proactively regulate emotions (Lawton et al., 1992).

In the same vein, socioemotional selectivity theory states that the life experiences older people have had over the years can successfully control and avoid potentially negative experiences by selecting social partners such as family and friends who provide positive emotional feedback and support (Carstensen, 1995; Carstensen et al., 1999). It is suggested that older adults, in general, often report higher levels of life satisfaction than younger adults (George, 2010). Furthermore, older adults tend to have higher levels of engagement with positive emotions than younger adults do, meaning that older adults pay more attention to and remember positive stimuli better than their younger counterparts (Martins et al., 2018). In general, Inglehart’s (1990) analysis of the Eurobarometer (surveys from 1980 to 1986) and World Values Survey suggested that people aged 65 years and older have higher levels of life satisfaction and happiness than younger age groups only once people differences in objective circumstances in income, education, and marital status were controlled for.

However, most of these studies were performed in developed countries with high GDPs, good healthcare policies, high income levels, and high life expectancies but not in developing nations or Africa. For example, Lucas and Gohm (2000) showed that age was positively related to life satisfaction in some countries but negatively related to life satisfaction in others. Happiness as a measure of subjective-well-being is greater in more economically developed countries than in less economically developed countries, as indicated by higher GDP per capita (Diener et al., 1995).

Research by Deaton (2008) using the Gallup World Poll revealed that life satisfaction declined linearly with age in most countries. However, the decline was most prominent for countries with lower GDP and the smallest for countries with mid-level GDP. In addition, social demographic factors such as marital status, income, education, number of children or wives, age, and religion have also been considered determinant factors of LS in Africa, especially in Nigeria (Adebayo & Arogundarde, 2011). Many studies have shown that being married, employed, religious, have better health, have higher income, receive more social support, and have higher education can each be positively related to aspects of wellbeing (Diener, 2000; Diener et al., 1999; Ellison, 1991; Kunzmann et al., 2000). In addition, in Nigeria, having children and being educated are associated with LS (Adebayo & Arogundarde, 2011).

Concerning at the complete idea of LS from the SOC’s viewpoint, a theoretical framework has been proposed that defines life satisfaction (LS) as an indicator of psychological adaptation to aging, where SOC is considered a psychological strategy for the management of resources (Lučanin, eta la, 2020). According to this view, older adults increase the positive effects of gains and decrease the adverse effects related to losses through selective investment in achievable, optimal goals thus, they compensate for their losses and limitations (Lučanin et al., 2020). By maintaining growth-related goals, older adults enhance their life satisfaction rather than focus on losses (Lučanin et al., 2020). Studies have established that greater SOC use at greater ages leads to greater well-being (Chou & Chi, 2001, 2002; Jopp & Smith, 2006). For example, Jopp and Smith (2006) proved that SOC strategies, used individually or in combination, counteract the adverse effects of low demographic (years of formal education), cognitive (perceptual speed), health (balance), and social resources (number of social partners) on wellbeing and satisfaction with age. More recently, SOC-based interventions have been found to have potential therapeutic effects on improving depression severity and the well-being of senior citizens in clinical settings (Samantaray & Kay, 2021). On this note, there is growing research evidence postulating that life satisfaction is a protective mechanism for the maintenance of health, reduces vulnerability to life-threatening illness, and promotes long life by way of positive emotions, which promote a more active lifestyle and motivation toward self-care (Carver et al., 2010).

SOC suggests that life satisfaction can be achieved by anyone who sets goals through effortful behaviors. Indeed, several studies have established that SOC is related to mental health, although the results are ambiguous (Jopp & Smith, 2006; Freund, 2008;Wiese et al., 2000; Wiese & Freund, 2000). A study examining the relationship between daily use of SOC and age groups showed that on days older people used more selection, optimization, and compensation strategies than did individuals with a week average, and individuals reported greater happiness. In particular, older and middle-aged adults showed a positive association between daily SOC usage and happiness (Jopp & Smith, 2006). A review by Freund (2008) suggested that using SOC could help individuals use resources effectively, thereby improving life satisfaction. Moreover, Jopp & Smith (2006) conducted a study among young and old people to assess the effect of resources and life management on life satisfaction. The results indicate that, generally, the SOC was significantly related to resources and aging satisfaction. When examining the associations between young-old and old-old individuals separately, substantial correlations were maintained between resources and aging satisfaction.

However, the relationships between SOC strategies and resources were weaker in old-old individuals than in young-old individuals. Moreover, SOC strategies were correlated with aging satisfaction only in the young-old individuals but not in old-old individuals (Jopp & Smith, 2006). The concurrent effects of resources and SOC usage, which correlate with well-being indicators in younger adults (Wiese et al, 200) and could benefit individuals across adulthood, should be examined (Wiese & Freund, 2000). In another study on improving depression and well-being, the results suggested that SOC-based psychotherapy had significant therapeutic effects on reducing depression severity and increasing the well-being of senior citizens individuals (Wiese & Freund, 2000).

On the other hand, PTSD has been associated with delay-on-set among older persons (Desmarais et al., 2020; Van der Wal, 2020; McCanlies et al., 2014) and other psychiatric symptoms (Deeg et al., 1989; Feller et al., 2013; Samaranayake, & Fernando, 2011) that might lead to low life satisfaction. Although there is an association between LF and PTSD (Karatzias et al, 2013; Owens et al., 2009), people who are dissatisfied with life are more likely to suffer psychiatric morbidity, including psychological distress are at an increased risk of suicide and have higher levels of mortality (Deeg et al., 1989; Feller et al., 2013; Samaranayake, & Fernando, (2011) which are comorbid with PTSD (Qureshi et al., 2009). Addditionally, one study indicated that people with moderate or severe PTSD are likely to experience low levels of LF (Karatzias et al., 2013), and meaning in life was associated with decreased PTSD symptoms (Owens et al., 2009). These findings imply for the first time that LF could be a reasonable explanatory variable for the association between SOC and PTSD.

Although no study has linked SOC to PTSD through LF, we intend to close this knowledge gap, and the study shows that a significant decrease in SOC competencies is associated with depressive symptoms (Weiland, 2011). On the other hand, studies have shown that sticking too much to the SOC procedures is associated with psychological distress, although it appears that those who have physical and emotional challenges are allergic to SOC usage, which in turn could exacerbate distress (Yuen & Vogtle, 2016; Lang et al.’ (2002). Yuen Vogtle (2016) explained that clinging to the SOC method was associated with greater physical health stressors. In a similar study among older adults with multimorbidity, a more significant disability led to greater use of SOC strategies (Yuen & Vogtle, 2016). They found that SOC strategies were unrelated to women’s loneliness or depressive symptoms. In a longitudinal study, Lang, Rieckmann, and Baltes (2002) reported that resource-rich participants were more likely to use SOC adaptive strategies than resource-poor participants were.

### Age groups as moderators of SOC and PTSD

Gorman (2000) posits that aging is a biological reality and process that no human can control. According to Shock (2018), when aging occurs in humans, physiological development is usually accompanied by psychological and behavioral changes and other changes involving social and economic factors. In their quest to find solutions to some of these challenges imposed by aging, scholars from the beginning of the mid-1980s developed considerable models that proposed that aging individuals can boost their health (Hartman-Stein & Potkanowiez, 2003; Haveman-Nies et al., 2003; Lang et al., 2005) while avoiding decreases and losses by taking responsibility for their health (Moody, 2001) and adapting their health behaviors (Holstein & Minkler, 2003).

Studies focusing on the use of the SOC procedure in very old adults could help researchers investigate the moderating effects of age groups on the association between SOC and PTSD. Age stratification and age groups (young-old, old-old, oldest-old) led to considerable content among scholars, policymakers, and social gerontologists. To this extent, there has yet to be a consensus among educators as to what constitutes age groups. Furthermore, many scholars have said that such conceptualization connotes ageism and should be disregarded (Kydd, 2020). Nevertheless, despite these contentions and misunderstandings, age stratifications have been adopted in policy documents, and today, the plurality of aging studies and policy documents use age bands to define an “old” population (Baars, 2007). The most common approaches in the aging literature include 65-74 (year (young old), 75-84 years (old-old), and 85 plus (oldest old) Neugarten (1974). Considering these factors, our reason for adopting this categorization in the present study is that the Age Harmonization Act of 2012 and 2022 of Nigeria put the retirement ages of judicial officers and professors of tertiary institutions at 70 years each and 65 years, respectively, for secondary and primary schoolteachers and judicial staff.

People aged 65-74 years, both young and old, are among the growing senior citizens in the Ekiti state and Nigeria. According to the National Population Commission (2003), young people constitute two-thirds (63%) of senior citizens nationwide, and in Ekiti State, this demographic sector comprises approximately 54.7%. Again, 35.9% of the elderly populations are aged 75-84 years, and this demographic population is also on the rise (NPS, 2003). Similarly, the oldest individuals (85+) in the state are few but increase in number. Recent data show that the oldest old people constitute 19.8% of the total population in Nigeria (National et al., 2001).

Some impairment associated with advanced age and resource deficits may limit the use of SOC strategies (Baltes & Lang, 1997). Freund and Baltes (1998), for example, demonstrated a negative age association for the use of optimization and compensation strategies in a sample of old and very old adults (73–103 years), with SOC usage decreasing with age (Feund & Baltes, 1998). In their landmark work, Feund and Baltes (1998) reported that the use of SOC strategies decreases with age as the execution of these strategies also requires resources. However, others believe that SOC usage has no limit across age groups. For example, Baltes (1998) reported that SOC behaviors occur from early to late adulthood, with middle-aged adults reporting the highest frequency of SOC behaviors. The developmental peak of SOC occurs in middle adulthood, when the desire to use most SOC strategies is greater than that in early and late adulthood. The use of elective selection increases steadily with age, from early to late adulthood, and only those in middle adulthood show the highest consistency for SOC components (Freund & Baltes, 2002).

In general, SOC seems to be particularly helpful for older individuals with fewer available resources (Jopp & Smith, 2006; Lang et al., 2002) and for younger adults (Young et al., 2007); however, the use of SOC strategies is related to positive outcomes well into ancient age as was shown in a sample spanning the age range of 72 to 102 years (Freund & Baltes, 1998). In research with young-old individuals (approximately 70-80 years) and oldest-old individuals (80-90 years), satisfaction with aging was greater when there were sufficient resources, and using an SOC strategy independently increased satisfaction, but only in the young-old group (Jopp & Smith, 2006). For old-old individuals greater resource availability significantly contributed to aging satisfaction; the use of the SOC strategy only affected old-old individuals who reported insufficient availability of resources. Hence, for the oldest people, having sufficient resources is the main contributor to life satisfaction. However, (Jopp & Smith, 2006).

On the other hand, resource deficits (especially in terms of physical and functional health, social networks, and cognitive capacity) are documented in many cultures for young, old, and oldest-old people when contrasted with those experienced when they enter the oldest individuals (Antonucci et al., 2001; Baltes, 1997). It has been suggested that the decline in the use of three of three SOC procedures (selection, optimization, and compensation) between middle and late adulthood is associated with the depletion of resources. However, older adults can compensate for this decline more efficiently using these strategies (Riediger et al., 2005; Nikitin, Freund, 2019).

However, some suggest that using the selection optimization compensation in individuals in different age groups leads to adverse mental health problems (Jopp & Smith, 2006). A study on the influence of SOC competencies on age groups showed that younger and older groups of hospitalized patients manifested no significant difference in their SOC competencies concerning the severity of symptoms of depression on admission to the hospital or during remission (Weiland et al., 2011).SOC is considered a trait variable that may increase or decrease the risk of depression and relapse (Zuroff et al., 1999). Depression will modify SOC competencies (state effect) (Weiland et al., 2011). Interestingly, when the participants were treated with SOC strategies, they were divided into young (21-49), medium (50-66), and old (N67-73) age groups. The medium SOC treatment group exhibited the best results (Weiland et al., 2011). Similarly, the SOC strategies of young and old workers the SOC strategies of young and old workers were compared. A decrease in SOC competencies with age could not be demonstrated (Knecht & Freund, 2017).

### Conceptual diagram

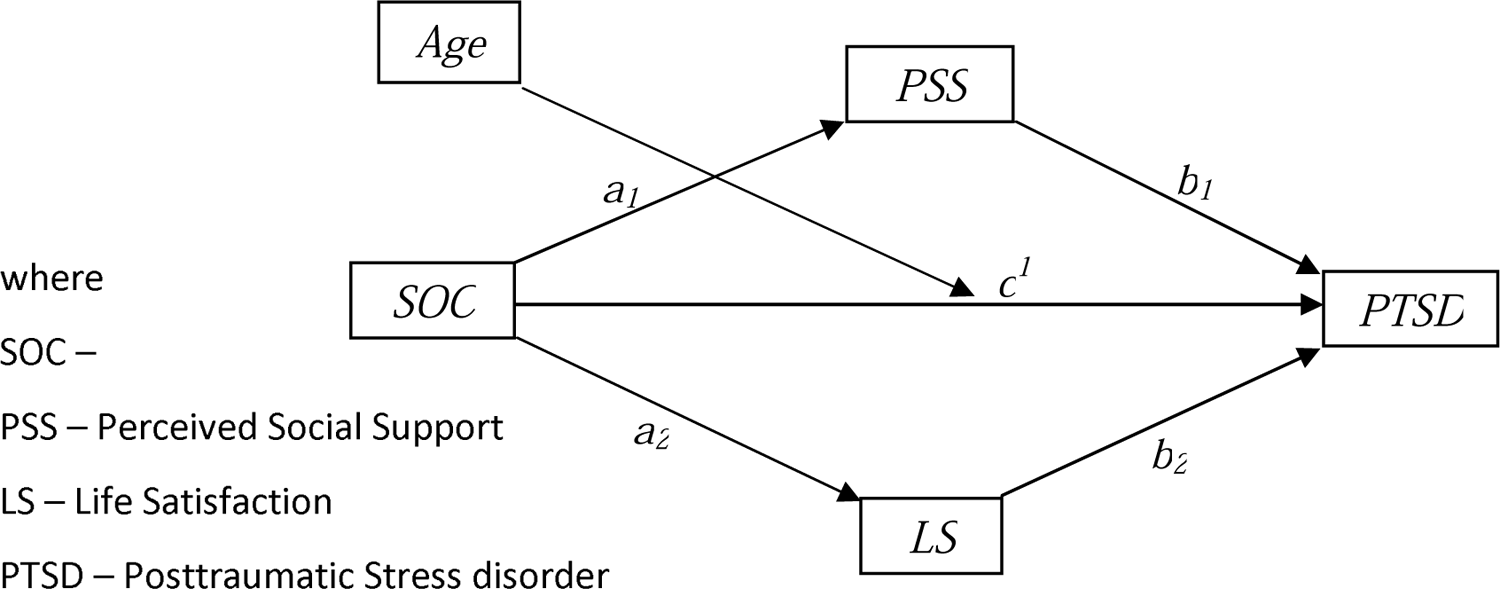

### Sampling procedure

The population of this current study was drawn from Ekiti State, southwestern Nigeria. Ekiti State is the smallest state in southwestern Nigeria and is currently the poorest state, however, the state with the highest life expectancy, highest dependency ratio, highest total fertility rate and highest underemployment rate is the elderly in the six Yoruba States in southwestern Nigeria (NPPSD, 2023; Sasu, 2022; Wan, 2022). The study employed a purposive sampling technique to select four local government areas (LGAs) out of the sixteen LGAs in Ekiti State. This technique was chosen based on the study objectives and the need to meet certain criteria relevant to the study scope.

The rationale for the selection of the five LGAs was driven by several factors, including geographical diversity, population distribution, and the availability of elderly people. One crucial rationale for choosing LGAs with tertiary institutions was the likelihood of these institutions having representatives from various local governments due to the customary quota systems employed in state and federal institutions, which allocate job positions based on demographic criteria.

This system facilitated a balanced representation of employees from different age groups and backgrounds, aligning with the study’s focus on understanding the experiences of older adults within the specified age range (65 to 85+ years). To identify and engage with local farmers and marketers the research team established contact with community leaders, agricultural cooperatives, and local farming and marketer associations within the selected LGAs. These community leaders played a pivotal role in facilitating access to eligible participants. Upon identifying the potential participants, the researcher obtained informed consent from each marketer and farmer before they participated in the study. Clear and culturally sensitive communication was essential during this process to ensure that participants fully understood the research’s objectives and their rights.

### Adaptation of Data Collection

We realized that the collection of data from elderly individuals is often a difficult challenge. Many senior citizens do not want to think of negative experiences, while others have cognitive difficulties, agitated behavior, self-neglect and difficulty with trust. Considering these limitations, it is noteworthy that collecting data on demographic characteristics, particularly when assessing sensitive topics related to mental health and well-being, presented specific challenges. These challenges included reluctance to share information due to privacy concerns and stigma surrounding mental health issues, limited literacy and digital literacy among some participants, and the need to accommodate various cultural and language preferences. To overcome these challenges we introduce the adaptation of data collection methods to ensure that older adults with varying levels of willingness and ability to participate in research could contribute meaningfully to the study. By accommodating these challenges, the study aimed to enhance data quality and participant engagement.

### Questionnaire Administration with Verbal Assistance in Yoruba

To include local farmers within the target age group (65 to 85+ years) in the study, a specific approach was adopted to accommodate potential challenges related to literacy and language proficiency. This approach involved questionnaire administration with verbal assistance in the Yoruba language, the predominant local language in the study area. Questionnaires, designed in English, were distributed to eligible local farmers and marketers who were artisans. Given potential literacy limitations, the questionnaires were carefully structured to include clear and concise language and words that are peculiar in Western countries were modified with words commonly used in Nigeria. All the questionnaires were accompanied by research team members who were fluent in both English and Yoruba. Upon receiving the questionnaire, participants were offered the option of receiving verbal assistance.

When requested, a research team member interpreted and read the questionnaire items aloud to the participants in Yoruba, ensuring that they fully comprehended the questions and response options. The research team was trained to approach this process with cultural sensitivity, respecting the participants’ preferences and ensuring that the verbal assistance did not influence their responses in any way. Participants were assured that their responses would remain confidential and that personal identifiers were not linked to their survey responses, thereby safeguarding their privacy. The final sample of local farmers, artisans and marketers were selected based on their willingness to participate and the completion of the questionnaire either independently or with verbal assistance in Yoruba, as per their choice.

### SOC INSTRUMENT

The short version of the SOC Questionnaire developed and validated by Baltes et al. (1999) was used to assess the behavior of older adults in Ekiti State. The questionnaire was framed with domain-general instructions (we are very interested in knowing how you accomplish things in your life). The measure consists of 12 items that are composed of four components of SOC: elective election selection (ES), loss-based selection (LS), optimization (O), and compensation (C). Each item is presented with a forced choice response scale and consists of two attractive response options, one illustrating the SOC strategy and the other the non-SOC strategy otherwise the response is known as the distracter item.

Participants are requested to choose the response (SOC-related or non-SOC-related distracter) that reflects their behavior on a 4-4-point scale.The scale ranged from “a little to exactly.” For example, one loss-based selection reads, “When things do not go as well as before, I choose one or two important goals” (SOC) and “When things do not go as well as before, I try to keep all my goals” (non-SOC a distracter). A preference for the distracter item is scored as Zero (0), and an aggregate score is calculated by summing all the SOC responses.

The coefficient alphas for the four SOC components were loss-based and elective selection.36, optimization=66, and compensation =35. The items measuring each SOC component were designed to tap into different facets of each component; thus, test-retest reliability is imposed. Previous research has shown satisfactory test–retest stability of(70-.80) for each component (Wiese et al., 2000). Further exploratory and confirmatory factor analyses have also supported the factor structure of the items in the short form of the SOC scale (Freund and Baltes 1998 and 2002). In our study, the reliability statistics revealed a Cronbach’s alpha of 0.648, indicating a moderate level of internal consistency. The moderate internal consistency suggested that there was some reliability in the measurements, but there was room for improvement in the homogeneity of the items within the scale.

### Posttraumatic Stress Disorder Checklist (Civilian Version) (PCL) C

Weathers et al. (1994) developed the PTSD Checklist Civilian Version PCL-C/S, which we used to assess trauma symptoms in the present study. This method aligns with the 4th edition of the Diagnostic and Statistical Manual of Mental Disorders. The scale contains 17 items asking respondents how often they have been bothered by each symptom in the past months on a five-point scale. This instrument is scored by summing all the items on the total severity scale. A total symptom severity score (range ¼ 17–85) can be obtained by summing the scores from each of the 17 items, although cutoff scores of 21, 44, and 35 have been used in previous studies in Nigeria as a screening instrument.

A total score of 44 was considered to indicate PTSD positive. The statement in the PCI-S was made to address three areas of the DSM IV. The IB items are re-experiencing symptoms 1-5; the three items are emotion avoidance symptoms 6-12 and the 2D items are hypervigilance symptoms (13-17).

Researchers have shown the reliability of the PCL-C or PCL-5 to be robust in a sample of motor vehicle accidents and sexual victims, of which 18 were found to have PTSD. Blanchard et al. (1996) reported an alpha of. 94, Ventura et al. (2002) reported a validity of .86 and a test retest score of .80 for individuals in France who had experienced severe traumatic events. In our study, the reliability statistics indicate that the scale has a high level of internal consistency (Cronbach’s alpha= 0.818). This finding implies that the reliability statistics suggest a generally reliable 17-item scale with moderate to high internal consistency.

### Multidimensional Scale of Perceived Social Support (MSPSS)

The PSS-2002 is a 12-item self-report instrument developed by Zimet et al. (1988) that assesses the PSS of the family (11,8,4,3), the PSS of friends (6,7,9,12), and the PSS of significant others (1,2,5,10). The participants responded on a seven-point scale ranging from 1-very strongly disagree to 7-very strongly disagree. A high score means that an individual has a high PSS. The MSPSS has a Cronbach’s alpha of .90. This recent study showed that the MSPSS has a Cronbach’s alpha of 0.92. However, in this recent study, the reliability statistics revaled a Cronbach’s alpha of 0.648, indicating a moderate level of internal consistency. The moderate internal consistency suggested that there was some reliability in the measurements, but there was room for improvement in the homogeneity of the items within the scale.

### Life satisfaction Index-z (LSIZ)

This is a survey of 18 items developed by Neugarten, Havighust, and Tobin (1961), which are statements about life in general that people feel differently about. A respondent is expected to rate each item 1, 2, or 3, depending on how the item describes how he/she feels. The number stands for 1 = Disagree, 2 =Unsure, 3 = Agree. All 18 items are scored directly by adding all the scores together, with the lowest score being 18 and the highest score being 54. The lower the score is, the lower the satisfaction with life; the higher the score is, the greater the satisfaction with life. The LSIZ is one of the most widely used measures of well-being in later life. As such, considerable attention has been given to its factor structure (Helmes et al., 1998; Hoyt & Creech, 1983; Liang,1984), including investigations of racial and ethnic differences (Liang et al., 1987; Liang et al., 1988; Shmotkin, 1991; Tran, 1992). This reliability generalization involved a search of 157 journal articles, which resulted in the inclusion of a total of 34 samples. The results revealed an average reliability of .79 (SD =.10, median =.79).

Bivariate correlational analyses revealed no relationships between score reliability and various sample characteristics, including sample size, number of items, mean age, standard deviation of age, proportion of females, mean LSI score, or standard deviation of LSI scores. However, in this present study, the reliability statistics indicate that the scale has a moderate to high level of internal consistency (Cronbach’s alpha = 0.770). This finding implies that the reliability statistics suggest a generally reliable 18-item scale with moderate to high internal consistency.

## RESULTS

We administered 376 questionnaires to the target population, and after scanning, we observed that some respondents were younger than the age range therefore, 321 were selected. We divided the participants into three age groups: young (65-74) 59.2% old (75-84) 23.4% and oldest (85+) 17.4%.

Overall, 81% (260) were male, female19% (61) were female, while 65.1% (209) werenChristian, 24.3% (78) were Muslim and 9% (29) were from the African traditional region. Additionally, 72.3% (232) were married, 8.1% (26) were separated, 3.1% (10) were divorced, and 26.5% (53) were widows. Again, 60.7 (195) of the respondents had one wife, 17.8% (57) had two wives, 12.1% (39) had three wives, and 9.4% (30) had four wives or more. Similarly, 3.1% (10) had no child, 4.7% (15) had one child, 4.7% (15) had two children, and 60% (199) had four wives or more. Indeed, 1.6% (5) could not read or write, 14.9% (48) had a primary education, 9.7% (31) had a secondary education, 22.7% (73) were above secondary school, 28% (90) had a first-degree education (90) and 23% (74) had a master’s degree or above. Furthermore, 31.2% (100) were schoolteachers, 10% (32) were traders, 24.9% (80) were farmers, 15.9% (51) were government workers, 10.3% (31) were professionals, and 7.8% (25) had nothing to do. However, 30.8% (99) had good health, 46.7% (150) had average health, 17.8% (57) could say their health status, and 4.7% (15) had poor health conditions. Approximately 95% of the 305 lived in their own house, and 5% (16) lived in rented apartments.

**Table 1:**
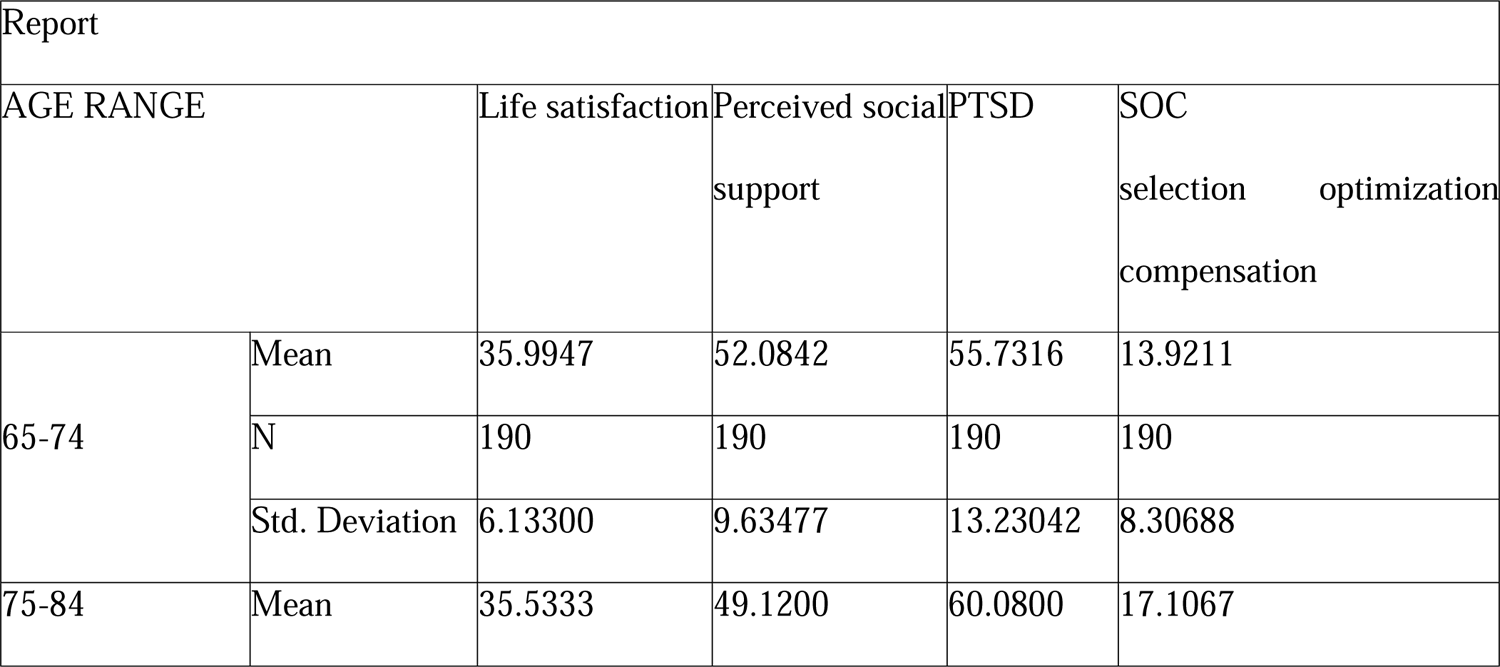
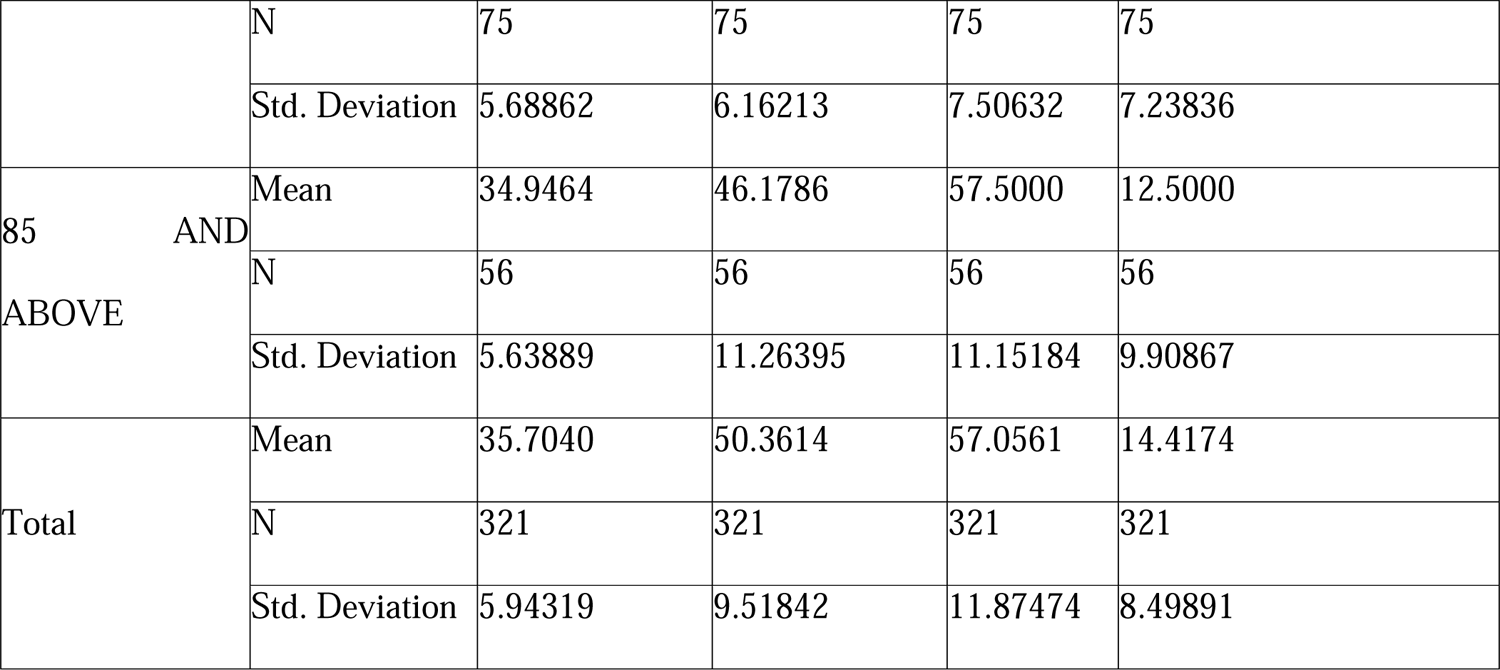
Means: Descriptive Statistics of the Variables across Age Groups Life Satisfaction: The mean life satisfaction scores seem to decrease slightly with increasing age. The pattern suggests that, on average, individuals in the 65-74 age range report slightly greater life satisfaction than do those in the 75-84 age range, and the life satisfaction further decreases for those aged 85 and above.

Perceived social support: Similar to life satisfaction, perceived social support also shows a decreasing trend with age. The mean scores decreased from the 65-74 years to the 85 years and older. Posttraumatic stress disorder: The mean PTSD scores increased with age. Individuals in the 75-84 age range had the highest mean PTSD score, followed by those in the 85 and above age range. This suggests that, on average, individuals in these age groups may experience higher levels of PTSD. Selection optimization compensation: The mean SOC scores show variability across age ranges. The 75-84 age range had the highest mean SOC score, suggesting a potential peak in the use of selection, optimization, and compensation strategies in this age group.

Therefore based on these findings, there is a trend toward decreasing life satisfaction and perceived social support with age. This finding implies that perceived social support is pivotal in enhancing the life satisfaction of the older people in the Ekiti State. To increase their level of life satisfaction, they need to increase their level of perceived social support. While PTSD scores show an increase with age, it could be said that the increase in PTSD scores with age, particularly in the 75-84 age range, raises questions about the relationship between age and the prevalence or severity of PTSD. It could be that individuals in this age range have experienced traumatic events due to the resource depletion of Ekiti State. The SOC scores varied across age ranges, with a peak in the 75-84 age range which could suggest that the use of selection, optimization, and compensation strategies may vary throughout the aging process. On the basis of these findings, we tested an ANOVA to find individual differences in these factors to obtain a comprehensive understanding of the findings.

**Table 2.**
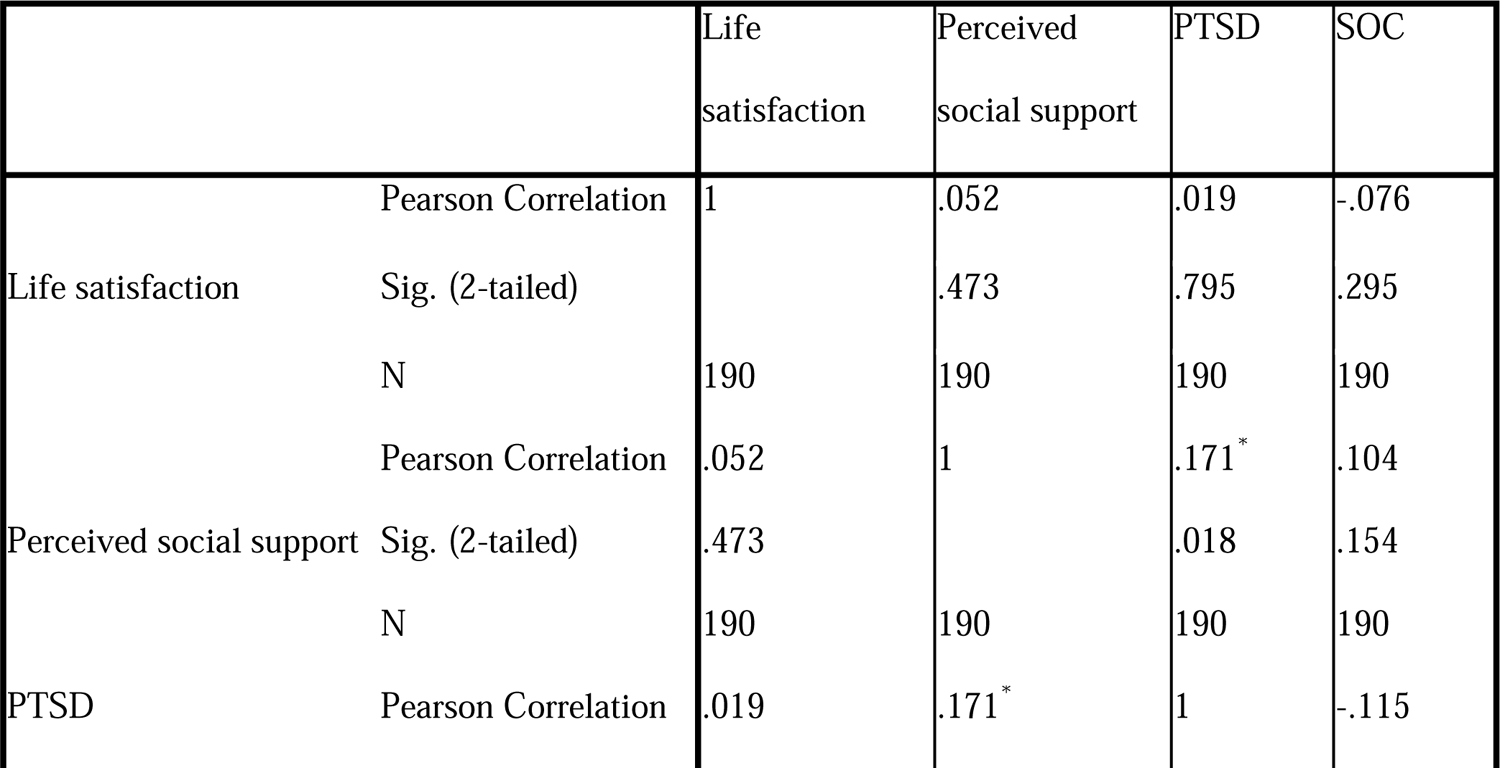
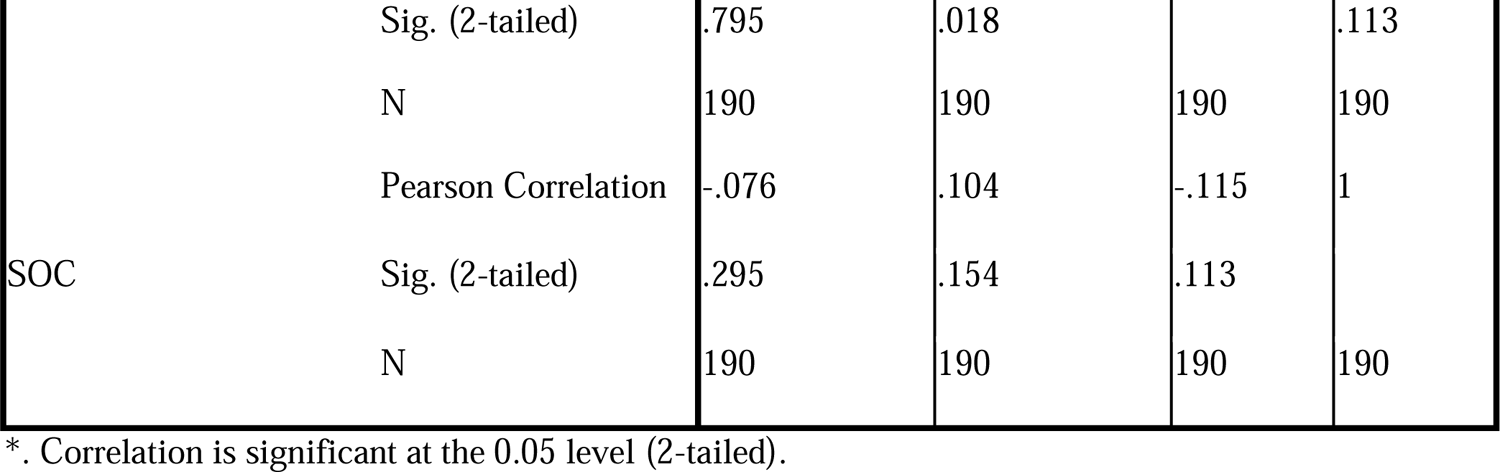
Correlations in the variables of Age Groups 65-74.

In the 65-74 age group, the correlations among life satisfaction, perceived social support, posttraumatic stress disorder (PTSD), and selection optimization compensation (SOC) revealed subtle relationships. Life satisfaction weakly positively correlated with perceived social support, suggesting that there are tendency for those individuals with higher social support tend to report greater life satisfaction, although this association is not statistically significant. Conversely, the correlation between life satisfaction and both PTSD and SOC was extremely weak and lacked statistical significance, indicating that there was no meaningful relationship between these variables. In terms of perceived social support, a weak positive correlation with PTSD was observed, implying that individuals with higher perceived social support may also experience slightly elevated PTSD symptoms. The correlations between life satisfaction and the SOC, while positive, are very weak and statistically insignificant. For PTSD, the correlation with perceived social support was weakly positive, indicating that those individuals with higher PTSD symptoms may report slightly greater perceived social support. The correlations between life satisfaction and the SOC, however, are extremely weak and not statistically significant. Concerning SOC, the correlation with perceived social support was weakly positive, suggesting a minor tendency for individuals with a stronger SOC to report greater perceived social support. The correlations between life satisfaction and PTSD severity were both weak and lacked statistical significance, indicating limited meaningful associations. These findings underscore the intricate nature of mental health dynamics in the senior citizens population, highlighting the need for nuanced interventions that consider the multifaceted relationships between life satisfaction, perceived social support, PTSD, and SOC.

**Table 3:**
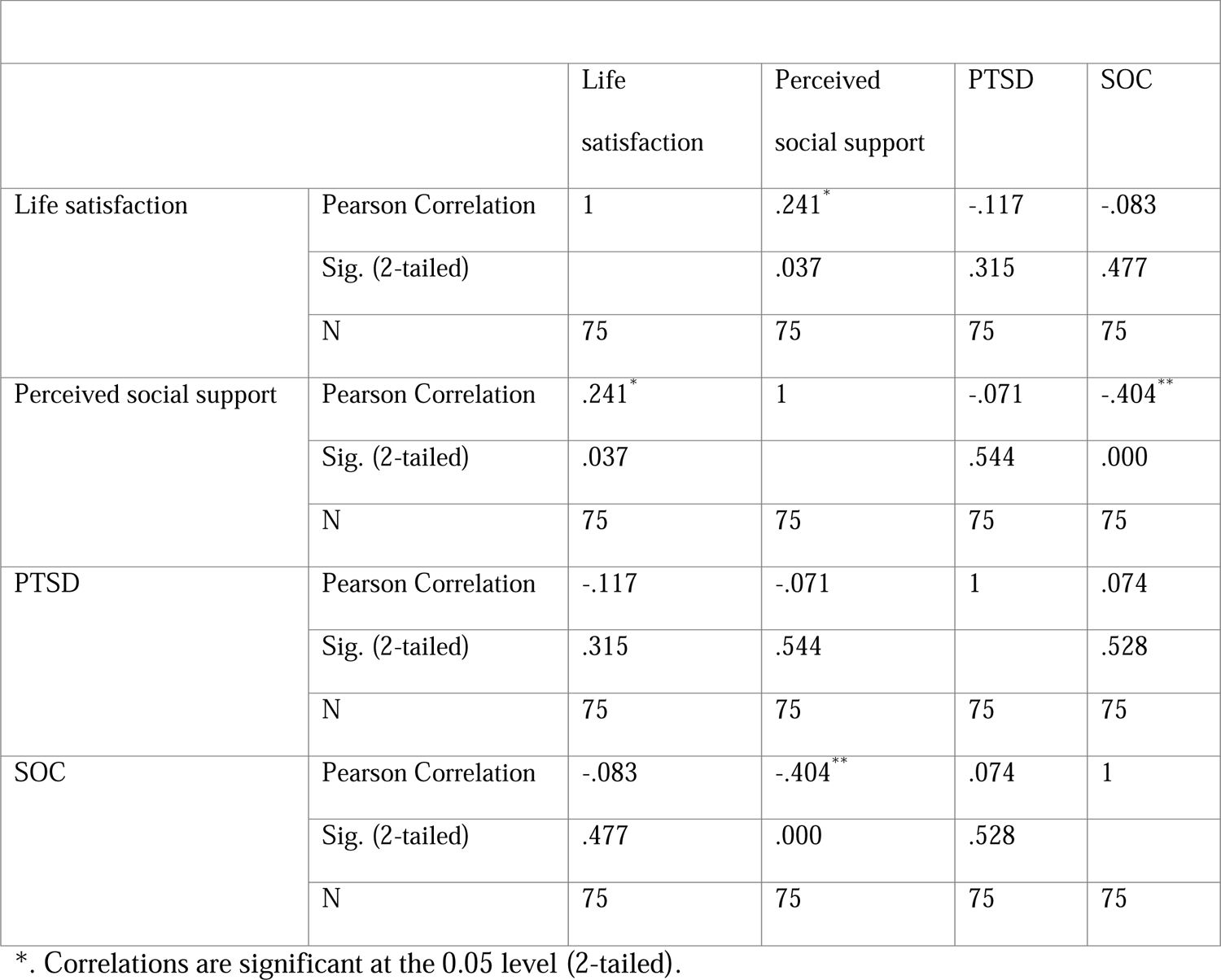
Correlations for the Age Groups 75-84 Variables.

In the 75-84 age group, the correlations among life satisfaction, perceived social support, posttraumatic stress disorder (PTSD), and selection optimization compensation (SOC) revealed interesting patterns. There was a weak positive correlation (0.241) between life satisfaction and perceived social support, suggesting that individuals with higher perceived social support are more likely to report greater life satisfaction (p = 0.037). The correlation with PTSD was weakly negative (−0.117), hinting at a minor tendency for individuals with higher PTSD symptoms to report lower life satisfaction, although this association was not statistically significant (p = 0.315). There was no statistically significant relationship between life satisfaction and the SOC, as indicated by the very weak negative correlation of −0.083 (p = 0.477). A very weak negative correlation (−0.071) with PTSD suggested that there was no meaningful relationship between perceived social support and PTSD symptoms in this age group (p = 0.544). A notable negative correlation of −0.404 with SOC indicates that individuals with higher perceived social support are more likely to have a stronger SOC (p = 0.000). The correlation with life satisfaction was weakly negative (−0.117), suggesting a minor tendency for individuals with higher PTSD symptoms to report lower life satisfaction, although this association was not statistically significant (p = 0.315).

The correlation with perceived social support was very weakly negative (−0.071), indicating that there was no meaningful relationship between perceived social support and PTSD symptoms (p = 0.544). There was no statistically significant relationship between PTSD and SOC, as indicated by the very weak positive correlation of 0.074 (p = 0.528). Additionally, the correlation with life satisfaction is very weakly negative (−0.083), suggesting that there is no meaningful relationship between SOC and life satisfaction in this age group (p = 0.477). A strong negative correlation of −0.404 with perceived social support indicates that individuals who use SOC strategies are more likely to report greater perceived social support (p = 0.000). There was no statistically significant relationship between SOC content and PTSD severity, as indicated by the very weak positive correlation of 0.074 (p = 0.528). The findings imply that social support plays a crucial role in influencing life satisfaction in individuals aged 75-84 years, highlighting the importance of fostering supportive networks for their well-being. The negative correlation between perceived social support and SOC suggested a complex interplay between social and psychological factors in this age group. While some correlations are statistically insignificant, the trends hint at nuanced relationships that warrant further exploration. These findings underscore the need for tailored interventions that consider both social and psychological aspects to enhance the overall well-being of older adults in this age group.

**Table 4:**
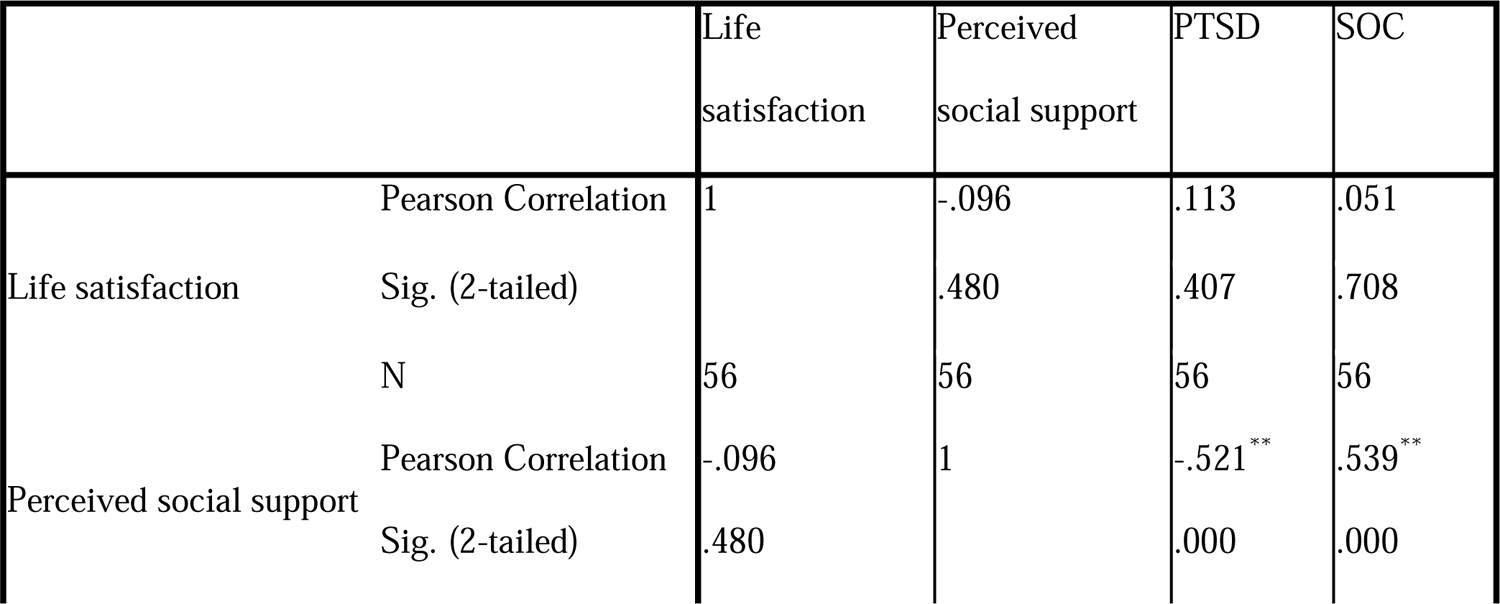
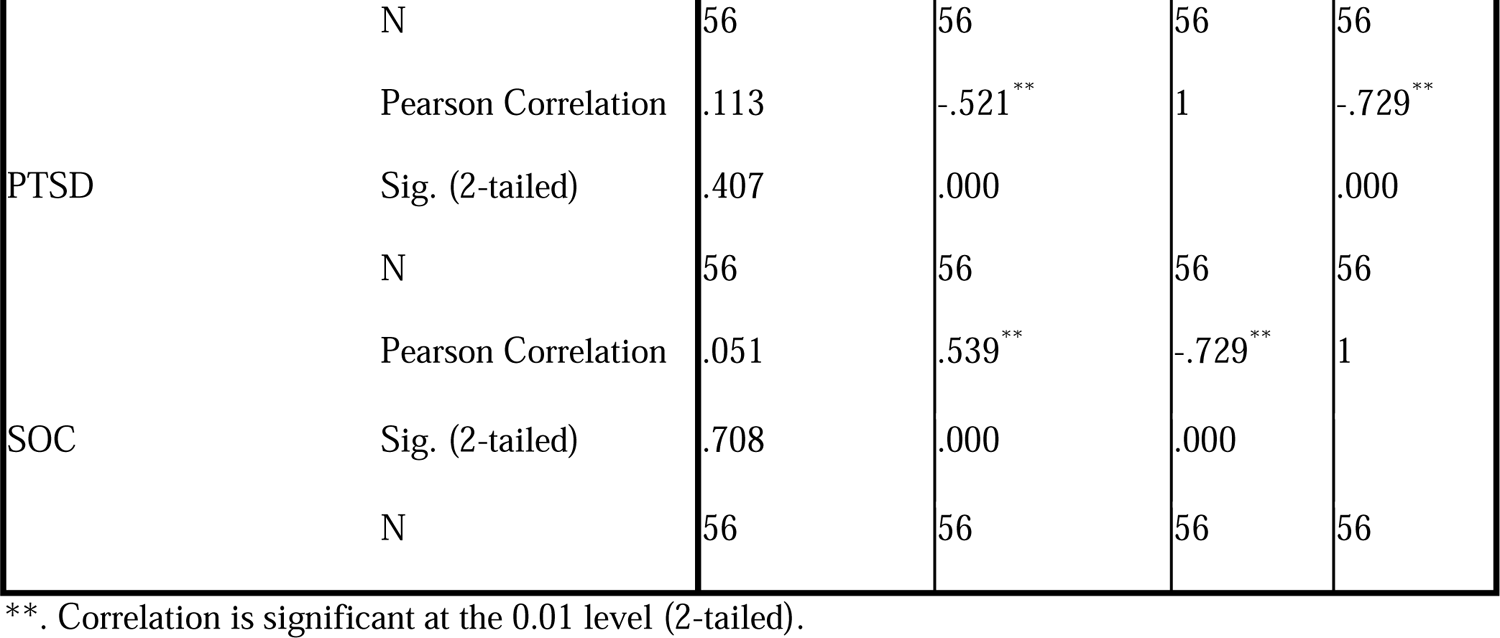
Correlations in the variables of Age Groups 85 and above.

The Pearson correlation coefficient was −0.096 (p = 0.480), indicating a very weak negative correlation between life satisfaction and perceived social support. This finding suggested that there was no statistically significant relationship between these variables in the 85 years or older group. The correlation with PTSD was 0.113 (p = 0.407), indicating a very weak positive correlation. This finding suggested that there was no statistically significant relationship between life satisfaction and PTSD in this age group. The correlation coefficient with SOC was 0.051 (p = 0.708), indicating a very weak positive correlation. This finding suggested that there was no statistically significant relationship between life satisfaction and SOC in individuals aged 85 years or older. The correlation with life satisfaction is −0.096 (p = 0.480), indicating a very weak negative correlation. This finding suggested that there was no statistically significant relationship between perceived social support and life satisfaction in this age group. The correlation with PTSD was −0.521 (p = 0.000), indicating a strong negative correlation. This finding suggested that individuals with greater perceived social support are more likely to have lower PTSD symptoms. The correlation coefficient with SOC was 0.539 (p = 0.000), indicating a strong positive correlation. This finding suggested that individuals with higher perceived social support are more likely to have a stronger SOC. The correlation with life satisfaction was 0.113 (p = 0.407), indicating a very weak positive correlation. This finding suggested that there was no statistically significant relationship between PTSD and life satisfaction in individuals aged 85 years or older.

Furthermore, the correlation with perceived social support is −0.521 (p = 0.000), indicating a strong negative correlation. This finding suggested that individuals with higher PTSD symptoms are less likely to report greater perceived social support. The correlation with SOC is −0.729 (p = 0.000), indicating a very strong negative correlation. This finding suggested that individuals with more PTSD symptoms are less likely to SOC strategies. The correlation with life satisfaction is 0.051 (p = 0.708), indicating a very weak positive correlation. This finding suggested that there was no statistically significant relationship between SOC and life satisfaction in individuals aged 85 year or older. The correlation with perceived social support is 0.539 (p = 0.000), indicating a strong positive correlation. This finding suggested that individuals who use SOC methods are more likely to report greater perceived social support. The correlation with PTSD was −0.729 (p = 0.000), indicating a very strong negative correlation. This finding suggested that individuals who strongly use SOC strategies are less likely to have higher PTSD symptoms suggesting the potential importance of adaptive coping mechanisms in reducing PTSD symptoms.

Indeed, in individuals aged 85 and above, there was no statistically significant relationship between life satisfaction and perceived social support or SOC. This finding suggested that factors influencing life satisfaction may be more complex in this age group. The strong negative correlation between perceived social support and PTSD symptoms implies that higher levels of perceived social support are associated with lower PTSD symptoms in individuals aged 85 years and older. This finding underscores the potential protective role of social support against PTSD in this population. The strong positive correlation between perceived social support and SOC suggested that individuals with greater perceived social support are more likely to make use of SOC strategies. This highlights the interconnectedness of social support and psychological resilience in older adults. The strong negative correlation between SOC and PTSD aligns with the concept of resilience, where individuals who effectively utilize adaptive coping mechanisms are better equipped to manage and overcome the impact of stressors, potentially reducing the risk of developing PTSD.

**Table 5:**
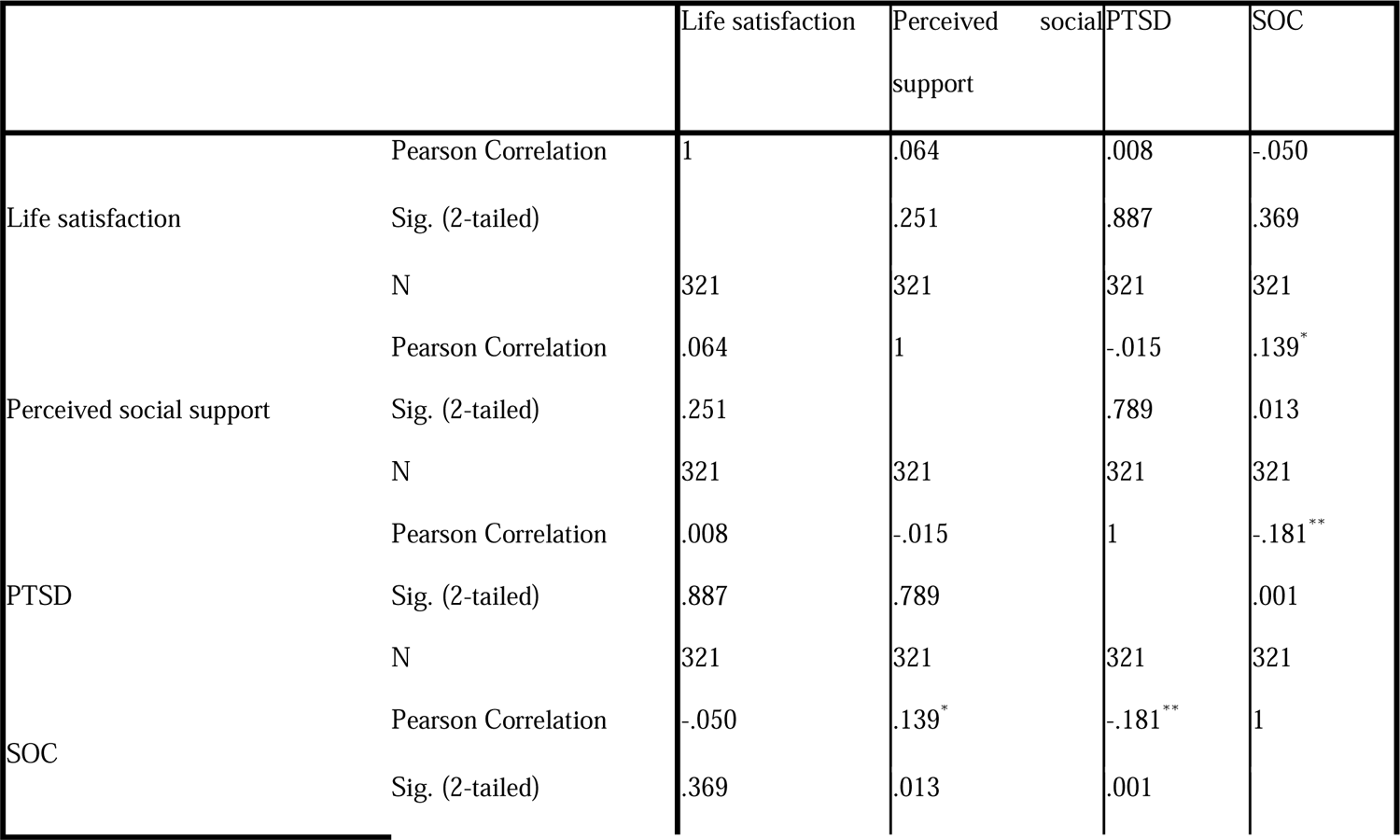
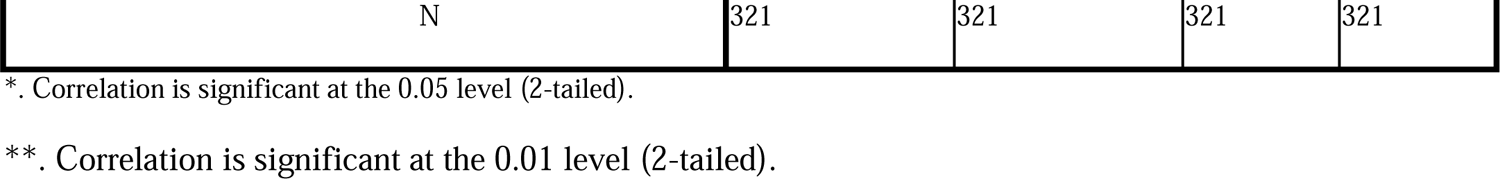
Correlations of the Age Groups of the entire sample.

The Pearson correlation coefficient was 0.064 (p = 0.251), indicating a very weak positive correlation between life satisfaction and perceived social support. However, this correlation was not statistically significant, suggesting that there was no clear relationship between these variables. The correlation with PTSD was 0.008 (p = 0.887), indicating an extremely weak positive correlation. The lack of statistical significance suggests that there was no meaningful relationship between life satisfaction and PTSD symptoms. The correlation with SOC is −0.050 (p = 0.369), indicating a very weak negative correlation. However, this correlation was not statistically significant, suggesting that there was no clear relationship between life satisfaction and SOC. The correlation with life satisfaction was 0.064 (p = 0.251), indicating a very weak positive correlation. This correlation was not statistically significant, suggesting that there was no clear relationship between perceived social support and life satisfaction. The correlation with PTSD was −0.015 (p = 0.789), indicating an extremely weak negative correlation. The lack of statistical significance suggested that there was no meaningful relationship between perceived social support and PTSD symptoms. The correlation coefficient with SOC was 0.139 (p = 0.013), indicating a weak positive correlation. This correlation is statistically significant, suggesting that individuals with higher perceived social support are more likely to have a stronger SOC. The correlation with life satisfaction is 0.008 (p = 0.887), indicating an extremely weak positive correlation. The lack of statistical significance suggests that there is no meaningful relationship between PTSD and life satisfaction. The correlation with perceived social support is −0.015 (p = 0.789), indicating an extremely weak negative correlation. The lack of statistical significance suggests that there is no clear relationship between PTSD symptoms and perceived social support. The correlation coefficient with SOC was −0.181 (p = 0.001), indicating a moderate negative correlation. This correlation was statistically significant, suggesting that individuals with higher PTSD symptoms are less likely to employ SOC strategies.

The correlation with life satisfaction is −0.050 (p = 0.369), indicating a very weak negative correlation. However, this correlation was not statistically significant, suggesting that there was no clear relationship between SOC and life satisfaction. The correlation coefficient with perceived social support is 0.139 (p = 0.013), indicating a weak positive correlation. This correlation is statistically significant, suggesting that individuals who embark on the use of stronger SOC strategies are more likely to report greater perceived social support. The correlation with PTSD was −0.181 (p = 0.001), indicating a moderate negative correlation. This correlation was statistically significant, suggesting that individuals who utilized SOC strategies were less likely to have greater PTSD symptoms.

Overall, the correlations between life satisfaction and perceived social support and between TSD and SOC are extremely weak or not statistically significant. The significant correlation between perceived social support and SOC implies that individuals with greater perceived social support are more likely to apply SOC strategies. This highlights the potential interconnectedness of social support and psychological resilience. The lack of significant correlations between life satisfaction and other variables, such as PTSD and the SOC, underscores the complexity of factors influencing life satisfaction.

### One-way ANOVA

The descriptive statistics provide insights into the key variables of life satisfaction, perceived social support, posttraumatic stress disorder (PTSD), and selection optimization compensation (SOC) across different age groups. For life satisfaction, the mean scores indicate a subtle decline with increasing age, ranging from 35.99 in the 65-74 years age group to 34.95 in the 85 years and older age group. However, the differences are relatively small, and the standard deviations suggest variability within each age group, with an overall moderate level of life satisfaction across all ages (mean = 35.70). Similarly, perceived social support gradually decreased from 52.08 in the 65-74 age group to 46.18 in the 85 and above group. The standard deviations indicate variability within age groups, with a moderate overall level of perceived social support (mean = 50.36). For PTSD, the mean scores varied from 55.73 in the 65-74 years age group to 57.50 in the 85 years and older age group. Standard deviations highlighted variability within age groups, reflecting a moderate overall level of PTSD symptoms (mean = 57.06). In terms of SOC, the mean scores ranged from 13.92 in the 65-74 years age group to 12.50 in the 85 years and older age group, with SDs indicating variability within the age groups. The overall mean suggested a moderate level of SOC strategies (mean = 14.42). While there are subtle variations in mean scores across age groups, the standard deviations emphasize the diversity of experiences within each category.

**Table 6:**
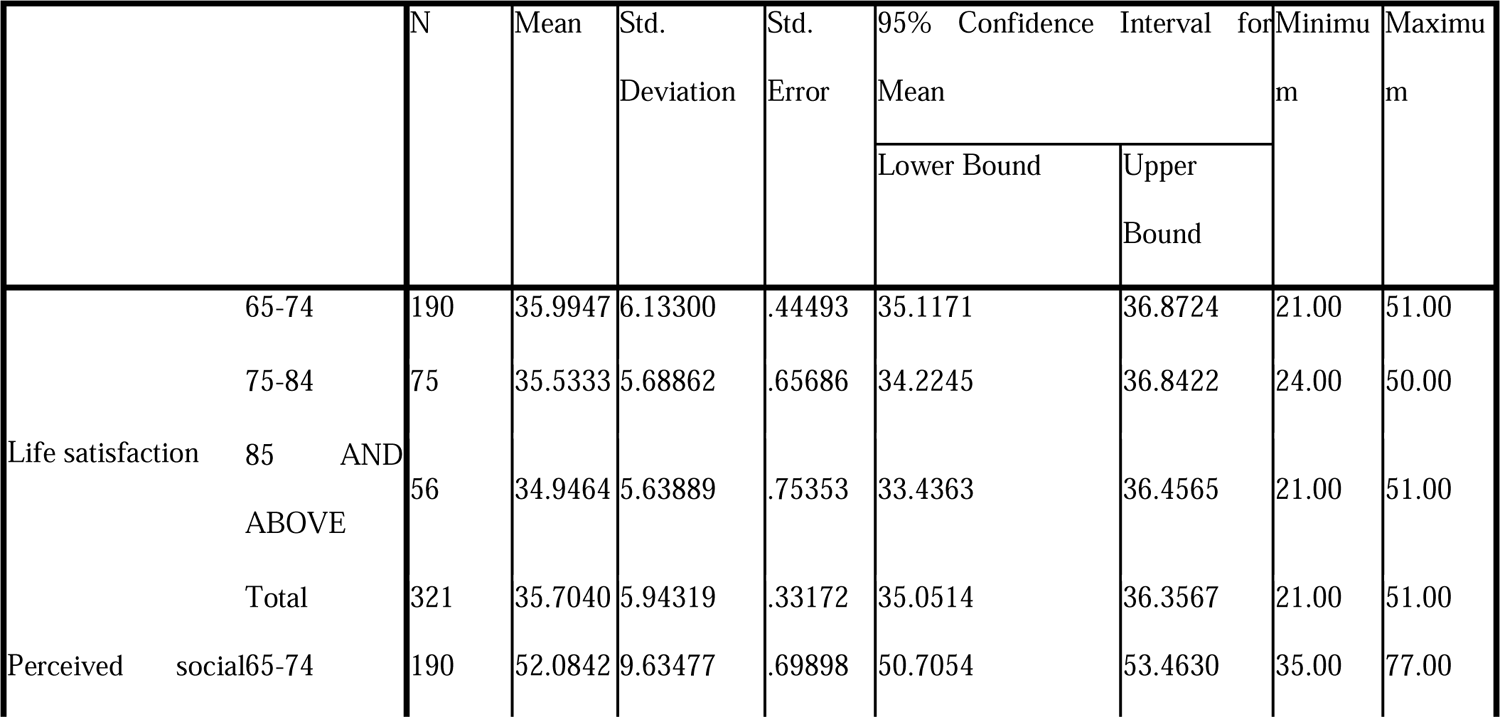
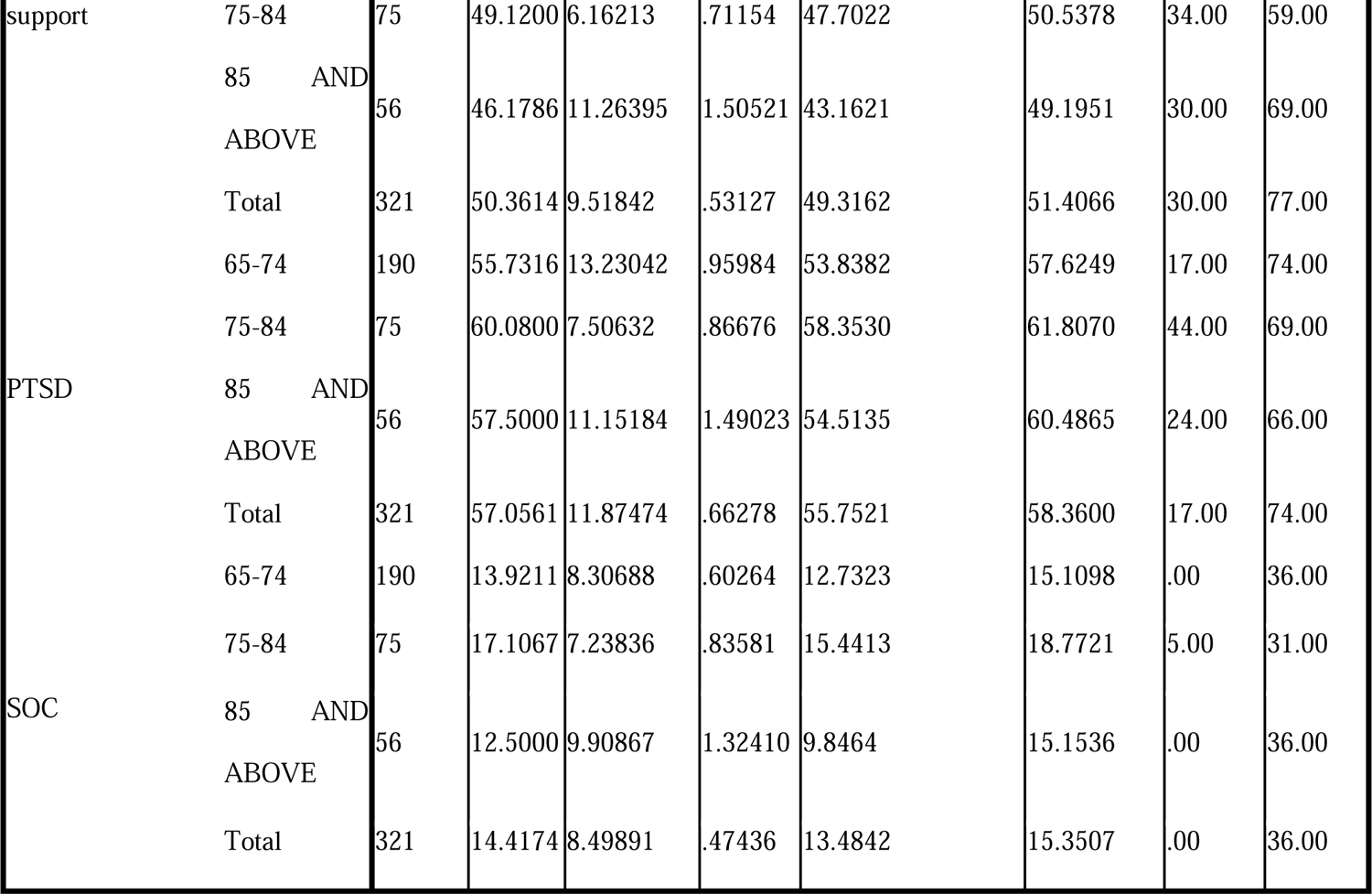
ANOVA Results of the Variables Across Age Groups.

**Table 7:**
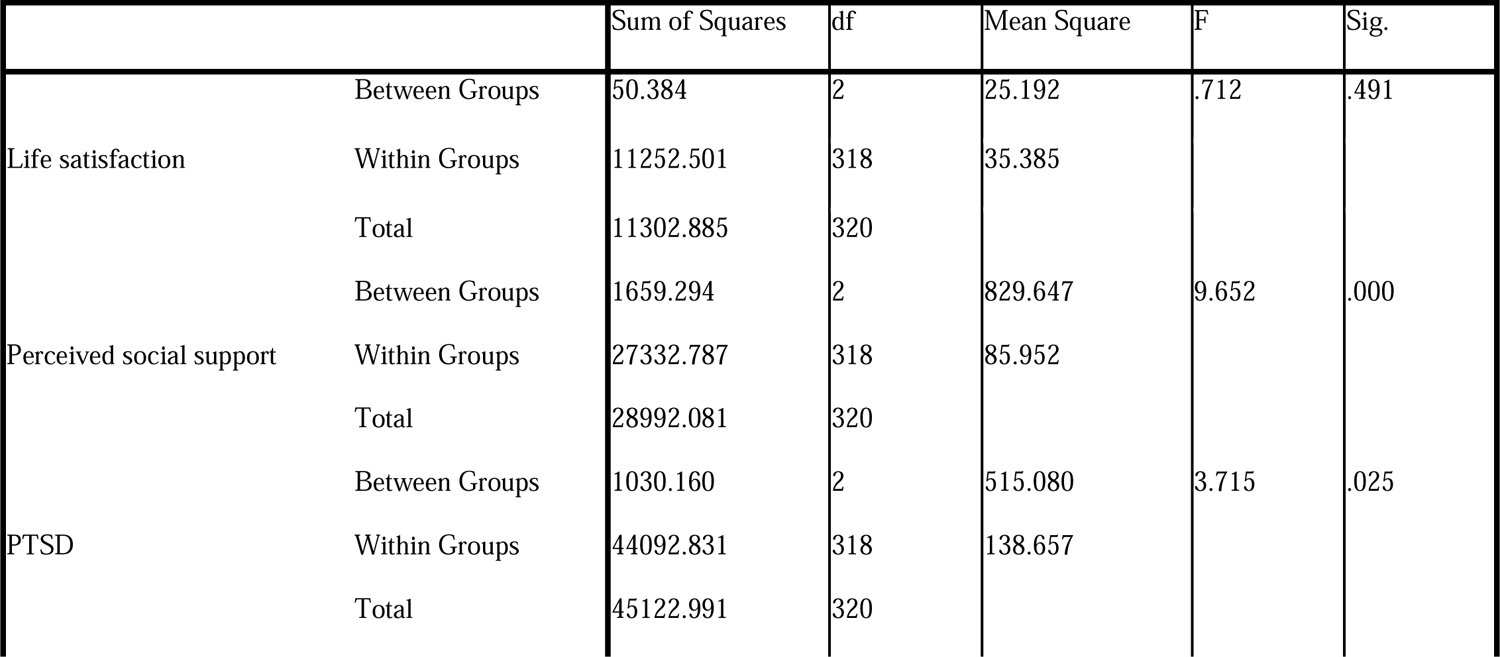
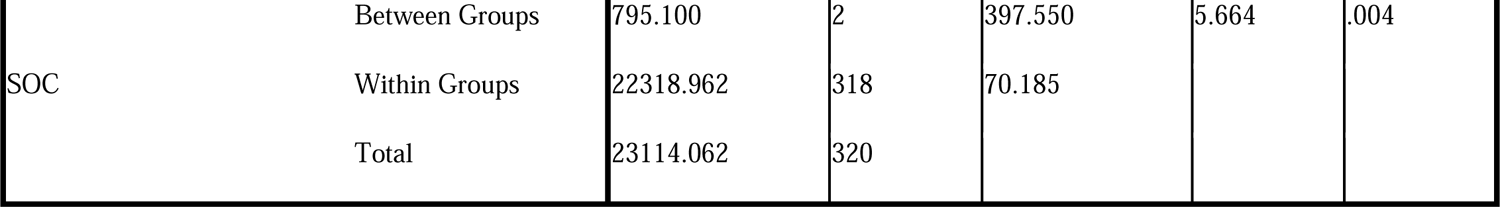
ANOVA Assessing Statistical Differences Between Age Groups for Variables.

The results of the one-way Analysis of Variance (ANOVA) shed light on the variations in the four variables across different age groups, providing insights into life satisfaction, perceived social support, posttraumatic stress disorder (PTSD), and selection optimization compensation (SOC). Regarding life satisfaction, the ANOVA did not reveal a statistically significant difference in mean scores among age groups, implying that the observed variation could be due to chance and that age may not play a substantial role in influencing overall life satisfaction levels. In contrast, the ANOVA results for perceived social support revealed a significant difference in mean scores across age groups. This finding suggested that individuals in distinct age categories experience varying levels of perceived social support, emphasizing the need for targeted interventions tailored to the specific social support needs of different age groups. Similarly, the ANOVA results for PTSD demonstrated a statistically significant difference in mean scores among age groups, indicating that individuals in different age categories exhibit distinct levels of PTSD symptoms. This underscores the importance of considering age-related factors when addressing and managing posttraumatic stress. Additionally, the results for SOC yielded a significant result, emphasizing that individuals in various age groups possess differing levels of SOC. Understanding these variations is essential for tailoring interventions that enhance individuals ability to comprehend and manage challenges in their lives. These findings underscore the nuanced relationship between age and mental health variables, highlighting the importance of age-specific approaches in designing effective interventions and support systems. Further exploration through posthoc analyses provided deeper insights into the specific age groups driving these observed differences.

### Post Hoc Test

#### Multiple Comparisons

Post hoc analyses, employing the Scheffe multiple comparisons method, were performed to delve deeper into the differences in mean scores for life satisfaction, perceived social support, posttraumatic stress disorder (PTSD), and selection optimization compensation (SOC) across distinct age groups. No statistically significant differences were observed in life satisfaction between the 65-74 age group and either the 75-84 or the 85 and above age groups. This finding suggested that, within this sample, age does not play a substantial role in influencing overall life satisfaction. Moreover, significant differences emerged in perceived social support between the 65-74 and 85 years and older age groups, with the latter reporting significantly greater levels of social support. No significant differences were found between the 65-74 and 75-84 age groups or between the 75-84 and 85 and above age groups. Statistically significant disparities in PTSD scores were noted between the 65-74 and 75-84 age groups, with the latter reporting higher PTSD scores.

**Table 8:**
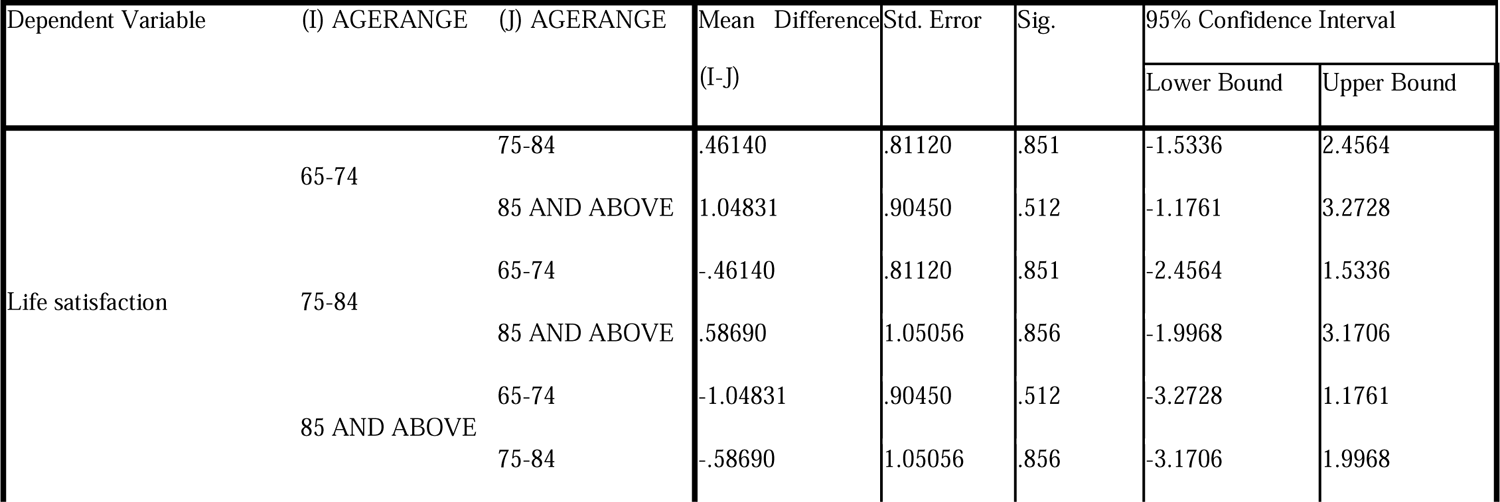
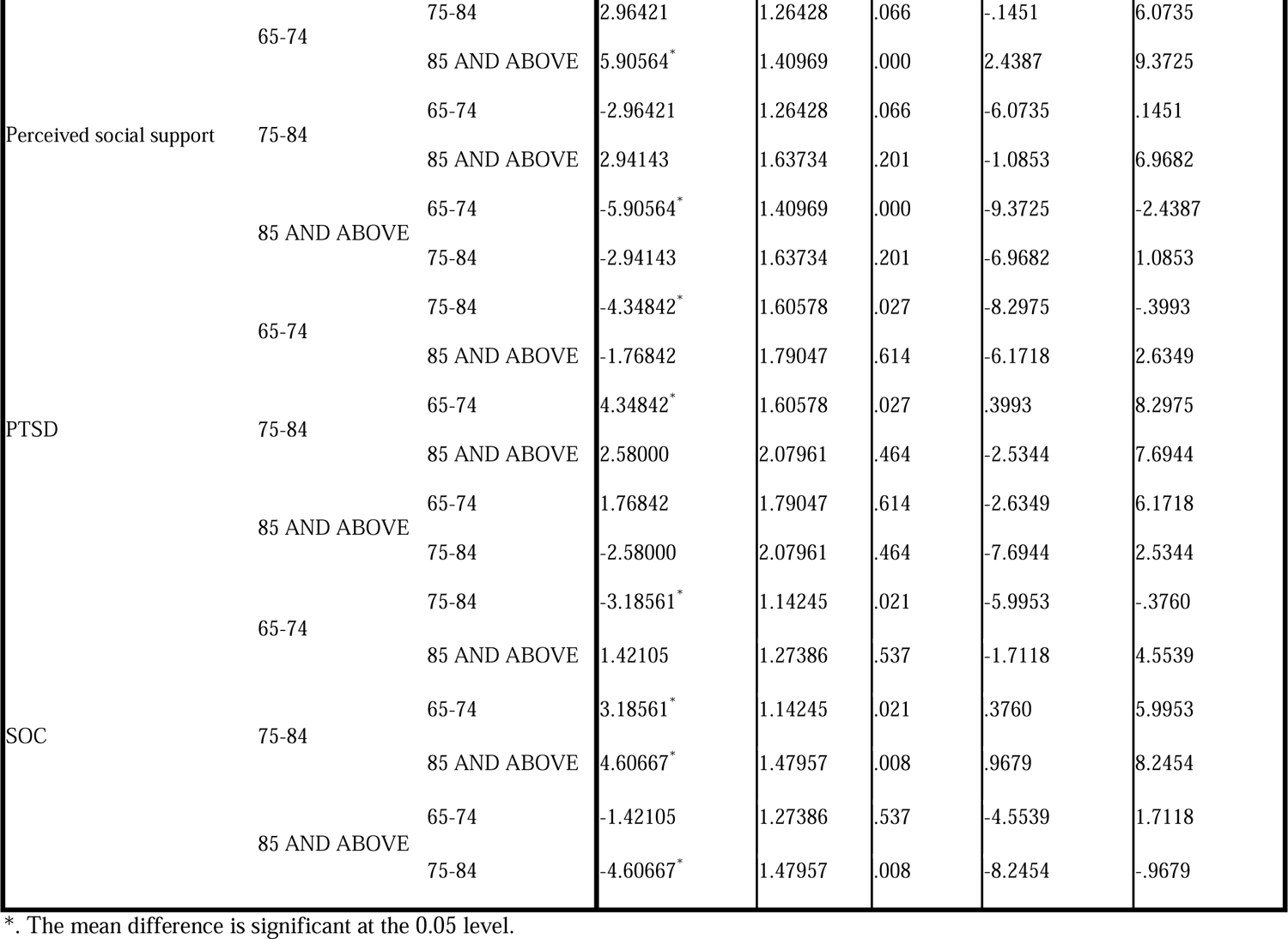
Post Hoc Test (Scheffe) of the mean difference between Age Groups of the variables.

In addition, no significant differences were observed in PTSD scores between the 65-74 and 85 years or between the 75-84 and 85 years and older age groups. The results revealed significant differences in SOC scores between the 65-74 and 75-84 age groups, with the 75-84 age group reporting higher SOC scores. Similarly, a significant difference was observed between the 65-74 and 85 years and older age groups, with the latter reporting higher SOC scores. There was no significant differences in SOC scores between the 75-84 years and 85 years or older age groups. These findings imply that life satisfaction is not significantly influenced by age group within this study, emphasizing the need to explore other contributing factors. The greater perceived social support in the 85 years and older age group suggested potential areas for targeted support programs, recognizing the unique social needs of this demographic. Elevated PTSD scores in the 75-84 age group highlight a specific area for intervention and support tailored to the mental health challenges of this age cohort. Moreover, differences in SOC scores emphasize the importance of considering age-related factors in strategies aimed at enhancing individuals’ selection optimization compensation, particularly within the 75-84 and 85 years and older age groups.

### RESULTS OF INFLUENTIAL STATISTICS AND INTERPRETATION

In this study, a moderated mediation analysis was implemented using theHayes model 5 macro process based on the data collected from the participants. According to the hypotheses raised for the study, the results are presented in the following order.

H_1_: Will the SOC influence perceived social support among the senior citizens in Ekiti State?

**Table 9:**
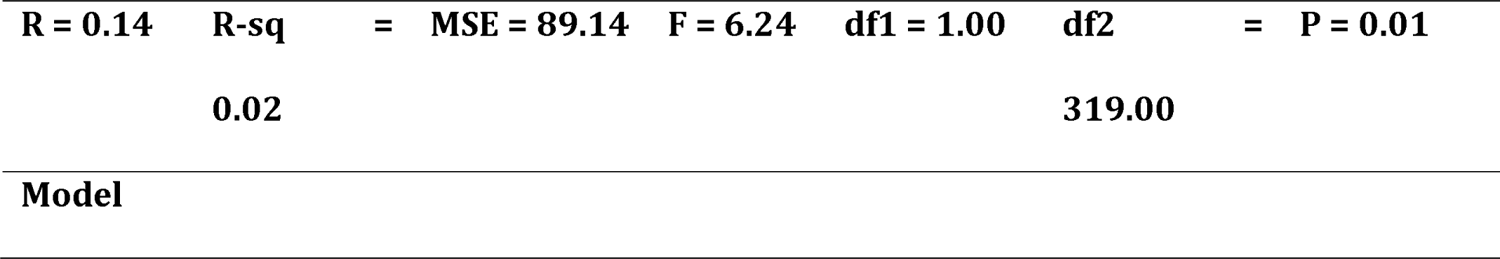
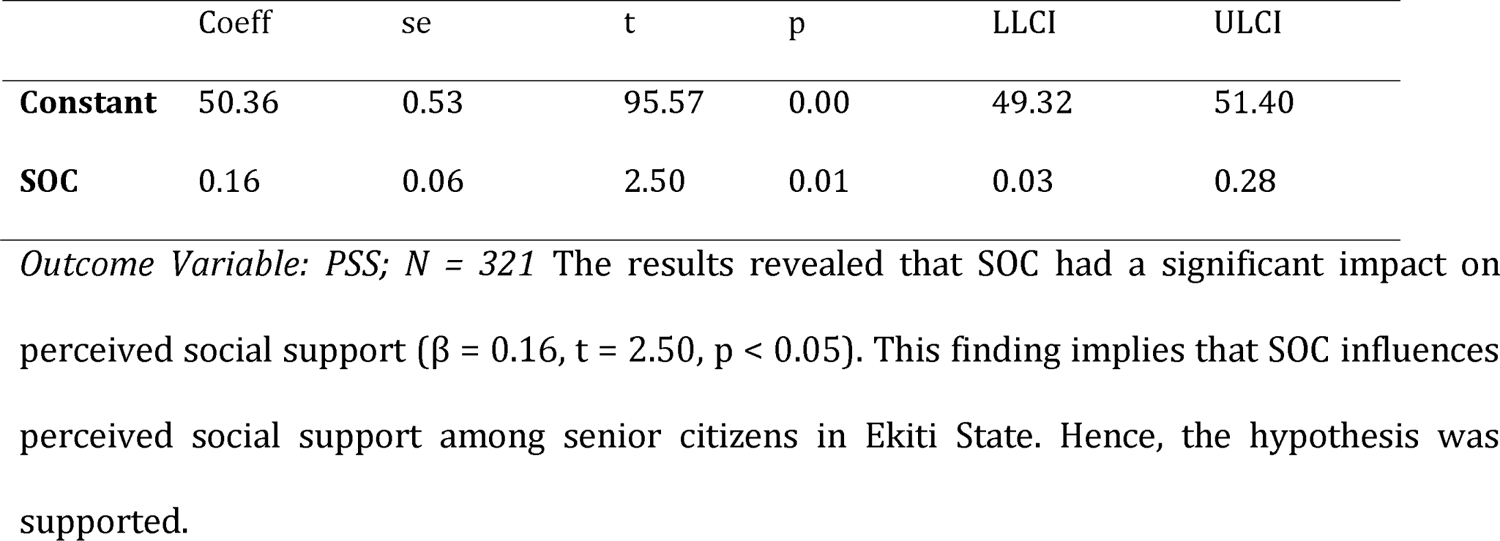
Impact of the SOC on the PSS.

H_2_: Will the SOC influence life satisfaction among senior citizens in Ekiti State?

**Table 10.**
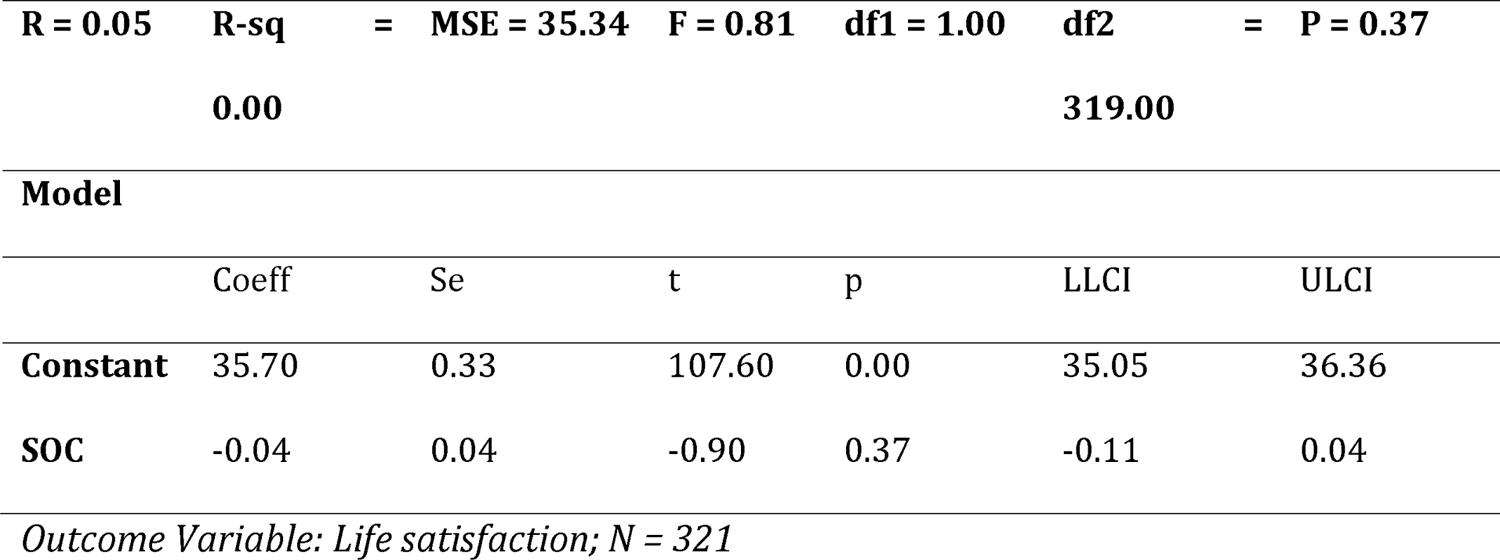
Impact of the SOC on life satisfaction.

| **R = 0.05** | **R-sq** | **=** | **MSE = 35.34** | **F = 0.81** | **df1 = 1.00** | **df2** | **=** | **P = 0.37** |
| --- | --- | --- | --- | --- | --- | --- | --- | --- |
|  | **0.00** |  |  |  |  | **319.00** |  |  |
| **Model** |  |  |  |  |  |  |  |  |
|  | Coeff | Se | t | p | LLCI | ULCI |  |  |
| **Constant** | 35.70 | 0.33 | 107.60 | 0.00 | 35.05 | 36.36 |  |  |
| **SOC** | -0.04 | 0.04 | -0.90 | 0.37 | -0.11 | 0.04 |  |  |

*Outcome Variable: Life satisfaction; N = 321*

The results revealed that the SOC had an insignificant impact on life satisfaction (β = −0.04, t = −0.90, p_(0.37)_ > 0.05). This finding implies that the SOC does not influence life satisfaction among the senior citizens in Ekiti State. Hence, this hypothesis was not supported. Table 10 shows the impact of SOC ON PTSD

H_3:_ Will SOC influence PTSD among senior citizens people in Ekiti State? The results revealed that SOC had a significant negative impact on PTSD (β = −0.24, t = −3.08, p < 0.05). This finding implies that SOC influences PTSD among older people in Ekiti State. Hence, the hypothesis was supported. Table 10 shows the impact of the PSS on PTSD

H_4_: Will perceived social support influence PTSD among senior citizens in Ekiti State? The results revealed that perceived social support had an insignificant impact on PTSD (β = 0.07, t = 0.98, p_(0.33)_ > 0.05). This finding implies that perceived social support does not influence PTSD among senior citizens in Ekiti State. Hence, this hypothesis was not supported. Table 10 shows the impact of life satisfaction on PTSD incidence

H_5_ hypothesis: Will life satisfaction influence PTSD among senior citizens in Ekiti State? The results revealed that life satisfaction had an insignificant impact on PTSD (β = 0.02, t = 0.16, p_(0.87)_ > 0.05). This finding implies that life satisfaction does not influence PTSD among senior citizens in Ekiti State. Hence, the s was not supported. Moderation analysis: Interaction effect of SOC and on PTSD incidence

H_6_: Will young, old and oldest moderate the indirect relationship between SOC and PTSD? To test the hypothesis that age moderates the indirect relationship between SOC and PTSD, the Hayes process macro was utilized.

**Table 10.**
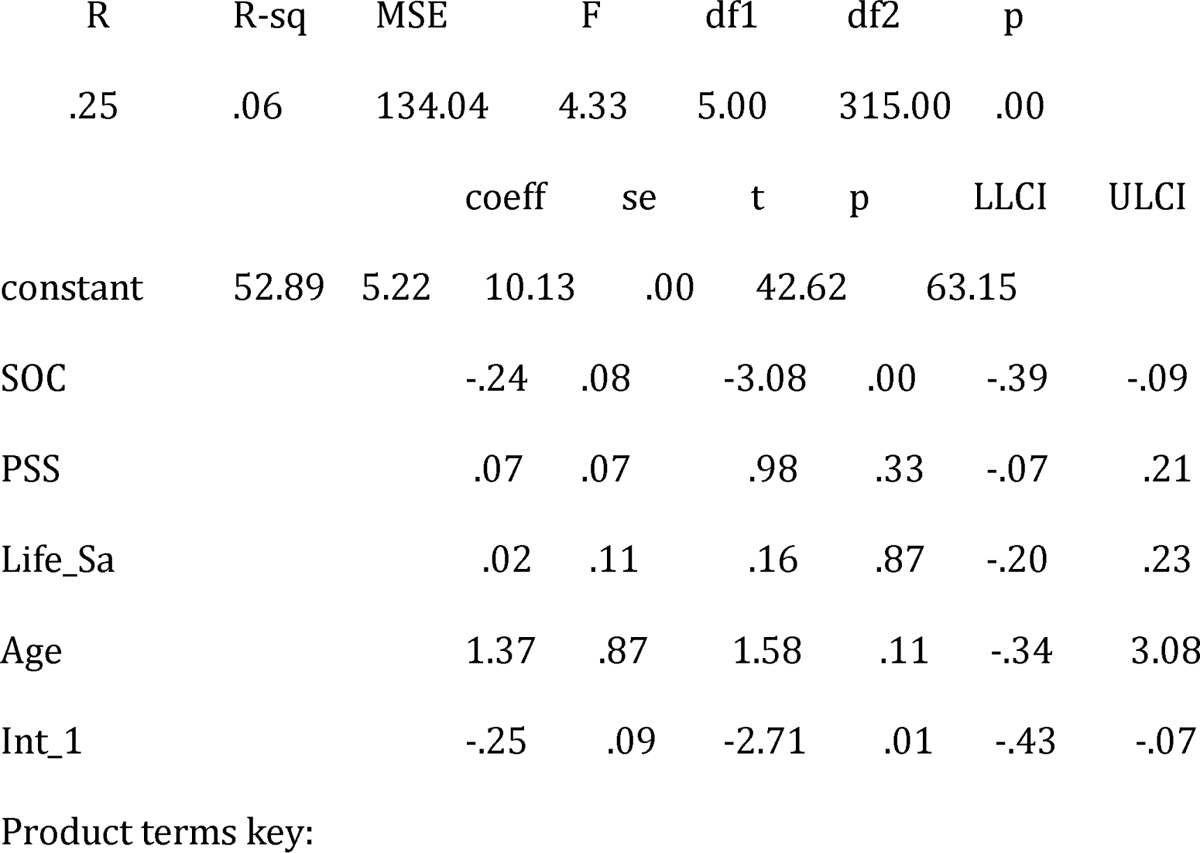
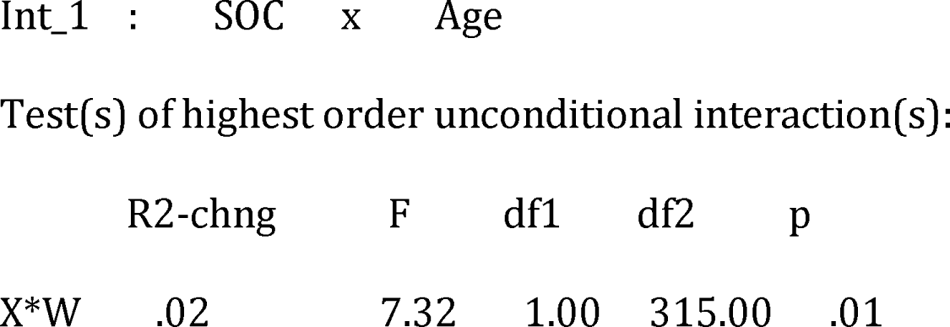
Model Summary.

The moderating interaction effect between age and SOC accounted for a significant amount of variance in PTSD (R^2^ = 0.02, p = 0.05). This finding implies that a 2% change in PTSD can be attributed to the interaction term. The results in Table 3 also reveal a significantly negative moderating role of age in the association between SOC and PTSD (β = −0.25, t = −2.71, p < 0.05). As shown in Table 11, the conditional effects of the focal predictor (SOC) on the moderator (Age) were low (−0.09) and not significant (P_(0.35)_ > 0.05). However, when age was average, its effect increased to (−0.24) and was statistically significant (p < 0.05). Additionally, when age peaked, the effect was high (−0.43) and found to be statistically significant (p < 0.05). This process is also depicted in Fig. 1. This finding suggested that SOC significantly strengthened PTSD symptoms at average (75-54) and high (85 years and older) ages.

**Fig 1:**
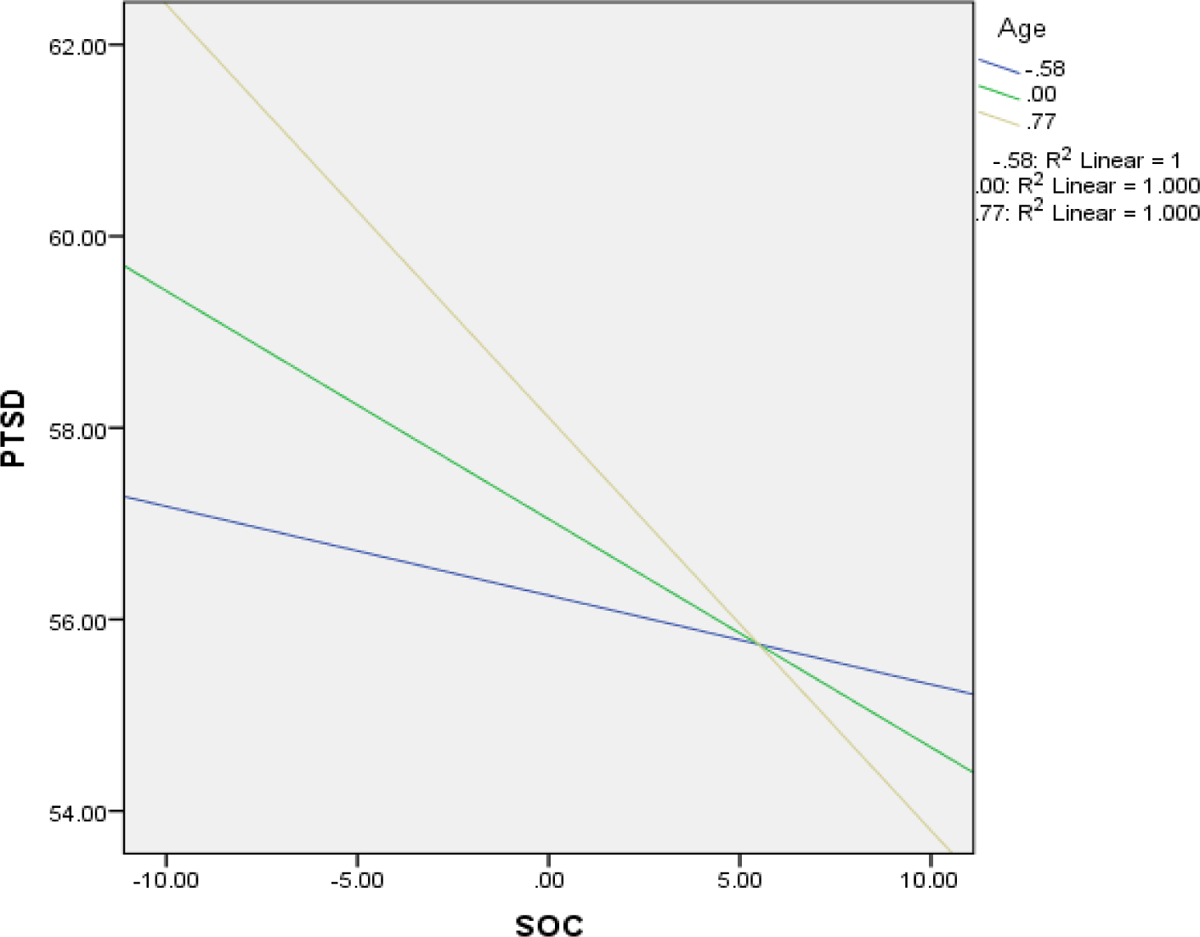
Impact of the SOC and age group on PTSD incidence Mediation analysisi.

**Table 11:**
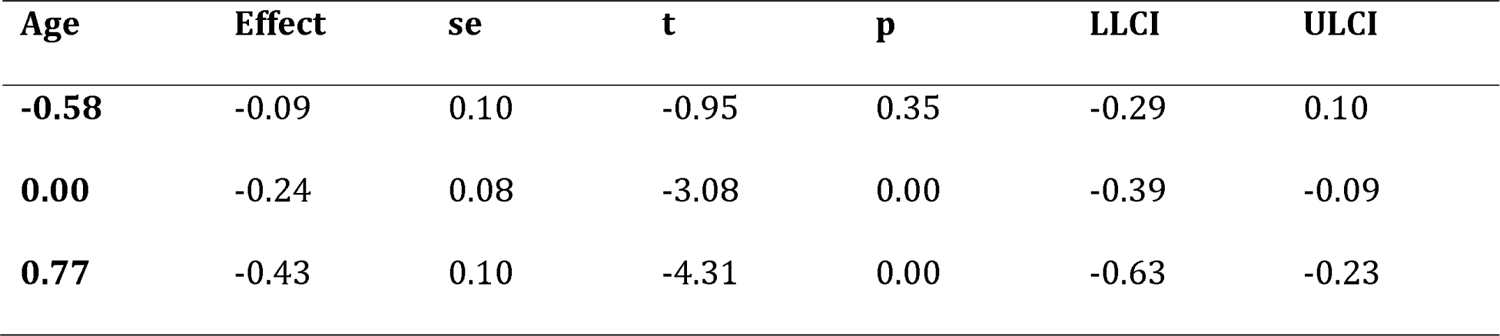
Conditional direct effect(s) of age on the association between SOC and PTSD.

H_7_: SOC strategies will significantly influence PTSD through perceived social support and life satisfaction among older people inEkiti State. Table 12 shows the mediating role of PSS and life satisfaction in the relationship between SOC and PTSD.

**Table 12:**
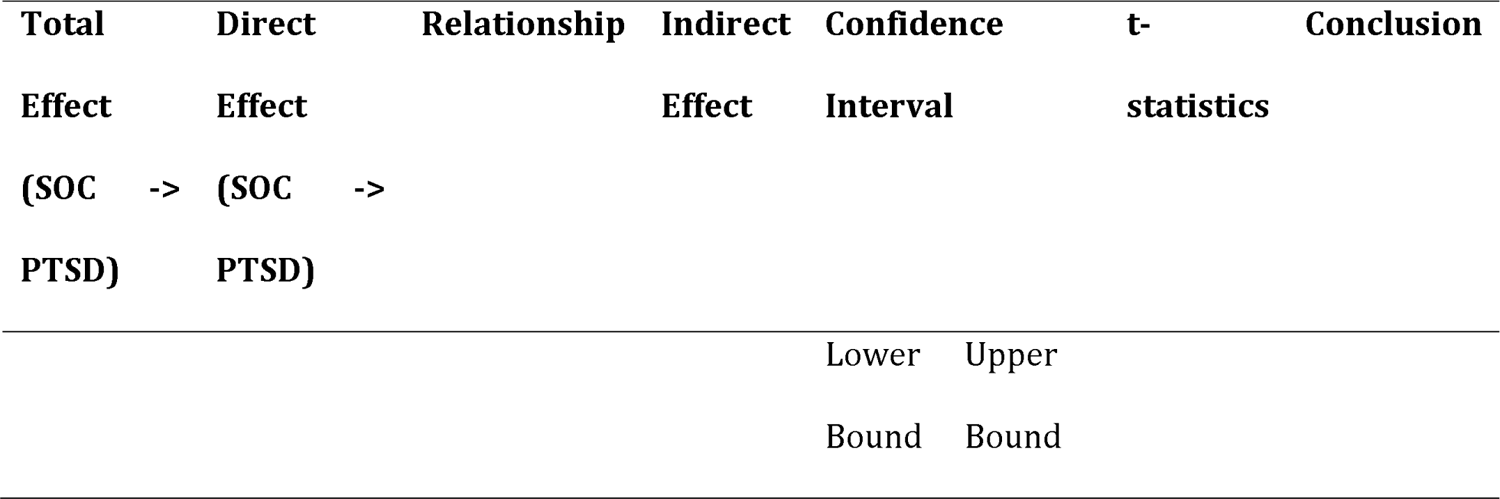
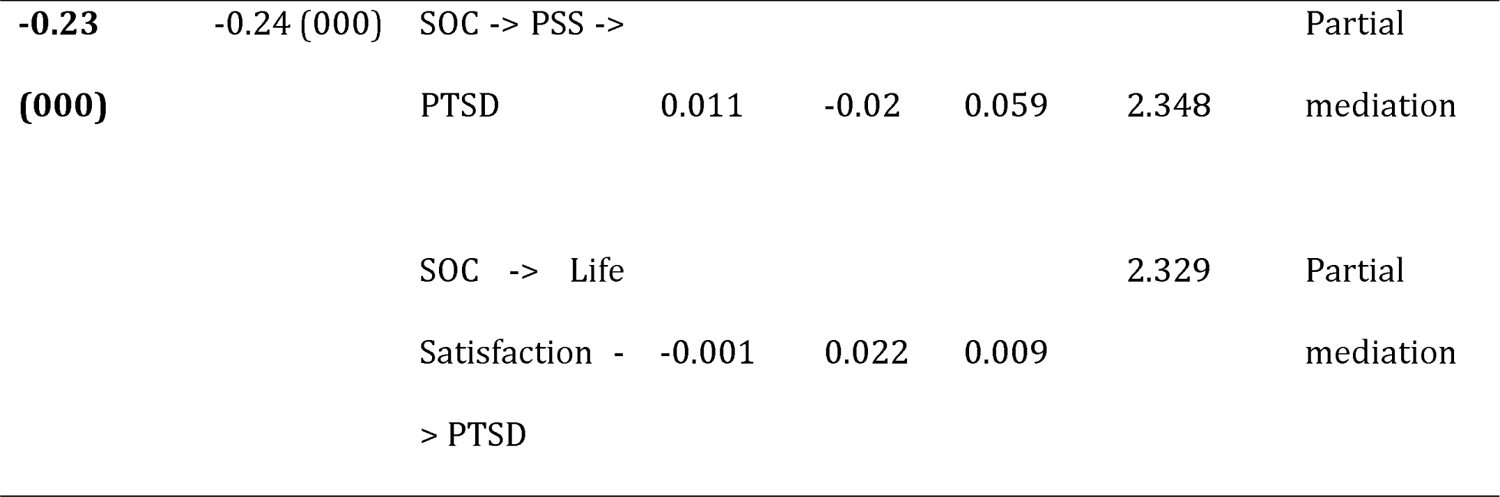
Mediation analysis summary.

## Discussion

Our research objectives were multifaceted: direct influences, moderating and mediating roles, and mechanism-focused exploration. We aimed to evaluate how SOC strategies impact PSS, LS, and PTSD among older Ekiti State individuals. Additionally, we investigated the associations between PSS or LS and PTSD to determine the potential buffering effect of these two variable on the psychological impact trauma. We also examined age groups as potential moderators of these dynamics, deepening our understanding of the topic. Eight hypotheses guided our analysis, including two on moderated mediation, evaluated with the Hayes process macro.

Our findings empirically support the intricate relationships between SOC, PSS, LS, and PTSD within the unique aging context of Ekiti State. The identified moderated mediation effects emphasize the importance of age groups in these relationships. As we delve into the findings, we reveal the interwoven nature of these variables, reflecting the complex tapestry of older individuals’ mental well-being. Our discussion sections further expound on the findings, their implications, and their contributions to the literature on aging, mental health, and the interplay of psychological factors, offering a comprehensive perspective on senior citizens mental well-being in the Ekiti State.

Regarding our first hypothesis, we found that SOC significantly influenced PSS scores with a beta weight of (β = 0.16 t = 2.50, p < 0.05). The finding that SOC significantly influences perceived social support (PSS) is essential and has important implications for understanding how older individuals manage their life challenges regarding the loss of resources in their quests for resource gains. A positive beta weight (β = 0.16) suggested that SOC strategies were directly and positively related to the perception of social support among senior citizens in Ekiti State.

This result indicates that older individuals who engage in effective (SOC) strategies are more likely to perceive more incredible social support from their network of family, friends, and significant others. Individuals who are adept at selecting meaningful relationships, optimizing their interactions with others, and compensating for any limitations may create an environment that fosters a stronger sense of support though their social connections. This finding is in according with the theory of socioemotional selectivity, which states that the life experiences that older people have had over the years can be used to successfully control and avoid potentially harmful experiences by selecting social partners such as family and friends who provide positive emotional feedback and support (Carstensen, 1995; Carstensen et al., 1999). However, these findings disagreed with the previous findings that advanced age and resource deficits may restrict the use of SOC strategies (Baltes & Lang, 1997).

Although this is the first study correlating perceived social support with the SOC, these findings bridge the gaps in the literature gaps. This finding aligns with the theoretical underpinnings of the conservation of resources (COR) theory, which emphasizes the importance of resource accumulation and preservation and that human beings must acquire and conserve both personal strengths and social bonds (Hobfoll et al., 2018). Considering the interplay between perceived social support and SOC strategies, COR theory suggests that individuals who perceive greater social support may have a more flexible resource pool from which to draw (Hobfoll, 1988). This enhanced resource pool could facilitate more effective SOC strategies, as individuals with a strong support network might experience lower stress levels and have more energy and capacity to engage in adaptive coping behaviors. By effectively managing their resources through SOC strategies, older individuals may be better equipped to cultivate and maintain supportive relationships. Furthermore, these findings complement previous research suggesting that SOC involves the active management of resources to adjust to changing circumstances and demands (und and Baltes,2002) and helps older adults use resources effectively and improve life satisfaction (Jopp & Smith, 2006; Freund,2008).

Indeed, the finding that SOC significantly influences PSS underscores the role of active resource management in shaping the social support experiences of older individuals. This insight aligns with the tenets of COR theory. This study offers valuable guidance for potential interventions that seek to augment the well-being of older adults by bolstering their capacity for effective life management strategies (Clark et al., 2012). This finding has far-reaching implications because it encourages the development of tailored interventions, resource allocation, and policies aimed at bolstering SOC strategies to benefit senior citizens’ overall quality of life and mental health.

In our second hypothesis, we examined the causal relationship between SOC and life satisfaction, but we still need to establish this relationship. We aimed to investigate whether emotional engagement with SOC strategies impacts life satisfaction. SOC sees aging as a loss for senior citizens and a gain for young citizens (Feund & Baltes, 1998). The theory posits that one’s resources (mental, physical, and social) are limited to one age. As a result, one tends to become stretch across many domains (Baltes & Baltes, 1998). Therefore, it was unsurprising that SOC strategies failed to impact LS among older people. Although other studies have shown a relationship between SOC and LS among young, old, and old individuals (Jopp & Smith, 2006; Freund, 2008), present study needed to establish SOC’s the influence of SOC on LS. The reason could be that this recent study differs from previous studies in that most studies investigated concurrent relationships without necessarily examining a causal relationship between SOC and life satisfaction. Nevertheless, in this present study, we investigated the causal relationship between SOC and life satisfaction among the senior citizens in Ekiti State.

Again, another possible explanation for the nonsiginificant relationship between SOC and LS could be contextual factors. LS is a multifaceted construct influenced by various factors, including individual characteristics, environmental conditions, and cultural contexts. These contextual factors might either amplify or dampen the direct influence of SOC strategies on LS. For example, Ekiti State is the poorest state in southwestern Nigeria, where the prevalence of underemployment of older adults is 39.1%, with Nigeria’s quality of socioeconomic well-being of senior citizens ranking 86 among 96 countries covering 91% of the world population aged 60 years and older (National Policy on Population for Sustainable Development (NPPSD), 2022; Globawatch, 2022; Sasu, 2022). Therefore, if external stressors such as imitations are partially pronounced, the positive effects of SOC might be constraints resulting in a weaker direct relationship between SOC and LS. This perception is highlighted in conservative resource theory that depleting individuals invest their resources in places and things that are more important for them and their goals (Hobfoll 1989). Therefore, in the context of SOC strategies, older adults in Ekiti State might use these strategies in specific life domains rather than others. For instance, in Africa, older adults prioritize social relationships through social optimization and compensation strategies, which could lead to improved social support. This means that these allocations of resources might come at the cost of other domains, such as personal hobbies, which are associated with life satisfaction.

Moreover, the relationship between LS and SOC is complex, indicating that LS is determined not only determined by SOC strategies but also by psychological, social, and environmental factors. The relationship between LS and SOC might be indirect and complex in Ekiti State. SOC strategies use intermediate variables such as perceived social support, coping strategies, and personal beliefs to impact life satisfaction. This indirect pathway might involve the interplay of various variables that moderate or mediate the relationship between SOC and LS. Thus, the influence of SOC on LS might manifest more strongly through these intermediary means rather than the through direct one-on-one connections. This factor was optimized by context and the absence of direct influence of SOC on L S in our study on contextual factors between SOC and LS. For example, LS in the senior citizens is a positive self-concept that reflects the appropriateness of past goals and outcomes based on current living conditions (Neugarten et al., 1961). The use of an SOC is characterized by goal inappropriateness due to resource depletion and the quest to select replicable relevant resources to meet life demands. Thus, following the literature, the current findings showed that the use of SOC strategies leads to significant variation in life satisfaction (Shirzadifard et al., 2020) and an increase in depressive symptoms (Feund & Baltes, 1998). It has been suggested that SOC is an awareness of loss and a desire for gain (Baltes & Baltes, 2006), which worsens life satisfaction in the present study. For example, studies have shown that sticking too much to the SOC procedures is associated with psychological distress (Yuen & Vogtle, 2016), and those having who have physical and emotional challenges and who align themselves with SOC usage exacerbate distress (Yuen & Vogtle, 2016; Lang et al.’ (2002). Yuen and Vogtle (2016) also explained that clinging to the SOC method was associated with greater physical health stressors. Although (SOC) considers life satisfaction an indicator of psychological adaptation during aging (Baltes, 1997), SOC could not influence life satisfaction in a recent study because LS is an enthusiastic response to life in general and an active acceptance of responsibility for life rather than passive acceptance (Neugarten et al., 1961). While this hypothesis was not supported in the present study, it is critical to recognize thatEkiti State is a resource constraint and that these contextual factors might contribute to life dissatisfaction therefore, SOC alone will not be the only significant predictor of lying satisfaction among senior citizens. Therefore, intervention programs that might boost life satisfaction among senior citizens might need to consider contextual factors other than SOC strategies. This finding stresses the complex factors contributing to life satisfaction among elderly people and highlights the need for a multifaceted approach to improving the well-being of senior citizens in the Ekiti State.

The third hypothesis states that the SOC has a significant influence on PTSD. Our statistical results support this finding (β = −0.24, t = −3.08, p < 0.05). The negative β coefficient (−0.24) suggested an inverse relationship between SOC and PTSD. These findings support our hypothesis that SOC strategies have a meaningful relationship with PTSD. This finding suggested that as SOC strategies increase, the incidence of PTSD tends to decrease among older individuals. This negative relationship suggested that when golden agers embark on higher levels of SOC strategies, their level of PTSD symptoms decreases.

We applied the second foundational principle of conservation of resources (COR) theory to understand how older adults in Ekiti State manage their resources and cope with their challenges. This principle states that people must spend resources to guard against resource loss, recover from losses, and gain resources (Hobfoll, 1998). This principle is particularly pertinent to our investigation, as it underscores the active and continuous nature of resource management in the State. According to the literature (NPPSD, 2022); Globawatch, (2022); Sasu, (2022), the older adults in Ekiti State are resource constrained and often face the dual challenge of resource scarcity and the risk of resource loss.

In this context, they must employ adaptive strategies to safeguard their existing resources, recover from potential losses, and seek opportunities to acquire new resources. This process is closely aligned with the Selection, Optimization, and Compensation (SOC) strategies we examined in our study (Baltes & Lang, 1997; Freund and Baltes (1998; Bates, 1999). Our findings indicate that SOC strategies serve as valuable resource management tools for mitigating PTSD in senior citizens in Ekiti State, allowing them to optimize their available resources, compensate for potential losses, and, in turn, influence their well-being.

Previous studies have shown that SOC management strategies play critical roles in alleviating mental health challenges caused by changes in various life domains during old age (Freund & Baltes, 200a). By understanding how these strategies operate in a resource-limited setting and their implications for mental health outcomes such as (PTSD), we contribute to the growing body of knowledge on the practical applications of COR theory in real-world contexts.

However, our current finding that SOC is a protective tool for mitigating mental health could not be in alignment with the findings of other studies indicating that the use of SOC strategies increases depressive symptoms, drinking, or smoking (Gestsgottir et al., 2010; Gestsgottir et al., 2009) wherein Feund and Baltes (1998), Zerof et al., (1999), and Weiland et al., (2011) added to this complexity of SOC as a trait variable. The intricate relationship between SOC strategies and mental health outcomes, as evidenced by varying research findings, hightlights the multifaceted nature of this construct. While some studies have suggested that using SOC strategies may increase depressive symptoms, drinking, and smoking (Gestsgottir et al., 2010; Gestsgottir et al., 2009) our current research presents contrasting findings. On the basis of our study focused on older individuals in Ekiti State, we discovered that SOC strategies are a protective factor against PTSD among senior citizens in Ekiti State. This discrepancy in results may be attributed to context-specific factors, individual differences, or the specific mental health outcome under investigation.

Moreover, the distinction between SOC as a trait variable (Zerof et al., (1999);Weiland et al. (2011) and its potential state effects further complicates the discussion. Our study’s alignment with the positive impact of SOC-based interventions on well-being and depression severity, as found by Samantaray and Kay (2021), highlights the potential practical implications of our research in the context of older populations facing specific mental health challenges. This discussion stresses the need for a nuanced understanding of the roles of SOC in mental health, considering the various factors that shape its effects.

Nonetheless, acknowledging the complexity of these relationships and considering future research endeavors will deepen our understanding of these dynamics and inform more nuanced approaches to mental health support in similar settings in the Ekiti State or Nigeria. The finding that SOC strategies have a significant negative impact on PTSD among senior citizens in Ekiti State holds promising implications for the psychological well-being of this population. These findings suggest that adopting SOC strategies can serve as a protective factor, potentially reducing the risk of PTSD among senior citizens.

In the fourth hypothesis, our inquiry centers on the potential influence of perceived social support on PTSD among older people. The rationale behind this hypothesis was rooted in the idea that social support serves as a crucial resource for individuals in coping with traumatic experiences (Cohen & Willis, 1985; Thoits, 1986). However, our analysis yielded an unexpected outcome, as the interaction between perceived social support and PTSD symptoms was statistically negligible. This result is evidenced by the nonsignificant beta coefficient (β = 0.07, t = 0.98, p > 0.05). This finding implies that variations in perceived social support among senior citizens did not significantly predict differences in PTSD symptoms. The PSS is often not a stable variable. Its effects on mental health are often based on the psychophysiological circumstances of the participants. According to the PSS literature PSS can also be viewed as variable over time, as it relies mainly on living circumstances, individuals’ recent mental health, and exposure to positive or negative life events [19–21]. Severe stressful event exposure could have a meaningful impact on perceived social support [22]. In the case of Ekiti State, 39.1% of the senior citizens are underemployed with no social security scheme, including health insurance provisions. The State is the poorest in the southwest of Nigeria, where the quality of the socioeconomic well-being of senior citizens is ranked 86 out of 96 nations (NPPSD, 2023; Sasu, 2022; Wan, 2022). Furthermore, individuals coping with severe stressful event exposure might have difficulties engaging in trusting relationships [19, 21]. Indeed, in a state characterized by high poverty rates and underemployment among older individuals, economic stress could impact individuals’ ability to maintain and access social support networks. The strain of economic hardship might lead to diminished social interactions and support availability, aligning with the social erosion or social deterioration theory that PTSD erodes because people with PTSD have an increased tendency to believe that others are dangerous and unsafe (Clapp& Gayle-Beck, 2009), suggesting that social support is influenced by (Wagner et al., 2016; Woodard et al, 2018). These perceptions increase the likelihood of those with PTSD having difficulty establishing trust and, as a result, isolating themselves (Erbes et al., 2012; Wagner et al., 2016).

Our negligible difference between PSS and PTSD agrees with the findings of previous studies suggesting that PTSD symptoms diminish PSS and increase the risk of developing PTSD symptoms (Johansen et al., 2020). Understanding this bidirectional relationship is particularly indispensable for mental health professionals and researchers, especially in a nation where 50% or one in four people suffer from mental health disordersn and without definite budgetary allocation for mental health (The Guardian 4 Dec. 2018; API and EpiAFRIC Jan. 2020; Al Jazeera 2 Oct. 2019; Thisday, 2019-09-05). These findings emphasize the importance of addressing both PTSD symptoms and social support in the treatment and treatment of older persons with PTSD. Effective interventions should aim to not only reduce PTSD symptoms but also strengthen an older individual’s social support network to reverse the sequence and promote recovery.

Indeed, social selection theory illuminates our findings by helping to provide a theoretical framework that helps us explain why symptoms of PTSD often include insecurity, mistrust, avoidance, and social isolation this framework can unravel rejection and gassessing PSS (Mossakowski, 2014) and have a negligible effect on PTSD (Freedman et al., 2015; Wagner et al., 2016; Woodward et al., 2018). Elders with PTSD may find maintaining or establishing social relationships challenging. They may experience rejection or avoidance from others in their social networks. This rejection can lead to a decrease in perceived social support (PSS), which others may not understand or know how to respond to the behaviors and emotions associated with PTSD, making it difficult for individuals with PTSD to receive the support they need.

Furthermore, the socioeconomic challenges older individuals face in Ekiti State and Nigeria could contribute to feelings of isolation and social inequality. These feelings could result from a lack of resources to participate in social activities, reduced access to healthcare and services, or limited opportunities for meaningful engagement. Such experiences of social isolation could exacerbate the development of PTSD symptoms despite the presence of perceived social support. This perception was supported in the literature by the finding that low or adverse social conditions such as social isolation, depressive financial status, social isolation, low socioeconomic status, small social network, or social convoy can lead to diminished mental health and increased psychopathology (Joseph et al., 1997; Mossakowski, 2014). Additionally, PTSD symptoms can destroy interpersonal relationships by increasing antisocial behaviors and damaging social convoys and social resources in close relationships. Indeed, the bidirectional relationship observed between perceived social support (PSS) and (PTSD) in this study calls attention to the complexity of mental health dynamics in resource-constrained environments, such as Ekiti State, Nigeria. PTSD symptoms can diminish PSS due to feelings of insecurity, mistrust, and social isolation (Freedman et al., 2015; Wagner et al., 2016; Woodward et al., 2018) however, recognizing that limited resources and access to mental health services can also impact the availability and effectiveness of PSS is equally desirable.

In hypothesis five, we explore the intricate relationship between life satisfaction and PTSD among senior citizens in the Ekiti State. Recent research has pointed to decreasing life satisfaction among senior citizens in southwestern Nigeria (Adebayo & Arogundarde, 2011; Aina et al., 2023). In contrast, Western societies and Europe have witnessed stable and even peak mental health outcomes among older adults characterized by increased well-being later in life and reduced experiences of negative emotions and daily life stressors (Carstensen et al., 2003; Carstensen et al., 1999; Fung et al., 2008; Gross et al., 1997; Stawski et al., 2008; Charles et al., 2001). Against this backdrop, we hypothesized that life satisfaction will influence PTSD among older individuals in the Ekiti State. This study aims to shed light on this complex relationship and its implications for the mental health and well-being of senior citizens in this specific context. Indeed, our findings suggest that LS has a negligible impact on life satisfaction and PTSD among older people.

Although the result of our study demonstrated by the regression coefficient (β) is 0.02, the t-value is 0.16, and the p-value is 0.87, indicating no significant relationship between life satisfaction and PTSD in our sample of senior citizens in Ekiti State. Despite these findings, the complex interactions between these variables have been evidenced when examining the broader literature. For instance, delay-on-set PTSD among senior citizens has been associated with low life satisfaction and other psychiatric symptoms (Desmarais et al., 2020; Van der Wal, 2020; McCanlies et al., 2014; Feller et al., 2013; Samaranayake & Fernando, 2011). These associations suggest that people with low levels of life satisfaction might be vulnerable to psychiatric conditions such as PTSD, depression, and anxiety, which are comorbid with PTSD (Qureshi et al., 2009). Moreover, our literature highlights that people with moderate or severe PTSD are likely to experience a low level of LF (Karatzias et al., 2013), and meaning in life was associated with decreased PTSD symptoms (Owens et al., 2009). This literature stresses the intricate relationships among life satisfaction, delayed-on-set PTSD, and related psychiatric conditions in older adults, contributing valuable insights into gerontology and mental health. However, further research is needed to understand these dynamic patterns comprehensively and their clinical implications for the well-being of older populations.

Our study’s findings indicate that life satisfaction decreases with age (65-74, 75-84, 85+), with the oldest-old (85+) being the least satisfied. These findings resonate with previous studies in Ekiti State (Adebayo & Arogundarde, 2011; Aina et al., 2023), affirming the consistency of our results with those of the southwestern State of Nigeria. This observation accentuates the challenges posed by resource constraints among these golden-agers. This finding suggested that older individuals, particularly the oldest (85+), encounter unique difficulties related to resource access, healthcare, and social support that contribute to diminishing life satisfaction. This trend is particularly relevant when considering mental health implications, especially in a nation with only 8 Federal and 15 State mental health institutions with limited access to individuals, especially those from rural areas[30-31].

The federal government of Nigeria has remained single digits with budgetary allocations moving from ascending order to descending 5.95% in 2012, 4.4% in 2014, 5.5% in 2015, 4.23% in 2016, 4.16% in 2017, and 3.9% in 2018 while the population is increasing with the worst allocation of budgets to the State and Local Government 29 and 88% respectively (Vanguard News) 2021-08-26), implying that the older population, given their declining life satisfaction and resource limitations, may be more susceptible to mental health challenges such as PTSD. Such findings accentuate the importance of policy initiatives and interventions to enhance the mental well-being of older individuals in resource-constrained settings such as the Ekiti State. The consistency of these findings with those of prior studies in Ekiti State and southwestern Nigeria (Adebayo & Arogundarde, 2011; Aina et al., 2023) strengthens the robustness and validity of the results. This finding suggested that the observed age-related decrease in life satisfaction is not an isolated phenomenon but aligns with broader regional studies.

In hypothesis six, we investigated whether age groups (young, old, old-old, and oldest-old) moderate the relationship between SOC and PTSD. We found that age plays a significant role in moderating the impact of SOC on PTSD incidence. For the young and old individuals (represented by −0.58), the effect of SOC on PTSD was negative (−0.09), suggesting that as SOC strategies increased, there was a tendency for PTSD symptoms to decrease, indicating a potential protective effect of SOC strategies on PTSD but the effect was not statistically significant (p = 0.35). For the “old-old group(represented by 0.00), the effect of SOC on PTSD was negative and statistically significant (−0.24, p = 0.00) for the oldest-old (represented by 0.77), the effect of SOC on PTSD was also negative and statistically significant (−0.43, p = 0.00). These figures show that the relationship between age and SOC score and PTSD symptoms varies across the various age groups, revealing the moderating effects of age group. A negative sign before each effect size (−0.09, −0.24, and-0.43) indicates a negative relationship. That is, as the use of SOC strategies increases, PTSD symptoms tend to decrease. This finding is meaningful because it suggests that SOC strategies may protect against or mitigate PTSD symptoms in all age groups. This finding means that as individuals in these age groups employ SOC strategies more effectively, there is a noticeable reduction in PTSD symptoms. These findings could be valuable for interventions and support programs to improve the mental well-being of senior citizens in Ekiti State.

For young-old, our analysis revealed a conditional direct effect of SOC on PTSD that was negative, indicating that higher levels of SOC were associated with lower PTSD symptoms. Therefore while our study provides some interesting insights into the potential relationship between SOC strategies and PTSD in the young-old group, it is essential to interpret these findings as preliminary. However, further research with a larger sample size or with different methodologies may be needed to conclusively confirm or refute this relationship.

In contrast, the older adults, our results showed a significant and negative conditional direct effect of SOC on PTSD. This finding suggested that among older participants, higher SOC levels are correlated with a statistically significant reduction in PTSD symptoms. The confidence intervals did not include zero, also confirming the strength of this relationship. Similarly, in the oldest-old individuals, we observed a significant and negative conditional direct effect of SOC on PTSD. This finding indicates that SOC plays a meaningful moderating role in mitigating PTSD symptoms among older individuals. The confidence intervals did not include zero, again confirming the strength of this relationship. While the young-old group shows no significant moderation effect, a compelling shift emerges within the old-old and oldest-old groups. Among older individuals aged 75 to 84 (old-old) and those aged 85 and above (oldest-old), SOC exhibited a robust and negative moderation effect. These findings indicate that SOC strategies have a significant protective influence on PTSD symptoms in these older age groups.

This finding helps to answer our general research question of the relationship between (SOC) strategies and (PTSD) among senior citizens in Ekiti State, Nigeria, considering the moderating role of age groups. This study breaks a critical research gap in an area that has received limited attention thereby contributing to the knowledge that the strong negative relationship between SOC and PTSD among young and old people signifies that SOC is a protective variable against mental health or PTSD; hence its presence decreases the level of PTSD. The negative conditional direct effect of SOC on PTSD among younger individuals suggested that SOC strategies are associated with reduced PTSD symptoms in this age group, thereby attesting to previous studies that SOC is a therapeutic variable (Teshale& Lachman, 2016; Samantary & Kay, 2021). These findings provide empirical support for the idea that interventions or therapeutic approaches that promote SOC may be beneficial for reducing the risk or severity of PTSD, especially among younger individuals. This finding implies that interventions focusing on SOC skills could be considered part of a broader therapeutic approach.

Our study depicts SOC as a mechanism older adults use for adapting to stressors, which includes selection, optimization, and compensation. These findings agree with previous studies showing that SOC can be depicted as a model of coping (Lazarus & Folkman, 1984), problem solving (Blanchard-Fields et al., 1997), self-development, and goal adjustment (Brandtsta & Rothermund, 2002).

The findings from our study significantly contribute to our understanding of the moderating role of age in the relationship between SOC and (PTSD), aligning with the literature and theories on age-related coping strategies (Lazarus & Folkman, 1984; Brandsta & Renner, 1990). Our study’s results are consistent with established theories on aging and coping, such as the SOC model proposed by Baltes and Baltes (1990). This model suggests that individuals adapt their coping strategies as they age, shifting from selection and optimization to compensation. Our findings align with this theory, demonstrating that SOC strategies have varying impacts across different age groups (Antonucci et al., 2001; Baltes, 1997; Jopp and Smith, 2006;). Our study showed that SOC use tends to increase from young to middle adulthood and a decrease in SOC use in late adulthood corresponds with the findings of previous studies suggesting that age and resource deficits may limit the use of SOC strategies (Baltes & Lang, 1997) which demonstrated that the use of SOC strategies declines with age. This finding highlights the importance of considering resource availability a critical factor in understanding how age moderates the relationship between SOC and PTSD.

The finding that young individuals have a stronger desire for SOC than older individuals aligns with the findings of previous research showing that SOC increases from young to middle adulthood compared to early and late adulthood (Freund & Baltes, 2002b). However, the study revealed a weak relationship between SOC and PTSD in old-old individuals with effect values for the “old-old” individuals (age group: 0.00) where the effect of SOC on PTSD was −0.24 and for the “oldest-old” individuals (age group: 0.77). When the effect of the SOC on PTSD incidence is −0.43, this weak relationship could not be as farfetched as the state or the nation in general social security schemes that include health insurance care for older people are lacking as life satisfaction decreases as one progresses in age (Adebayo & Arogundarde, 2011), leading to 73.8% of the senior citizens being dissatisfied.

Eighty-one percent of senior citizens depend on their relatives for support (Aina et al., 2023). As resources tend to decline with age, the SOC concentration also declined leading to lower SOC use in late adulthood (Knecht & Freund, 2017), indicating that age can influence preferences for coping strategies. This finding supports the notion that age is a critical factor in shaping an individual’s coping behavior and how they respond to traumatic experiences. Overall, our study’s results provide empirical evidence that age is not a uniform factor in the relationship between SOC and PTSD but rather a nuanced factor. The literature supports the idea that aging is a biological reality and process over which no one has control (Gorman, 2000). According to Shock (2018), when aging occurs in humans, physiological development is typically accompanied by psychological and behavioral changes and other changes involving social factors. The old-old (65-74) and oldest-old (85+) have individuals had accumulated more life experiences and lived through different historical periods than did the young-old individuals. These experiences may include significant life events, challenges, retirement, bereavement, traumatic experiences, and coping strategies that shape their SOC and mental health outcomes. Our findings further suggest that as people age, they might build resilience and adapt over time which might be encompassed in SOC strategies that play a more significant role in mitigating the impact of PTSD and other mental health challenges, suggesting that over time, individuals might become better at selecting, optimizing and compensating to maintain psychological well-being. Top of Form The differing impacts of SOC on PTSD symptoms across age groups highlight the need for age-tailored interventions and support programs for individuals dealing with trauma and stress. This research adds depth to our understanding of coping mechanisms throughout the aging process, offering valuable insights for theoretical frameworks and practical applications in psychology and gerontology.

Furthermore, our findings revealed a significant relationship between SOC and PTSD in the old-old (75-84) and oldest-old (85+) age groups but not in the young-old (65-74) group, which could be interpreted as supporting the decline in SOC (Feund & Baltes, 1998; Baltes, 1998). However, It is essential to acknowledge that our study’s findings may not conclusively settle the debate as they might not cover the complexity of SOC usage, which has been noticed to apply models of coping (Lazarus & Folkman, 1984), problem solving (Blanchard-Fields et al., 1997), self-development, and goal adjustment (Brandtstädter & Rothermund, 2002).

As the first study of its kind in Nigeria and worldwide, our research has broken new ground and laid the foundation for future investigations into SOC and PTSD among older people. This pioneering aspect of this study signifies the importance of exploring this critical area of research on the dimensions of SOC and PTSD in different age groups. Our study contributes to the body of related knowledge by addressing a research gap, providing insights into a unique cultural context, offering age-specific findings, shedding light on mental health in aging populations, highlighting the complexity of SOC, and serving as a foundation for further research. The findings can inform academic discussions and practical interventions related to the mental well-being of senior citizens. Additionally, contextual factors in Ekiti State, Nigeria, might help to explain our findings on how age groups moderate the relationship between SOC and PTSD. According to the Nigerian Population Projection and Demographic Indicator (2020), Ekiti State has the highest total fertility rate (TFR 4.3 in Southwest Nigeria in 2022, the highest dependency ratio 0.8 and the highest level of life expectancy.

level of poverty in southwestern Nigeria in 2022 (Sasu, 2022). These contextual factors might interplay to moderate the effects of SOC on PTSD. For example, these contextual factors suggest that families in Ekiti State have, on average, more children than do those in other states in southwestern Nigeria, which could lead to resource strain. This characteristic can impact the roles and responsibilities of individuals within families, wherein older individuals in the “old-old” and “oldest-old” age groups may have experienced higher levels of caregiving and support responsibilities due to larger family sizes. This caregiving role might influence how individuals use SOC strategies to cope with and mitigate the effects of PTSD and trauma-related stressors. On the other hand, increased family size could be associated with resource strain, which could lead to financial stress, limited personal time for self-care, interpersonal stress, and burnout, potentially influencing how older individuals in these age groups experience and manage PTSD. For older individuals in the “old-old” and “oldest-old” age groups, this could mean they have greater autonomy and independence in their daily lives, allowing them to make choices related to their well-being, including how they use SOC strategies. On the other hand, the high poverty rate of 28% in Ekiti State can significantly impact the stressors and challenges faced by individuals, including older adults, increasing vulnerability to traumatic experiences. It may also affect access to resources for coping and recovery. In the context of our study, older individuals across age groups may have experienced economic hardship due to poverty, influencing their ability to select, optimize, and compensate in their daily lives.

The relatively high life expectancy in Ekiti State suggested that people in this region live longer lives. This longer lifespan can provide more opportunities for older individuals to accumulate life experience and develop coping strategies, including SOC. It may be that older individuals in the “old-old” and “oldest-old” age groups have had more time to refine their use of SOC strategies, potentially making them more effective at dealing with stressors and trauma. Identifying age as a moderator implies that interventions targeting SOC strategies and their impact on PTSD should consider the specific age groups within senior citizens (young-old, old-old and oldest-old). Tailored approaches may be more effective at addressing the mental health needs of individuals in each age group.

For hypothesis seven, we ventured into an uncharted territory by exploring a complex mediation analysis assessing whether the relationship between SOC and PTSD is mediated by perceived social support and life satisfaction among senior citizens in Ekiti State. To provide a comprehensive analysis of our findings, we employed the conservative resource theory and the contextual factors of the Ekiti State. Our study revealed a significant direct effect of SOC on PTSD with an effect size of −0.24 (or −0.240) even when the mediating variables of perceived social support and life satisfaction were present. This regression coefficient (B=-0.24 t=p< 0.05) demonstrated that SOC notably influenced PTSD incidence, indicating that older adults who engaged in adaptive strategies were associated with fewer PTSD symptoms. Our findings accentuate the critical role of SOC in regulating the health outcome of the older individuals in Ekiti State, most significantly PTSD. Although previous studies have examined coping strategies and their effect on PTSD (Ogle et al., 2013; McLean & Foa, 2017), our study represents a pioneer in specifically probing the influence of SOC strategies on PTSD. This novel exploration highlights the adaptability of SOC strategies and their significance in resource allocation for mitigating PTSD symptoms among elderly people.

The total effect of (−0.23 or −0.230) and the direct effect of (−0.24 or −0.240) present the primary relationship between SOC and PTSD both with and without the mediating values of perceived social support and life satisfaction, signifying that the influence of SOC on PTSD remains substantial. These effect sizes aligns with the core concept of SOC, which posits that individuals try to maintain balance in their lives by seeking the best way to pay off for biopsychosocial and cognitive impairment and to become more proficient in activities they know how to do best (Baltes et al., 2006). These findings suggest that the SOC theory is a life management model for adjusting to physical and behavioral changes (Fruend & Baltes, 200a). On the other hand, most significantly, the negative total effect (−0.23 or −0.230) of SOC within the resource constraints (high dependency ratio, high fertility rate, high poverty level) environment of the Ekiti State proposes that effective use of SOC strategies by older could help to mitigate resource stressors on PTSD and further justify the conservation of resources that individuals often face challenges related to potential or actual losses; additionally, these individuals are motivated to retain, gain and protect valuable resources for anticipated future needs (Hobfoll, 1988; 1989).

A relevant study conducted by support Basharpoor & Eyni (2021) suggested that senior citizens’ level of PTSD depends on life satisfaction and perceived social support, in connection with other findings that perceived social support and life satisfaction play essential roles in the physical and mental balance of senior citizens (Borg et al., 2006, Harandi, el al., 2017). The studies by (Basharpoor and Eyni (2021 and Borg et al. 2005 and Harandi et al., 2017) indicate that perceived social support enables senior citizens to cope with PTSD and other mental health challenges. Studies by Basharpoor and Eyni (2021) and Borg et al. (2005 and Harandi et al., 2017) further indicated that life satisfaction plays a vital role in PTSD and mental health. The findings of Basharpoor & Eyni (2021) and Harandi et al., (2017) suggest that perceived social support and life satisfaction might serve as critical explanatory mechanisms for the relationship between SOC and PTSD among senior citizens in the Ekiti State. These studies are crucial to our research because they support the examination how SOC strategies might influence PSS and LS and ultimately affect the likelihood of PTSD among the older adults in the Ekiti State.

Based on our study objectives of investigating the relationship between SOC and PTSD considering the explanatory roles of perceived social support and life satisfaction among older people and in the Ekiti State, we unearth the substantial mediating role of perceived social support and life satisfaction in explaining the association between SOC strategies and PTSD which has not been done before in a previous study. These results resonate with established theories that PSSs are primary interpersonal resources that are critical for coping with stress and are associated with psychological distress (Norris & Kaniasty, 1996) and the protective role of life satisfaction in health protection dampening the risk of chronic physical illness and promoting longevity promoting a more active lifestyle and motivating individuals toward self-care (Carver et al., 2010). Importantly, our study illuminates SOC strategies as dynamic mechanisms through which senior citizens manage their resources to augment social support networks and enhance life satisfaction.

Indeed, in our mediation analysis, the indirect effect of SOC > PSS > PTSD was 0.011, indicating the extent of the mediated relationship through perceived social support and indirect effects. Additionally, we identified an indirect effect of SOC > Life satisfaction> PTSD, amounting to −0.001, signifying the extent of the mediated relationship through life satisfaction. SOC > Life satisfaction > PTSD of −0.001, indicating the extent to which the relationship was mediated through life satisfaction. These indirect effects help us understand the magnitude and direction of the relationship between PSS and LS. The positive and negative mediation signs of the indirect effects reveal different relationships within our mediation model. In essence, a positive relationship means that as senior citizens of Ekiti State employ SOC strategies (such as selecting and optimizing their social support networks) they are more likely to perceive more incredible social support. This finding aligns with the idea that SOC strategies can lead to a subjective perception of increased social support among senior citizens (Weiland, 2011). Additionally, these findings emphasize the role of SOC strategies in shaping individuals’ perception of the support available to them, which, in turn, can influence various aspects of their well-being (Ejigu & Tessema, 2017), including mental health outcomes such as PTSD. However, as indicated in the current study, this increase in PSS score could be associated with a greater likelihood of experiencing PTSD. This finding could be because Ekiti State is the fastest-growing State in the Southwest of Nigeria, with a large family size, high dependency ratio, and the poorest State in Southwest Nigeria (NNPCO, 2020; 2022; Sasu, 2022). These contextual factors could lead to poor perceived social support because of an increasing financial burden on caregivers, potentially increasing stress levels, and the risk of developing PTSD, implying that PSS could adversely influence the mental health of the senior citizen population. These findings are consistent with previous studies showing that poor PSSs are an essential risk vulnerability factor for the development and maintenance of PTSD symptoms (Brewin et al., 2000; Ehlers & Clark, 2000; Holeva et al., 2001; Ozer et al., 2003; Robinaugh et al., 2011) which often leads to life dissatisfaction (Weyers, 2008).

Contextually, Ekiti State has been characterized by a diminishing level of life satisfaction (Adebayo & Arogundarde, 2011) as 73.8% of the senior citizens are dissatisfied with life, while 81% of them are dependent on their relationships for support (Aina et al., 2023). However, our negative indirect effect has demonstrated that senior citizens who use SOC strategies to optimize and allocate their resources toward their chosen goals and activities are more likely to experience increased life satisfaction. This increased life satisfaction is associated with a lower likelihood of experiencing PTSD, indicating a potential protective effect of higher life satisfaction against PTSD.

Considering these findings, it becomes evident that the senior citizens in Ekiti State face uncommon resource challenges that demand targeted interventions and support programs. The conservative resources theory offers insights through which we can view these challenges, emphasizing the significance of resource acquisition, protection, and enhancement for the well-being of senior citizens. Given the contextual factors, such as the high dependency on social support and low levels of life satisfaction among senior citizens in Ekiti State, our study highlights the urgency of designing interventions that bolster social support networks and empower senior citizens to manage their resources effectively through SOC strategies. By doing so, we can strive to enhance the mental health needs of these vulnerable senior citizens and contribute to the broader field of gerontology.

In hypothesis eight, we investigated the variation in life satisfaction, perceived social support, PTSD and SOC among the three age groups. Our finding revealed that between the ages 65-74 years, LS and PSS had weak variation that was not significant (p = 0.473). Between the ages 75 and 84 years, PSS and LS had a moderate positive correlation (0.241) that was significant (p = 0.037) implying that an increase in PSS leads to a corresponding increase in LS. Moreover, between the ages of 85+ years, we found a weak negative correlation (0.096) between LS and PSS but this correlation was also not significant (p = 0.480). While this study revealed that there was a positive correlation between LS and PSS among older person aged 75-84 years which has been supported in previous studies (Chruœciel, et al., 2018; Ghimire, et al., 2018; Prakash & Srivastava 2020), there was a negative correlation between PSS and LS among seniors aged 85 years and older suggesting that those with high PSS scores tend to have low LS. While this relationship was not significant, contextual factors in Ekiti State such as large family size, poverty and unemployment of caregivers as well as the psychophysiological characteristics of those aged 85+ might have influenced this finding. The quantity (among the support, frequency of support) and quality (how satisfied one is with the support, duration of support) of PSS are often associated with life satisfaction (Ward, 1985). In fact, PSSs can also be considered recidivists over time and often depend on one’s living circumstances, individuals one’s current mental illness, and one’s exposure to positive or negative life events (Thoresen et al, 2019; Kaniasty K, Norris, 2008).

On the other hand, in the 65-74 age group, there was a positive correlation between LS and PTSD (0.795) although this correlation was not significant at 0.05 which is consistent with the findings of previous studies (Brandstadter & Greve, 1994; Krauss-Whitbourne & Sneed, 2002). Regardless of natural age-related declines in health, income, and physical or cognitive abilities older adults are often satisfied due to their improved ability to engage and proactively regulate emotions (Lawton et al., 1992). Our finding of a positive correlation in the 65-74 age groups could be interpreted through the lens of Socioemotional selectivity theory of (Carstensen, 1995) which suggests that, despite the challenges associated with PTSD, older adults in this age range may be prioritizing positive emotional experiences and regulating their emotions effectively. Therefore therapeutic approaches that emphasize positive emotional experiences and coping strategies could be tailored to this age group However, in the 75-84 age group, there was no statistically significant correlation (−0.117) between life satisfaction and PTSD among individuals in this age group. This negative correlation suggests that a weak tendency for lower life satisfaction to be associated with higher PTSD symptoms (Hegel et al., 2005) which could be correlated with diminished interest in customary activities has also been noted (Avrill & Beck, 2000). However, this association was not statistically significant suggesting that further studies are needed to delve deeper into this complex intricacy. Again in the 85 + age group, there was no statistically significant correlation (0.113) between life satisfaction and PTSD among individuals in this age group. The positive correlation suggested that a weak tendency for higher life satisfaction was associated with slightly greater PTSD symptoms, but this association was not statistically significant.

The finding of a nonsignificant positive correlation (0.795) between life satisfaction and PTSD symptoms in individuals aged 65-74 and 85 years (0.795; 0.113) prompts a nuanced discussion. This result may be interpreted through the lens of socioemotional selectivity theory, which suggests that older individuals, drawing on a lifetime of experiences, become more discerning in their social interactions (Carstensen, 1995; Carstensen et al., 1999). Despite the potential challenges posed by PTSD symptoms, these age groups strategically select positive social partners, prioritize emotional well-being and forster high-quality relationships. Regardless of natural age-related declines in health, income, and physical or cognitive abilities, senior citizens may remain satisfied because they make an accommodative shift that aligns their aims and goals with what is achievable (Brandstadter & Greve, 1994; Krauss-Whitbourne & Sneed, 2002) The observed nonsignificant correlation may reflect the success of adaptive strategies employed by older individuals, who, through engagement with positive emotions and selective social connections, maintain a higher level of life satisfaction even in the face of PTSD symptoms despite age-related declines in health, income, and physical or cognitive abilities, may remain satisfied because they make an accommodative shift that aligns their aims and goals with what is achievable (Brandstadter & Greve, 1994; Krauss-Whitbourne & Sneed, 2002). This aligns with broader trends where older adults, on average, report greater life satisfaction and exhibit greater resilience, emphasizing the importance of considering the quality and purpose of social relationships in understanding well-being.

Furthermore, these findings resonate with the literature indicating that older adults often exhibit heightened levels of life satisfaction compared to their younger counterparts (George, 2010; Martins et al., 2018). Greater attention and recall of positive stimuli at older ages contribute to the cultivation of positive memories, potentially acting as a buffer against the negative impact of PTSD symptoms. The nonsignificant correlation underscores the complexity of the relationship between mental health and subjective well-being in older age, emphasizing the need for a holistic understanding that considers individual coping mechanisms, adaptive strategies, and the unique socioemotional landscape of older adulthood. Future research could further explore the mechanisms behind the observed correlation, exploring the role of specific adaptive strategies, social support structures, and individual resilience factors that contribute to the maintenance of life satisfaction among older individuals with PTSD symptoms.

Additionally, at age 65-74, there is a weak negative correlation (−0.076, p = 0.295) between selection optimization compensation (SOC) and life satisfaction (0.076) at age 75-84 there is a negative relationship (−0.083, p= 0.477) between SOC and LS. Furthermore at age 85 above, there are a positive relationship (0.051, p=0.706) between SOC and life satisfaction. This study failed to reveal a significant relationship between these age groups, but the weak negative relationships between the ages 65-74 adn75-84 years old signify that when senior citizens are satisfied and perceived their social environment to be supportive they might relax through the use of compulsory coping strategies such as SOC. While studies have shown that using SOC could help individuals improve life satisfaction (Freund 2008), our study although the first to break the ground between SOC and LS demonstrates that life satisfaction would have a diminishing effect on SOC usage among seniors between the ages of 65 and 84. This finding was confirmed in a previous study when older adults with more significant disability made greater use of SOC strategies (Yuen & Vogtle, 2016). On the other hand, the positive relationship between life satisfaction and the SOC of the seniors in 85 above aligns with the perceptions that SOC could be a model of coping (Lazarus & Folkman, 1984), problemsolving (Blanchard-Fields et al., 1997), self-development, and goal adjustment (Brandtstadter & Rothermund, 2002).

However, the weaker mean scores indicate that, on average, the oldest-old group may not engage in SOC strategies as extensively as younger age groups. Although the oldest-old reported the weakest mean scores SOC or strategies in the present study, Jopp & Smith (2006) reported the same weaker mean scores. Considering the current finding that perceived social support and life satisfaction are associated with reduced use of SOC strategies, interventions aimed at enhancing perceived social support and life satisfaction may need to consider how this could impact individuals’ adaptive coping mechanisms.

On the other hand, at 65-74 years there was a weak positive correlation (0.104, p=0.015) between SOC and perceived social support, while at 75-84 year, there was a negative correlation (−0.0404, p=0.00) between SOC and PSS at 85 years above, there was a strong positive correlation (0.539, p=0.00) between SOC and PSS among the senior citizens of the Ekiti State. The weak but statistically significant positive correlation between age groups 65 and74 years indicates that higher perceived social support is associated with increased engagement in SOC strategies.

This finding aligns with the concepts that aging is a process of self-development problem solving and adjustment (Blanchard-Fields et al., 1997; Brandtstädter & Rothermund, 2002). Hence, individuals in this age group are still more actively engaged in adapting to aging challenges, and higher perceived social support could positively influence their use of SOC strategies. While the current study is the first to link SOC with perceived social support, we propose that the relationship between SOC and PSS aligns with the COR theory of Hobfoll (1988) which posits that individuals who perceive higher social support may have a more incredible resource pool from which to draw (Hobfoll, 1988). This enhanced resource pool could facilitate more effective SOC strategies, as individuals with a strong support network might experience lower stress levels and have more energy and capacity to engage in adaptive coping behaviors. As Brandstadter & Rothermund, 2002) noted, the early stages of late adulthood might involve a proactive approach to maintaining well-being and ajusting to life challenges as SOC usage has no limit across age groups since SOC behaviors are used from early to late adulthood (Baltes, 1998).

Additionally, at 75-84 years, there was a slight shift in adaptive strategies hence, there was a negative correlation (−0.0404, p=0.00) between SOC and PSS suggesting that social support is associated with reduced life management strategies among this age group. The negative correlation between SOC and PSS score (−0.0404) suggested that as perceived social support increases, the utilization of life management strategies (SOCs) decreases. This finding aligns with the idea of a buffering effect, where the presence of perceived social support is associated with a reduction in the need for specific coping mechanisms (Cohen & Wills,1985). These findings suggest that heightened social support may contribute to a reduction in the reliance on specific life management strategies, indicating that the protective role of social support in buffering the impact of aging-related challenges is in line with the COR theory that individuals who perceive higher social support may have a more incredible resource pool from which to draw (Hobfoll, 1988).

Again in the 85+ age group, there was a moderate to strong positive correlation (r = 0.539, p = 0.000) between perceived social support and SOC. This finding implies that individuals in this oldest age group who perceive higher levels of social support are more likely to engage in selection optimization compensation (SOC) strategies. This finding showed that above age 85 perceived social support increases and the use of SOC methods for life management increases. While there is no study linking SOC and PSS, these recent findings align with the strength and vulnerability integration (SAVI) model which posits that as people age, they hierarchically engage themselves in thoughts and behaviors that lead to more positive emotional experiences than younger groups (arstensen and Mikels 2005).The positive relationship between PSS and SOC among the oldest-old people is associated with strength and vulnerability integration (SAVI) signifying that despite potential vulnerabilities associated with aging, individuals can leverage social support as a source of strength to enhance their adaptive coping strategies. These findings further explain whyolder adults tend to have higher levels of engagement with positive emotions than younger adults do, meaning that older adults pay more attention to and remember positive stimuli better than their younger counterparts do (Martins et al., 2018).

The links among PSS, SOC, and SAVI underscores the dynamic interplay between social support, adaptive coping, and nuanced experiences of strength and vulnerability in the aging process. The study’s findings contribute to a deeper understanding of how social connections influence the utilization of adaptive strategies among the oldest-old individuals, aligning with the core principles of the SAVI framework. These findings have profound implications for understanding the adaptive strategies employed by individuals in different stages of late adulthood. The age-specific variations underscore the need for tailored interventions and support systems. For the 65-74 age group, fostering social support may contribute positively to the utilization of adaptive strategies. In contrast, for the 75-84 age group, the negative correlation suggested that reliance on external support might diminish the need for certain coping strategies. The oldest-old group (85+), which displayed a positive correlation, emphasizes the potential significance of social support in sustaining adaptive behaviors. These insights highlight the importance of recognizing the diversity within the aging population and tailoring interventions that consider age-specific dynamics in promoting well-being and adaptive functioning.

### IMPLICATIONS OF THE STUDY

One climactic finding of this study is the role of SOC strategies as protective measures against PTSD among senior citizens in Ekiti State. The positive relationship observed, where an increase in SOC strategies corresponds to a decrease in PTSD symptoms, emphasizes the psychotherapeutic potential of SOC strategies in enhancing mental well-being in this population. These findings hold significant implications for developing targeted interventions and programs to bolster older adults’ psychological resilience, offering a promising avenue for mitigating the burden of PTSD in similar contexts. Furthermore, this was the first study to establish a significant negative association between SOC strategies and PTSD among elderly people. As the first of its kind in this context, our research extends the current knowledge base and opens new pathways for future investigations into the interaction between SOC strategies and mental health outcomes, offering fresh insights and avenues for developing targeted interventions.

Additionally, our study used age groups as a moderator, implying that interventions targeting SOC strategies and their impact on PTSD should consider the specific age groups within senior citizens (young-old, old-old and oldest-old). Tailored approaches may be more effective at addressing the mental health needs of individuals in each age group.

For instance, intervention programs for young-old individuals (65-74) for coping with stress and trauma, such as stress management, mindfulness practices, and emotional regulation skills, as well as promoting opportunities for social and community involvement techniques, could be employed to enhance the psychological well-being of the golden aged people. For the old-old (75-84), encouraging reminiscence therapy, where individuals share and reflect on their life experiences, could help older people make sense of their past and promote a sense of meaning and coherence, aligning with the compensation aspect of SOC. Similarly, focusing on health and wellness programs to address physical and mental health issues common in this age group could boost older persons’ psychological self-worth, thereby supporting their physical well-being and positively impacting their overall SOC.

For the oldest-old (85+), providing adaptive strategies to help individuals in this age group cope with the challenges of aging, including physical limitations and health issues, aligns with the selection aspect of SOC. Moreover, offering holistic care that takes into account the physical, emotional, and social well-being of this age group while employing comprehensive healthcare services that could support the overall SOC of the “oldest-old and Connect individuals in this age group with community resources and services, such as home healthcare and senior centers, to address their unique needs and services Again, the interaction effect indicates that the relationship between SOC and PTSD varies across different age groups. This complexity emphasizes the importance of considering age as a multifaceted variable when designing interventions and support systems for senior citizens with PTSD.Given the aging population and the decline in life satisfaction among older adults, with the oldest-old (85+) reporting the lowest levels of satisfaction, there is a pressing need for policy considerations that should prioritize the well-being of this demographic, recognizing age as a crucial determinant of well-being in this population.

Advocating for increased budgetary allocations to healthcare, especially in states such as Ekiti State, becomes crucial for addressing the specific needs of older individuals. Considering that SOC strategies impact PSS among older people, promoting and fostering SOC strategies among senior citizens in Ekiti State can positively impact their perceived social support. These findings can inspire the development of interventions and programs to enhance social support systems for older adults, potentially leading to improved well-being and mental health outcomes. In conjunction, policymakers and healthcare providers can use findings to tailor support programs designed to help seniors develop and implement SOC strategies. These programs may include training, counseling, or community-based initiatives that empower seniors to optimize their social resources effectively.

Finally, scholars should consider that in some cases, the total effect (−0.23 (p < 0.001) being lower than the direct effect (−0.24 (p < 0.001) may align with the unique contextual factors of our study. These contextual influences, such as the specific characteristics of the Ekiti State and its senior citizens population, can shape how SOC strategies, PTSD, and mediating variables interact. It is essential to recognize that research findings are not one-size-fits-all and should be interpreted within the study’s context. Therefore, the observed pattern is theoretically valid when considering the complex interplay of factors in the context of this study.

### LIMITATIONS AND RECOMMENDATIONS FOR FURTHER STUDIES

While this study made some significant contributions to the body of related knowledge, it also has several limitations. One of the limitations of this current study is that it examines composite scores of SOC, PTSD, (PSS), and LF, treating these variables as general constructs. However, these variables may exhibit domain-specific nuances that our study must fully capture. Future research should consider exploring the domain-specific aspects of SOC, PTSD, PSS, and LF. This approach can involve a more granular examination of the subcomponents or facets within each construct to gain a deeper understanding of their relationships and potential moderating/mediating factors. Additionally, employing domain-specific measurement instruments or qualitative methods may provide richer insights into the intricacies of these constructs within the context of the study population. By addressing this limitation, researchers can refine their understanding of these variables and enhance the validity and specificity of their findings in future studies.

The second limitation of this study is the absence of sociodemographic variables in our analysis. While we thoroughly examined the relationships between SOC strategies, PSS, LS, and PTSD incidence, the influence of soiodemogragphic factors such as sex, education, socioeconomic status, and marital status was not explored. These variables can significantly contribute to the outcomes under investigation, and their inclusion in future research could provide a more comprehensive understanding of the complexities involved. Future research should build upon this study’s findings and address the limitations mentioned above by incorporating sociodemographic variables into the analysis. Investigating how age, sex, educational background, socioeconomic status, and other demographic factors interact with SOC strategies, PSS, LS, and PTSD among older adults in Ekiti State could yield valuable insights. This approach allows for a more nuanced examination of the various factors influencing the psychological well-being of the senior citizens. The findings could guide the development of more targeted interventions and support programs.

The third limitation of this study could be its quantitative nature. While this study employed quantitative methods to examine the relationships between SOC, PSS, LS, and PTSD among senior citizens in Ekiti State future research could benefit from a more diverse methodological approach such as case studies, in-depth interviews, or focus group discussions, to capture older adults’ lived experiences and narratives. Such qualitative studies could help individuals better understand the psychosocial dynamics and contextual factors influencing their well-being. Additionally, combining quantitative and qualitative data, mixed-methods research could offer a more holistic perspective on the complex interplay of variables affecting senior citizens’ mental health and life satisfaction in the Ekiti State.

The fourth limitation of this study is that the current research was conducted within the specific context of Ekiti State, Nigeria, which is characterized by unique socioeconomic conditions, including poverty, a high dependency ratio among older adults, elevated life expectancy, and a significant level of underemployment among senior citizens. These contextual factors may limit the generalizability of the study’s findings to other regions or populations with different socioeconomic and cultural backgrounds. Consequently, the results should be interpreted within the context of Ekiti State, and caution should be exercised when extrapolating the findings to broader populations. To address these limitations, future research should explore the influence of socioeconomic and cultural factors on the relationships between SOC strategies perceived social support life satisfaction and PTSD among senior citizens in other states in Nigeria or other nations and settings. Comparative studies across different Nigerian states or countries with diverse socioeconomic conditions could provide insights into how these contextual factors impact the well-being of older people. Additionally, investigations into the effectiveness of tailored interventions and policies designed to address the unique challenges faced by senior citizens in resource-constrained settings such as the Ekiti State, may offer valuable insights for improving the mental health conditions and quality of life of vulnerable people.

## CONCLUSIONS

Examining the mediating roles of LS and PSS in SOC and PTSD and the moderating roles of age groups is paramount in mitigating the effects of PTSD among the older people of Ekiti. The present study aimed to examine the influence of SOC on PSS, LS, and PTSD among senior citizens in Ekiti State. This study also aimed to investigate the influence of PSS and LS on PTSD. Most importantly, this study investigated the moderating effects of age groups (young, old, and oldest-old) on SOC and PTSD while also examining the mediating effects of LS and PSS on SOC and PTSD among the senior citizens in the Ekiti State in Nigeria. Conservation of Resources (COR) theory, a well-established framework in psychology, guided our exploration of how resource management strategies, such as SOC, interact with mental health outcomes, particularly PTSD. It is essential to note that this study is among the pioneering research efforts to investigate the relationship between SOC and PTSD in any context, making our findings particularly novel and groundbreaking.

Indeed, our moderated mediation analysis showed that age group negatively moderated the relationship between SOC and PTSD. The higher the age group is the greater the level of moderation, and the lower the age group is the lower the moderation level. The findings revealed distinct age-related patterns, with the oldest old exhibiting the most robust relationship between SOC strategies and PTSD. This negative moderation of our findings implies that SOC is not only a protective life management strategy but also a therapeutic variable that alleviates and weakens mental health effects among older people such as PTSD. This finding aligns with the core tenets of COR theory, which emphasizes the importance of resource management strategies in safeguarding mental health. By revealing that higher utilization of SOC strategies is correlated with lower levels of PTSD among older individuals, our study underscores the relevance of this theory in the context of aging and mental well-being.

This finding offers a potential path for addressing and mitigating the effects of mental health challenges among senior citizens in the challenging state of Ekiti. In addition, our study introduces a nuanced perspective by exploring how age groups (young, old, and oldes) moderate the relationship between SOC and PTSD, help extend the applicability of COR theory, and emphasize the need for tailored interventions that account for the unique needs of senior citizens at different stages of aging. In this in our study’s contributions to the broader geropsychology field and to COR theory highlight the importance of considering resource management strategies, such as SOC, in addressing mental health challenges among senior citizens in resource-constrained environments in the Ekiti State.

Furthermore, in our mediation analysis, we revealed negative total and direct effects of SOC on PTSD, implying that higher utilization of SOC strategies was associated with low levels of PTSD among older persons. The indirect effects of the partial mediation of perceived social support and life satisfaction suggest that part of the relationship between SOC and PTSD can be explained by the presence or activities related to life satisfaction and perceived social support, indicating that these factors play roles in alleviating mental health burdens among the older persons in the Ekiti State. While our study unequivocally demonstrated that SOC strategies play a pivotal role in mitigating PTSD symptoms among older individuals in Ekiti State, these finding underscore the importance of fostering these strategies as potential interventions for enhancing the mental health and well-being of the older individuals.

Contrary to our initial hypotheses, we found no direct influence of SOC strategies on life satisfaction, nor did we find a direct relationship between life satisfaction and perceived social support for PTSD among older adults. However, it is crucial to acknowledge the nuanced nature of life satisfaction as life satisfaction partially mediates the relationship between SOC strategies and PTSD. However, our study reveals a notable trench where life satisfaction decreases as age increases among senior citizens in Ekiti State. It is important to consider the complex interactions of factors contributing to this observation. One plausible explanation could be the accumulation of life stressors and health-related challenges as individuals age. Senior citizens may face various physical, cognitive, and social changes that impact their overall life satisfaction.

Moreover, the socioeconomic conditions and limited access to healthcare resources in Ekiti State may play a substantial role. High levels of poverty, a high dependency rate on family members, and limited employment opportunities for senior citizens can lead to financial and emotional stress, which, in turn, may negatively influence life satisfaction. The absence of dedicated mental health resources and support programs within the state could also contribute to these findings. This might lead to an underreporting of life satisfaction and may affect perceived social support, thereby influencing overall life satisfaction among older people. Future research could delve deeper into these complexities to better understand the specific elements contributing to the observed decline in life satisfaction among senior citizens in Ekiti State. This complexity emphasizes the need for a comprehensive understanding of the factors contributing to life satisfaction. On the other hand, although perceived social support did not directly influence PTSD incidence, it served as a critical mediator between SOC strategies and PTSD. This underscores the subjective and cognitive aspects of perceived social support, highlighting its significance for the mental well-being of older individuals.

Our study not only advances our understanding of the mental health demands faced by senior citizens in resource-constrained settings such as the Ekiti State but also makes meaningful contributions to geronpsychology. Specifically, our findings align with the conservation of resources (COR) theory, a well-established framework in psychology. This theory posits that individuals strive to acquire, maintain, and protect resources essential for their well-being and psychological functioning.

The present study offers a pioneering exploration of SOC strategies, perceived social support, life satisfaction, and PTSD within the context of the older adults. These findings carry profound implications for healthcare providers, policymakers, and support programs aiming to enhance the psychological well-being of older people in resource-constrained environments. While our novel study closed a research gap and significantly contributed to the body of related knowledge, it has several limitations. SOC is a complex variable encompassing a wide range of strategies that individuals adapt to by affecting different life domains, such as occupational success, career management, physical health and well-being, and interpersonal relationships which our study did not explore these strategies. Therefore, future studies can examime these nuances. However, despite these limitations, our study has advanced the understanding of the mental health challenges faced by senior citizens in resource-constrained settings such as Ekiti State and has made meaningful contributions to geronpsychology. Specifically our findings align with the conservation of resources (COR) theory, a well-established framework in psychology.

Our study’s findings hold critical practical implications for the well-being of senior citizens in Ekiti State. Healthcare providers can benefit from these insights by tailoring mental health interventions to include the promotion of SOC strategies among older individuals. Policymakers should consider allocating resources to bolster mental health support programs for senior citizens, particularly in resource-constrained local government areas of the state. Additionally, support programs may find value in fostering perceived social support and life satisfaction as integral components in reducing the burden of PTSD among senior citizens.

## Supporting information

SOC AND PTSD Tables

## Data Availability

Alldata produced in this study are available upon reasonable request to the corresponding author

https://github.com/jamboyar/Senior-Citizen-in-Ekiti-State

