## Supplementary material for "Examining the indirect effects of life satisfaction and perceived social support on selection optimization compensation and PTSD among the senior citizens of Ekiti State: A moderated mediation approach": SOC AND PTSD Tables

Table 1: Means: Descriptive Statistics of the Variables across Age Groups

| Report | | | | | |
| --- | --- | --- | --- | --- | --- |
| AGE RANGE | | Life satisfaction | Perceived social support | PTSD | SOC  selection optimization compensation |
| 65-74 | Mean | 35.9947 | 52.0842 | 55.7316 | 13.9211 |
|  | N | 190 | 190 | 190 | 190 |
|  | Std. Deviation | 6.13300 | 9.63477 | 13.23042 | 8.30688 |
| 75-84 | Mean | 35.5333 | 49.1200 | 60.0800 | 17.1067 |
|  | N | 75 | 75 | 75 | 75 |
|  | Std. Deviation | 5.68862 | 6.16213 | 7.50632 | 7.23836 |
| 85 AND ABOVE | Mean | 34.9464 | 46.1786 | 57.5000 | 12.5000 |
|  | N | 56 | 56 | 56 | 56 |
|  | Std. Deviation | 5.63889 | 11.26395 | 11.15184 | 9.90867 |
| Total | Mean | 35.7040 | 50.3614 | 57.0561 | 14.4174 |
|  | N | 321 | 321 | 321 | 321 |
|  | Std. Deviation | 5.94319 | 9.51842 | 11.87474 | 8.49891 |

| **Table 2 Correlations in the variables of Age Groups 65-74** | | | | | |
| --- | --- | --- | --- | --- | --- |
|  | | Life satisfaction | Perceived social support | PTSD | SOC |
| Life satisfaction | Pearson Correlation | 1 | .052 | .019 | -.076 |
|  | Sig. (2-tailed) |  | .473 | .795 | .295 |
|  | N | 190 | 190 | 190 | 190 |
| Perceived social support | Pearson Correlation | .052 | 1 | .171^*^ | .104 |
|  | Sig. (2-tailed) | .473 |  | .018 | .154 |
|  | N | 190 | 190 | 190 | 190 |
| PTSD | Pearson Correlation | .019 | .171^*^ | 1 | -.115 |
|  | Sig. (2-tailed) | .795 | .018 |  | .113 |
|  | N | 190 | 190 | 190 | 190 |
| SOC | Pearson Correlation | -.076 | .104 | -.115 | 1 |
|  | Sig. (2-tailed) | .295 | .154 | .113 |  |
|  | N | 190 | 190 | 190 | 190 |
| *. Correlation is significant at the 0.05 level (2-tailed). | | | | | |

Table 3: Correlations for the Age Groups 75-84 Variables

|  | | Life satisfaction | Perceived social support | PTSD | SOC |
| --- | --- | --- | --- | --- | --- |
| Life satisfaction | Pearson Correlation | 1 | .241^*^ | -.117 | -.083 |
|  | Sig. (2-tailed) |  | .037 | .315 | .477 |
|  | N | 75 | 75 | 75 | 75 |
| Perceived social support | Pearson Correlation | .241^*^ | 1 | -.071 | -.404^**^ |
|  | Sig. (2-tailed) | .037 |  | .544 | .000 |
|  | N | 75 | 75 | 75 | 75 |
| PTSD | Pearson Correlation | -.117 | -.071 | 1 | .074 |
|  | Sig. (2-tailed) | .315 | .544 |  | .528 |
|  | N | 75 | 75 | 75 | 75 |
| SOC | Pearson Correlation | -.083 | -.404^**^ | .074 | 1 |
|  | Sig. (2-tailed) | .477 | .000 | .528 |  |
|  | N | 75 | 75 | 75 | 75 |

*. Correlations are significant at the 0.05 level (2-tailed).

| Table 4: Correlations in the variables of Age Groups 85 and above | | | | | |
| --- | --- | --- | --- | --- | --- |
|  | | Life satisfaction | Perceived social support | PTSD | SOC |
| Life satisfaction | Pearson Correlation | 1 | -.096 | .113 | .051 |
|  | Sig. (2-tailed) |  | .480 | .407 | .708 |
|  | N | 56 | 56 | 56 | 56 |
| Perceived social support | Pearson Correlation | -.096 | 1 | -.521^**^ | .539^**^ |
|  | Sig. (2-tailed) | .480 |  | .000 | .000 |
|  | N | 56 | 56 | 56 | 56 |
| PTSD | Pearson Correlation | .113 | -.521^**^ | 1 | -.729^**^ |
|  | Sig. (2-tailed) | .407 | .000 |  | .000 |
|  | N | 56 | 56 | 56 | 56 |
| SOC | Pearson Correlation | .051 | .539^**^ | -.729^**^ | 1 |
|  | Sig. (2-tailed) | .708 | .000 | .000 |  |
|  | N | 56 | 56 | 56 | 56 |
| **. Correlation is significant at the 0.01 level (2-tailed). | | | | | |

| **Table 5: Correlations of the Age Groups of the entire sample** | | | | | |
| --- | --- | --- | --- | --- | --- |
|  | | Life satisfaction | Perceived social support | PTSD | SOC |
| Life satisfaction | Pearson Correlation | 1 | .064 | .008 | -.050 |
|  | Sig. (2-tailed) |  | .251 | .887 | .369 |
|  | N | 321 | 321 | 321 | 321 |
| Perceived social support | Pearson Correlation | .064 | 1 | -.015 | .139^*^ |
|  | Sig. (2-tailed) | .251 |  | .789 | .013 |
|  | N | 321 | 321 | 321 | 321 |
| PTSD | Pearson Correlation | .008 | -.015 | 1 | -.181^**^ |
|  | Sig. (2-tailed) | .887 | .789 |  | .001 |
|  | N | 321 | 321 | 321 | 321 |
| SOC | Pearson Correlation | -.050 | .139^*^ | -.181^**^ | 1 |
|  | Sig. (2-tailed) | .369 | .013 | .001 |  |
|  | N | 321 | 321 | 321 | 321 |
| *. Correlation is significant at the 0.05 level (2-tailed). | | | | | |

One-way ANOVA

| **Table 6: ANOVA Results of the Variables Across Age Groups** | | | | | | | | | | | | | | | | |
| --- | --- | --- | --- | --- | --- | --- | --- | --- | --- | --- | --- | --- | --- | --- | --- | --- |
|  | | | | N | Mean | Std. Deviation | | Std. Error | 95% Confidence Interval for Mean | | | | Minimum | | Maximum | |
|  |  |  |  |  |  |  |  |  | Lower Bound | | | Upper Bound |  |  |  |  |
| Life satisfaction | | 65-74 | | 190 | 35.9947 | 6.13300 | | .44493 | 35.1171 | | | 36.8724 | 21.00 | | 51.00 | |
|  |  | 75-84 | | 75 | 35.5333 | 5.68862 | | .65686 | 34.2245 | | | 36.8422 | 24.00 | | 50.00 | |
|  |  | 85 AND ABOVE | | 56 | 34.9464 | 5.63889 | | .75353 | 33.4363 | | | 36.4565 | 21.00 | | 51.00 | |
|  |  | Total | | 321 | 35.7040 | 5.94319 | | .33172 | 35.0514 | | | 36.3567 | 21.00 | | 51.00 | |
| Perceived social support | | 65-74 | | 190 | 52.0842 | 9.63477 | | .69898 | 50.7054 | | | 53.4630 | 35.00 | | 77.00 | |
|  |  | 75-84 | | 75 | 49.1200 | 6.16213 | | .71154 | 47.7022 | | | 50.5378 | 34.00 | | 59.00 | |
|  |  | 85 AND ABOVE | | 56 | 46.1786 | 11.26395 | | 1.50521 | 43.1621 | | | 49.1951 | 30.00 | | 69.00 | |
|  |  | Total | | 321 | 50.3614 | 9.51842 | | .53127 | 49.3162 | | | 51.4066 | 30.00 | | 77.00 | |
| PTSD | | 65-74 | | 190 | 55.7316 | 13.23042 | | .95984 | 53.8382 | | | 57.6249 | 17.00 | | 74.00 | |
|  |  | 75-84 | | 75 | 60.0800 | 7.50632 | | .86676 | 58.3530 | | | 61.8070 | 44.00 | | 69.00 | |
|  |  | 85 AND ABOVE | | 56 | 57.5000 | 11.15184 | | 1.49023 | 54.5135 | | | 60.4865 | 24.00 | | 66.00 | |
|  |  | Total | | 321 | 57.0561 | 11.87474 | | .66278 | 55.7521 | | | 58.3600 | 17.00 | | 74.00 | |
| SOC | | 65-74 | | 190 | 13.9211 | 8.30688 | | .60264 | 12.7323 | | | 15.1098 | .00 | | 36.00 | |
|  |  | 75-84 | | 75 | 17.1067 | 7.23836 | | .83581 | 15.4413 | | | 18.7721 | 5.00 | | 31.00 | |
|  |  | 85 AND ABOVE | | 56 | 12.5000 | 9.90867 | | 1.32410 | 9.8464 | | | 15.1536 | .00 | | 36.00 | |
|  |  | Total | | 321 | 14.4174 | 8.49891 | | .47436 | 13.4842 | | | 15.3507 | .00 | | 36.00 | |
|  | **Table 7: ANOVA Assessing Statistical Differences Between Age Groups for Variables** | | | | | | | | | | | | | | | |
|  |  | | | | | | Sum of Squares | | | df | Mean Square | | | F | | Sig. |
|  | Life satisfaction | | Between Groups | | | | 50.384 | | | 2 | 25.192 | | | .712 | | .491 |
|  |  |  | Within Groups | | | | 11252.501 | | | 318 | 35.385 | | |  | |  |
|  |  |  | Total | | | | 11302.885 | | | 320 |  | | |  | |  |
|  | Perceived social support | | Between Groups | | | | 1659.294 | | | 2 | 829.647 | | | 9.652 | | .000 |
|  |  |  | Within Groups | | | | 27332.787 | | | 318 | 85.952 | | |  | |  |
|  |  |  | Total | | | | 28992.081 | | | 320 |  | | |  | |  |
|  | PTSD | | Between Groups | | | | 1030.160 | | | 2 | 515.080 | | | 3.715 | | .025 |
|  |  |  | Within Groups | | | | 44092.831 | | | 318 | 138.657 | | |  | |  |
|  |  |  | Total | | | | 45122.991 | | | 320 |  | | |  | |  |
|  | SOC | | Between Groups | | | | 795.100 | | | 2 | 397.550 | | | 5.664 | | .004 |
|  |  |  | Within Groups | | | | 22318.962 | | | 318 | 70.185 | | |  | |  |
|  |  |  | Total | | | | 23114.062 | | | 320 |  | | |  | |  |

| Multiple Comparisons | | | | | | | |
| --- | --- | --- | --- | --- | --- | --- | --- |
| Table 8: Post Hoc Test (Scheffe) of the mean difference between Age Groups of the variables | | | | | | | |
| Dependent Variable | (I) AGERANGE | (J) AGERANGE | Mean Difference (I-J) | Std. Error | Sig. | 95% Confidence Interval | |
|  |  |  |  |  |  | Lower Bound | Upper Bound |
| Life satisfaction | 65-74 | 75-84 | .46140 | .81120 | .851 | -1.5336 | 2.4564 |
|  |  | 85 AND ABOVE | 1.04831 | .90450 | .512 | -1.1761 | 3.2728 |
|  | 75-84 | 65-74 | -.46140 | .81120 | .851 | -2.4564 | 1.5336 |
|  |  | 85 AND ABOVE | .58690 | 1.05056 | .856 | -1.9968 | 3.1706 |
|  | 85 AND ABOVE | 65-74 | -1.04831 | .90450 | .512 | -3.2728 | 1.1761 |
|  |  | 75-84 | -.58690 | 1.05056 | .856 | -3.1706 | 1.9968 |
| Perceived social support | 65-74 | 75-84 | 2.96421 | 1.26428 | .066 | -.1451 | 6.0735 |
|  |  | 85 AND ABOVE | 5.90564^*^ | 1.40969 | .000 | 2.4387 | 9.3725 |
|  | 75-84 | 65-74 | -2.96421 | 1.26428 | .066 | -6.0735 | .1451 |
|  |  | 85 AND ABOVE | 2.94143 | 1.63734 | .201 | -1.0853 | 6.9682 |
|  | 85 AND ABOVE | 65-74 | -5.90564^*^ | 1.40969 | .000 | -9.3725 | -2.4387 |
|  |  | 75-84 | -2.94143 | 1.63734 | .201 | -6.9682 | 1.0853 |
| PTSD | 65-74 | 75-84 | -4.34842^*^ | 1.60578 | .027 | -8.2975 | -.3993 |
|  |  | 85 AND ABOVE | -1.76842 | 1.79047 | .614 | -6.1718 | 2.6349 |
|  | 75-84 | 65-74 | 4.34842^*^ | 1.60578 | .027 | .3993 | 8.2975 |
|  |  | 85 AND ABOVE | 2.58000 | 2.07961 | .464 | -2.5344 | 7.6944 |
|  | 85 AND ABOVE | 65-74 | 1.76842 | 1.79047 | .614 | -2.6349 | 6.1718 |
|  |  | 75-84 | -2.58000 | 2.07961 | .464 | -7.6944 | 2.5344 |
| SOC | 65-74 | 75-84 | -3.18561^*^ | 1.14245 | .021 | -5.9953 | -.3760 |
|  |  | 85 AND ABOVE | 1.42105 | 1.27386 | .537 | -1.7118 | 4.5539 |
|  | 75-84 | 65-74 | 3.18561^*^ | 1.14245 | .021 | .3760 | 5.9953 |
|  |  | 85 AND ABOVE | 4.60667^*^ | 1.47957 | .008 | .9679 | 8.2454 |
|  | 85 AND ABOVE | 65-74 | -1.42105 | 1.27386 | .537 | -4.5539 | 1.7118 |
|  |  | 75-84 | -4.60667^*^ | 1.47957 | .008 | -8.2454 | -.9679 |
| *. The mean difference is significant at the 0.05 level. | | | | | | | |

**Table 9: Impact of the SOC on the PSS**

| R = 0.14 | R-sq = 0.02 | MSE = 89.14 | F = 6.24 | df1 = 1.00 | df2 = 319.00 | P = 0.01 |
| --- | --- | --- | --- | --- | --- | --- |
| Model |  |  |  |  |  |  |
|  | Coeff | se | t | p | LLCI | ULCI |
| Constant | 50.36 | 0.53 | 95.57 | 0.00 | 49.32 | 51.40 |
| SOC | 0.16 | 0.06 | 2.50 | 0.01 | 0.03 | 0.28 |

Table 10: Impact of the SOC on life satisfaction

| R = 0.05 | R-sq = 0.00 | MSE = 35.34 | F = 0.81 | df1 = 1.00 | df2 = 319.00 | P = 0.37 |
| --- | --- | --- | --- | --- | --- | --- |
| Model |  |  |  |  |  |  |
|  | Coeff | Se | t | p | LLCI | ULCI |
| Constant | 35.70 | 0.33 | 107.60 | 0.00 | 35.05 | 36.36 |
| SOC | -0.04 | 0.04 | -0.90 | 0.37 | -0.11 | 0.04 |

*Outcome Variable: Life satisfaction; N = 321*

**Table 11: Model Summary**

R	R-sq	MSE	F	df1	df2	p

.25	.06	134.04	4.33	5.00	315.00	.00

coeff	se	t	p	LLCI	ULCI

constant	52.89	5.22	10.13	.00	42.62	63.15

SOC	-.24	.08	-3.08	.00	-.39	-.09

PSS	.07	.07	.98	.33	-.07	.21

Life_Sa	.02	.11	.16	.87	-.20	.23

Age	1.37	.87	1.58	.11	-.34	3.08

Int_1	-.25	.09	-2.71	.01	-.43	-.07

Product terms key:

Int_1	: SOC	x	Age

Test(s) of highest order unconditional interaction(s):

R2-chng	F	df1	df2	p

X*W	.02	7.32	1.00	315.00	.01

Table 12: Conditional direct effect(s) of age on the association between SOC and PTSD

| Age | Effect | se | t | p | LLCI | ULCI |
| --- | --- | --- | --- | --- | --- | --- |
| -0.58 | -0.09 | 0.10 | -0.95 | 0.35 | -0.29 | 0.10 |
| 0.00 | -0.24 | 0.08 | -3.08 | 0.00 | -0.39 | -0.09 |
| 0.77 | -0.43 | 0.10 | -4.31 | 0.00 | -0.63 | -0.23 |

Table 13: Mediation analysis summary

| Total Effect (SOC -> PTSD) | Direct Effect  (SOC -> PTSD) | Relationship | Indirect Effect | Confidence Interval | | t-statistics | Conclusion |
| --- | --- | --- | --- | --- | --- | --- | --- |
|  |  |  |  | Lower Bound | Upper Bound |  |  |
| -0.23 (000) | -0.24 (000) | SOC -> PSS -> PTSD | 0.011 | -0.02 | 0.059 | 2.348 | Partial mediation |
|  |  | SOC -> Life Satisfaction -> PTSD | -0.001 | 0.022 | 0.009 | 2.329 | Partial mediation |
